# Diurnal brain temperature rhythms and mortality after brain injury: a prospective and retrospective cohort study

**DOI:** 10.1101/2021.01.23.21250327

**Authors:** Nina M Rzechorzek, Michael J Thrippleton, Francesca M Chappell, Grant Mair, Ari Ercole, Manuel Cabeleira, The CENTER-TBI High Resolution ICU (HR ICU) Sub-Study Participants and Investigators, Jonathan Rhodes, Ian Marshall, John S O’Neill

**Affiliations:** MRC Laboratory of Molecular Biology, Francis Crick Avenue, Cambridge, CB2 0QH, UK; Edinburgh Imaging (Royal Infirmary of Edinburgh) Facility, 51 Little France Crescent, Edinburgh, EH16 4SA UK; Centre for Clinical Brain Sciences, University of Edinburgh, 49 Little France Crescent, Edinburgh, EH16 4SB UK; Division of Anaesthesia, Department of Medicine, Cambridge University Hospitals NHS Foundation Trust, University of Cambridge, Box 93, Addenbrooke’s Hospital, Hills Road, Cambridge, CB2 0QQ UK; Division of Neurosurgery, Department of Clinical Neurosciences, University of Cambridge, Box 167, Cambridge Biomedical Campus, Addenbrooke’s Hospital, Cambridge, CB2 0QQ UK; Department of Anaesthesia, Critical Care and Pain Medicine NHS Lothian, Room No. S8208 (2nd Floor), Royal Infirmary of Edinburgh, 51 Little France Crescent, Edinburgh, EH16 4SA UK

## Abstract

**Objective:** To determine the clinical relevance of brain temperature (*T*_Br_) variation in patients after traumatic brain injury (TBI).

**Design:** Cohort study with prospective (healthy participant) and retrospective (TBI patient) arms.

**Setting:** Single neuroimaging site in the UK (prospective arm); intensive care sites contributing to the Collaborative European NeuroTrauma Effectiveness Research in TBI (CENTER-TBI) High Resolution ICU (HR ICU) Sub-Study (retrospective arm).

**Participants:** 40 healthy adults aged 20-40 years recruited for non-invasive brain thermometry and all patients up to May 2020 that had *T*_Br_ measured directly and were not subjected to Targeted Temperature Management (TTM).

**Main outcome measures:** A diurnal change in *T*_Br_ (healthy participants); death in intensive care (patients).

**Results:** In healthy participants, mean *T*_Br_ (38.5 SD 0.4°C) was higher than oral temperature (36.0 SD 0.5°C), and 0.36°C higher in luteal females relative to follicular females and males (95% confidence interval 0.17 to 0.55, P=0.0006 and 0.23 to 0.49, P<0.0001, respectively). *T*_Br_ increased with age, most notably in deep brain regions (0.6°C over 20 years; 0.11 to 1.07, P=0.0002). The mean maximal spatial *T*_Br_ range was 2.41 (SD 0.46)°C, with highest temperatures in the thalamus. *T*_Br_ varied significantly by time of day, especially in deep brain regions (0.86°C; 0.37 to 1.26, P=0.0001), and was lowest in the late evening. Diurnal *T*_Br_ in cortical white matter across participants ranged from 37.0 to 40.3°C. In TBI patients (n=114), mean *T*_Br_ (38.5 SD 0.8°C) was significantly higher than body temperature (*T*_Bo_ 37.5 SD 0.5°C; P<0.0001) and ranged from 32.6 to 42.3°C. Only 25/110 patients displayed a diurnal temperature rhythm; *T*_Br_ amplitude was reduced in older patients (P=0.018), and 25/113 patients died in intensive care. Lack of a daily *T*_Br_ rhythm, or an age increase of 10 years, increased the odds of death 12-fold and 11-fold, respectively (OR for death with rhythm 0.09; 0.01 to 0.84, P=0.035 and for death with ageing by 1 year 1.10; 1.05 to 1.16, P=0.0002). Mean *T*_Br_ was positively associated with survival (OR for death 0.45 for 1°C increase; 0.21 to 0.96, P=0.040).

**Conclusions:** Healthy *T*_Br_ exceeds *T*_Bo_ and varies by sex, age, menstrual cycle, brain region, and time of day. Our 4-dimensional reference resource for healthy *T*_Br_ can guide interpretation of *T*_Br_ data in multiple clinical settings. Daily temperature variation is frequently disrupted or absent in TBI patients, in which *T*_Br_ variation is of greater prognostic use than absolute *T*_Br_. Older TBI patients lacking a daily *T*_Br_ rhythm are at greatest risk of death in intensive care. Appropriately controlled trials are needed to confirm the predictive power of *T*_Br_ rhythmicity in relation to patient outcome, as well as the clinical utility of TTM protocols in brain-injured patients.

**Registration:** UK CRN NIHR CPMS 42644; ClinicalTrials.gov number, NCT02210221.

**SUMMARY BOX:** *What is already known on this topic:*

- Brain temperature (*T*_Br_) can be measured directly in brain-injured patients via intracranial probe, but this method cannot be used in healthy individuals.
- *T*_Br_ can be measured non-invasively using magnetic resonance spectroscopy (MRS), but this method is not appropriate for most brain-injured patients.
- Since physiological reference ranges for *T*_Br_ in health have not been established, the clinical relevance of *T*_Br_ variation in patients is unknown, and the use of TTM in neurocritical care remains controversial.

*What this study adds:*

- A reference map for healthy adult *T*_Br_ at three clinically-relevant time points that can guide interpretation of *T*_Br_ measured directly, or by MRS, in multiple clinical settings.
- Our results suggest that loss of diurnal *T*_Br_ rhythmicity after TBI increases the odds of intensive care death 12-fold; some TTM strategies may be clinically inappropriate.

## INTRODUCTION

Elevated temperature has been recognized as a sign of disease for more than two millennia.^1^ Both the spatial and temporal dynamics of temperature contain additional diagnostic information, exemplified by malarial fever cycles and local warming at sites of injury or infection.^2-9^ Internal organ temperature is rarely measured directly since invasive methods are required; in practice, temperature is assumed to be uniform throughout the brain and body core, overlooking the true clinical value of regional temperature measurements in individual tissues. Brain cell function is unequivocally temperature-dependent however,^10^ and it is accepted that absolute *T*_Br,_ its relationship to *T*_Bo_, and the apparent temperature-sensitivity of brain tissue are frequently altered following injury.^3,7,8,11,12^ Indeed, understanding of human *T*_Br_ has largely been informed by studies of brain-injured patients, where intracranial probes allow precise (±0.1–0.3°C), direct measurement from a single brain locus.^13,14^

The temperature-dependence of brain function has perpetuated the assumption that *T*_Br_ must be relatively homogenous and static in health. However, several lines of evidence suggest that *T*_Br_ may vary over time, and between brain regions.^4-5,7,9,11,12,15-19^ For example, human core *T*_Bo_ is 1-2°C lower during sleep at night, when cerebral blood flow is also ∼20% higher^20,21^; therefore, brain heat removal should be more efficient at night than during the day. Moreover, direct measurements in non-human primates show that deep brain structures are warmer than the brain surface, and that *T*_Br_ varies at least as much as *T*_Bo_ across a 24-hour period.^18,19^

Establishing how *T*_Br_ varies in health is critical; deviations from normal may have transformative diagnostic and/or prognostic value in neurological disease and injury, but only if these deviations can be distinguished from physiological variation over time.^22^ With magnetic resonance spectroscopy (MRS), spatially resolved *T*_Br_ data can now be obtained through non-invasive brain imaging.^13^ Brain thermometry has proven to be a powerful research application of MRS but, with respect to healthy humans, it has only been used in studies that were poorly controlled for parameters that influence physiological temperature variation (Supplementary Table S1). We sought to establish the daily spatiotemporal variation of healthy *T*_Br_ to enable evidence-driven appraisal of the clinical value of *T*_Br_ monitoring in brain-injured patients. We hypothesized that healthy *T*_Br_ would vary diurnally, and that disruption of diurnal temperature variation would be associated with outcome after TBI.

## METHODS

Reporting adheres to STROBE guidelines.

### Prospective study design and recruitment

We conducted a prospective, single-site, cohort study in healthy adults, controlled for age, sex, body mass index (BMI), menstrual cycle phase, seasonal variation, and individual chronotype. Our primary objective was to determine whether healthy *T*_Br_ varies by time of day. Our secondary objectives were to compare variability in brain and oral temperatures, to test for differences between males and luteal-phase females, and explore brain-regional changes in *T*_Br_ with time. We hypothesized that *T*_Br_ would (i) exceed and vary more than oral temperature across the day, (ii) be higher in luteal females relative to males, and (iii) increase with increasing brain tissue depth. Sample size was estimated for achieving the primary outcome (a change in mean global *T*_Br_ between time points) using a linear mixed model, considering published data on the reliability of MRS brain thermometry in healthy men.^13^ With 36 subjects, and a conservative true mean *T*_Br_ difference of 0.5°C, we estimated 80% power to detect a statistically significant difference between time points at the 5% significance level. A health-related finding (HRF) on MRI was the key exclusion criterion. Completion of a feedback pathway for two volunteers was expected (based on 5% prevalence of HRFs using high-resolution MRI).^23^ We aimed to scan 40 eligible participants (20 females) to account for potential withdrawal, exclusion, and/or technical scan failure.

Recruitment for our Circadian Brain Temperature (CiBraT) Study was based on meeting criteria for our primary outcome (Supplementary Table S2), and was conducted locally using mailshots to University of Edinburgh and NHS staff, social media posts, and posters displayed at University of Edinburgh campuses and NHS Lothian hospitals. By completing an online eligibility questionnaire, all prospective participants provided written informed consent for their personal data to be used to schedule consenting interviews, and to notify general practitioners of their intention to participate. The questionnaire provided access to inclusion and exclusion criteria, the Participant Information Sheet and Consent to Participate Form (Supplementary Appendix 1), and Data Protection Information sheet. All participants provided written, informed consent to participate during face-to-face interview conducted by the Chief Investigator (NMR) at the University of Edinburgh. Additional written informed consent was obtained for publication of individual data which, by nature of its distinctive features, could potentially be recognized by participants as their own data. The Study Protocol is presented in Supplementary Appendix 2.

### Prospective data collection

During a consenting interview at the study site, one week in advance of scanning, each participant was given a wrist-worn actimeter (ActTrust2, Condor Instruments, Sao Paulo, Brazil). Each participant then underwent three identical brain scans in the morning (9-10am), afternoon (4-5pm), and late evening (11pm-midnight) of their scheduled scanning day. Multiple time points spanning >12 hours were selected because the human circadian rhythm (body clock) impacts almost every aspect of physiology (Supplementary Text *‘Internal rhythms and health’*).^24-27^ The exact alignment, or phase relationship, between the body clock and the day-night cycle is dictated by individual chronotype, which is determined by genetic and lifestyle factors, and can be derived from longitudinal monitoring of locomotor activity.^28^ To assign scan times to the appropriate part of each participant’s circadian cycle, we determined individual chronotypes using wrist actigraphy to extract the sleep-corrected midpoint of sleep on free (non-work) days (MSF_sc_) (Supplementary Methods).^28^ Height and weight were measured immediately before the morning scan to calculate BMI. Oral temperature was recorded before each scanning session using a digital Clinical Thermometer (S.Brannan & Sons, UK) covered in a single-use Probe Cover (Bunzl Retail & Healthcare Supplies Limited, Middlesex, UK) and placed sublingually. For females, hormonal influences were controlled through urine-based ovulation testing (ClearBlue®), or documenting hormonal contraception type. We aimed to scan females during the luteal phase of their natural menstrual cycle, or on a day when an active combined pill would be taken, or combined patch worn. Females using other forms of contraception (implant or intrauterine device) were excluded. On the day of scanning, food consumption was restricted to 6am–8am, 12noon to 2pm, and 6pm–8pm, and caffeine consumption was restricted to 6am–8am and 12noon to 2pm. Alcohol was strictly prohibited at all times. Participants were asked not to participate in excessive physical activity on the day of scanning. Data collection was limited to a 14-week period between July and October 2019 to avoid daylight savings clock changes and large seasonal variation in environmental light and temperature conditions. Data management procedures are described in Supplementary Methods.

### Brain imaging

All brain imaging was conducted at the Edinburgh Imaging (Royal Infirmary of Edinburgh) Facility using a 3-T MAGNETOM Prisma scanner (Siemens Healthcare, Erlangen, Germany) with a 32-channel head coil. All participants were screened for MRI contraindications and changed into hospital scrubs for each scanning session—conducted in a temperature-controlled room (target 21.5°C). Room lights were off and the scanner lighting and fan were maintained on their lowest setting. Ear protection was provided and a mirror was attached to the head coil so participants had the choice of closing their eyes or viewing the MRI control room; no visual or acoustic entertainment was provided. Participants were permitted to sleep during scans and were asked to report on this event at the end of each session. At each time point, after whole-brain structural MRI, MRS data were collected from 82 brain locations (voxels). The scanning protocol was well tolerated, with no serious adverse events reported during 7-day follow-up. Further details on the scanning protocol and MRS data processing are provided in Supplementary Methods and Appendix 3; the dedicated Study Participant Data Form (Case Report Form) is provided in Appendix 4.

### Calculation of MRS-derived brain temperature

MRS brain thermometry exploits the fact that the chemical shift of water is exquisitely temperature-dependent (−0.01ppm/°C), whilst that of the reference metabolite NAA is not.^29^ The chemical shift difference between water and NAA can estimate absolute *T*_Br_ in healthy men with a short-term precision of 0.14°C at 3-T.^13^ *T*_Br_ for each brain tissue voxel in this study was calculated using the following relationship:

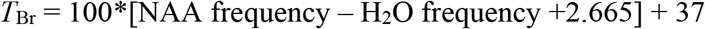

where frequency is in ppm and temperature is in °C

The reliability and accuracy of *T*_Br_ determination using this MRS protocol was thoroughly tested using *in vivo* human and *in vitro* phantom measurements; the latter validated with an MR-compatible industrial thermometer that meets international standards.^13^

### Retrospective study design and patient data sources

To determine the clinical relevance of *T*_Br_ variation, we conducted a multicentre, retrospective cohort study of TBI patients that had high temporal-resolution *T*_Br_ data collected directly from the brain. Data for all eligible patients were extracted using version 2.0 of the CENTER-TBI dataset, compiled between 2015 and 2017. Additional eligible patients monitored at one of the contributing sites (the Intensive Care Unit, Western General Hospital, Edinburgh, UK) were included up to May 2020 and comprised 109 of the 134 eligible patients screened. The Western General Hospital is the tertiary referral centre in South East Scotland for neurosurgical emergencies. Patients with moderate to severe TBI admitted to intensive care requiring intubation, sedation, and intracranial pressure (ICP) management also received brain oxygen tension and temperature monitoring using the Integra Licox system (Integra, France). Patients were managed in accordance with Brain Trauma Foundation guidelines.^30^ Patients were either admitted directly to intensive care or following surgical intervention for mass lesions. *T*_Br_ was measured via a thermistor, inserted into the brain parenchyma via a dedicated bolt placed via a burr hole (Integra Neurosciences, Andover, UK). The bolt was placed so that the thermistor inserted into frontal white matter; for diffuse injuries this was into the non-dominant hemisphere. When the main injury was focal, the bolt was placed on the side of maximal injury, unless this would place the monitors into non-viable tissue. High temporal-resolution physiological data were recorded at a minimum of 1-minute intervals to either a bedside computer running ICU Pilot software (CMA, Sweden) or to a Moberg neuromonitoring system (Moberg Research Inc., USA). Data were collected continuously (except for interruptions due to computed tomography scanning or surgical intervention) and until ICP monitoring was no longer required, or the patient died. Data for the CENTER-TBI study were collected through the Quesgen e-CRF (Quesgen Systems Inc, USA), hosted on the INCF platform and extracted via the INCF Neurobot tool (INCF, Sweden). For patient monitoring and data collection in the High-Resolution repository, the ICM+ platform (University of Cambridge, UK) and/or Moberg Neuromonitoring system (Moberg Research Inc., USA) were used. For *T*_Bo_, the primary method of measurement was documented in 26 of 134 screened patients and included tympanic (21), bladder (3), external axillary (1), and nasopharyngeal (1). Secondary sites included rectal, external axillary, oesophageal, and skin.

### Patient temperature data processing

Four inclusion criteria levels were applied to ensure that sufficient temperature data were available to assess for a diurnal rhythm (Table 1), and that any data potentially affected by TTM protocols were excluded. Analysis of patient temperature data was blinded to outcome. Data from the first 2 hours of monitoring were excluded from the analysis to ensure the results were not influenced by the time required for the electrode to stabilise. Raw data processing was performed in Excel to exclude artefactual data, identify any gaps in the time series, and define the analysis window. Temperature data were visualized in GraphPad Prism version 8.2 and assessed for the presence of daily rhythmicity. Visual analyses were validated with a combination of rhythm-detection algorithms using GraphPad Prism, BioDare2 (biodare2.ed.ac.uk)^31^ and the Harmonic Regression package in R.^32^ To be categorized as diurnally rhythmic, the patient’s temperature pattern need not be conventionally aligned with the day-night cycle, but it had to meet both of the following criteria:

**Table 1.** Inclusion criteria for retrospective analysis of temperature data from TBI patients. TTM, Targeted Temperature Management; ICP, Intracranial pressure; PbTO_2_, partial pressure of brain oxygen; MAP, mean arterial pressure, CT, computed tomography; MRI, Magnetic Resonance Imaging; IMPACT, International Mission for Prognosis and Analysis of Clinical Trials in TBI; GCS, Glasgow Coma Score, GCSM, Glasgow Coma Score Motor response; TIL, Therapy Intensity Level; GOSE, Glasgow Outcome Scale Extended.

|  |
| --- |
| **Level A: Criteria for extracting maximum and minimum daily brain and/or body temperatures** |
| • Known sex<br>• Known age<br>• Minimum 24 hours of temperature data collection under ‘constant’ conditions. The first data point recorded in the intensive care setting that exceeds the minimum recorded temperature from that patient in the absence of TTM will be taken as the start point (to exclude low temperature points surrounding insertion of probe or those relating to patient hypothermia on arrival in intensive care)<br>• For data where only the maximum and minimum daily $T_{Bo}$ (and in some cases $T_{Br}$) are recorded with their respective times, a minimum of two days’ worth of data is needed<br>• If TTM was applied, only data relating to time preceding TTM or after the first inflection of data after cessation of TTM can be used and must meet the above requirements for minimum time length in the absence of TTM<br>• When extracting the time of the minimum and maximum temperature point, the first occurrence of that specific temperature point under intensive care ‘constant’ conditions will be selected |
| **Level B: Additional criteria for performing diurnal rhythmic temperature analyses** |
| • Minimum hourly $T_{Bo}$ or $T_{Br}$ data with $T_{Br}$ data extracted via intracranial probe (standard depth and positioning in cortical white matter) recorded continuously over a minimum of 36 hours. The same rules as above apply in relation to TTM.<br>• Ideally minimum hourly data of another matched parameter (ICP, $PbTO_2$, MAP) with expected diurnal rhythm |
| **Level C: Additional criteria for correlation with outcome** |
| • Mortality/survival in intensive care<br>• Ideally GOSE at 3 and/or 6 months (imputed where necessary) |
| **Level D: Additional criteria for correlation with injury severity** |
| • One of more of the following parameters: presence of pupillary light reflex in one/both/no eyes; GCS, GCSM<br>• Ideally injury type (focal/diffuse; from CT scoring) and/or severity (IMPACT imputed GCS) on admission to Study Hospital and/or TIL score (including individual components of this)<br>• Ideally site of probe insertion for focal injury (ipsilateral or contralateral to injury—to be determined using CT/MRI images if available) |

1. a period length of ∼22–26h in at least part (but not necessarily all) of the time series as determined by blinded visual analysis of the raw data in GraphPad Prism and
2. a period length of 22-26h as determined by period analysis in (i) cosinor analysis in GraphPad Prism and/or (ii) statistically significant output from Harmonic Regression in R and/or (iii) BioDare2.

In GraphPad Prism, period results were only considered valid if a cosinor curve fit was significantly preferred over a straight line. When using the Harmonic Regression package, the period length term (Tau) of the model to test for was set to 24 hours. In BioDare2, period analysis was performed using 6 different algorithms. A full description of these algorithms can be found at https://biodare2.ed.ac.uk/documents/period-methods.

### Statistical analysis

To determine healthy temperature variation, we applied a linear mixed modelling approach. The fixed effects (predictors) were specified *a priori* based on published literature describing factors that were most likely to affect body and/or brain temperature in humans and other mammals.^22^ In each case, the upper limit for the number of fixed effects was set to a maximum of five to avoid over-fitting each model within the confines of our sample size.^33^ Random effects for intercept and slope were included, allowing participants to have different baseline temperatures and different changes in temperature over time. The models for oral temperature (*OralTemp*) and *T*_Br_ (*BrainTemp*) were built as follows:

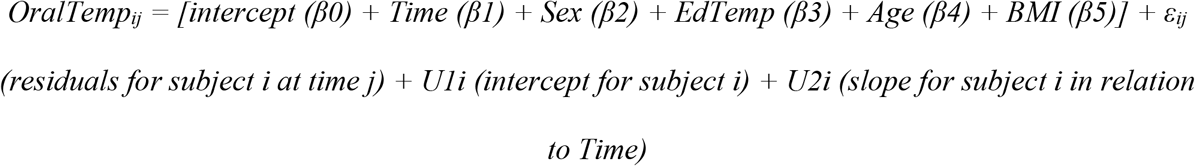

where fixed effects include:

*Time* (time of day normalized for chronotype using the ‘time distance’ between the *T*_Oral_ measurement and MSF_sc_ for that participant as a proportion of a linearized unit circle where 0=MSF_sc_ and 1=24 hours)

*Sex* (participant biological sex categorized as male, luteal female, or non-luteal female) *EdTemp* (environmental temperature in Edinburgh on that date and at the time of temperature measurement)

*Age* (participant age on date of temperature measurement)

*BMI* (participant BMI on date of temperature measurement)

with random effects for intercept by subject, and for slope by subject with respect to Time

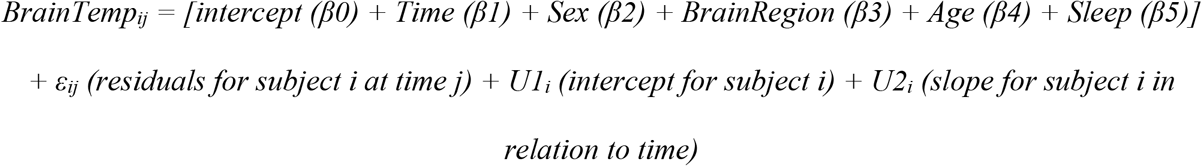

where fixed effects include:

*Time* (time of day normalized for chronotype using the ‘time distance’ between the *T*_Br_ measurement and MSF_sc_ for that participant as a proportion of a linearized unit circle where 0=MSF_sc_ and 1=24 hours)

*Sex* (participant biological sex categorized as male, luteal female, or non-luteal female) *BrainRegion* (brain voxel categorized to one of six regions: Superficial 1, Superficial 2, Superficial 3, Superficial 4, Thalamus, Hypothalamus)

*Age* (participant age on date of temperature measurement)

*Sleep* (whether participant reported falling asleep during scanning; categorized as ‘yes’, ‘maybe’ or ‘no’)

with random effects for intercept by subject, and for slope by subject with respect to Time To confirm that there was no relationship between *T*_Br_ and BMI, the model was run a second time, but replacing the *Sleep* effect with *BMI*. The model for deep *T*_Br_ was identical to the *BrainTemp* model above except that only thalamic and hypothalamic regions were included (Supplementary Appendix 5).

For retrospective data, a generalized linear mixed model (GLMM) for logit binomial distribution of patient outcome was chosen (the rationale for model choice is provided in Supplementary Methods). Survival in intensive care or ‘alive’ was specified as a miss, and death or ‘dead’ as a hit. The model incorporated fixed effects and random effect for intercept and was built as follows:

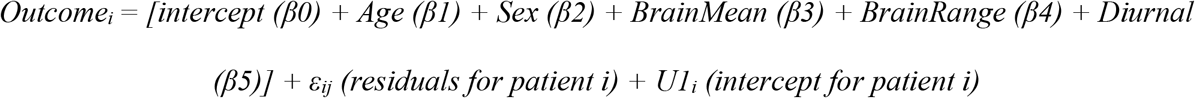

where fixed effects include:

*Age* (patient age in intensive care)

*Sex* (patient biological sex categorized as male or female)

*BrainMean* (absolute mean *T*_Br_ throughout analysis window)

*BrainRange* (*T*_Br_ range across analysis window)

*Diurnal* (presence or absence of a daily temperature rhythm within analysis window—categorized as ‘yes’ or ‘no’; see above for details on how tests for diurnal rhythmicity were performed) with random effects for intercept by subject

The final choice of fixed effects (predictors) to include in the model was based on our core study objectives, avoiding redundant terms, and optimising the model fit (Appendix 5). Missing data values for any of the model components were input as ‘NA’, and thus patients with values missing for one or more of the components were excluded from the model output. The most conservative approach was taken i.e. multiple imputation was not performed since the random nature of missing data could not be assumed.

Statistical modelling and other circular analyses were performed using R version 3.6.3 (R Core Team, 2020) and the circular (v0.4–93; Lund et al., 2017); cosinor (v1.1; Sachs 2015), cosinor2 (v0.2.1; Mutak 2018), lme4 (v1.1–23; Bates et al. 2020), effects (v4.1-4; Fox et al. 2019), afex (Singmann et al., 2020), Matrix (v1.2-18; Bates et al., 2019), Cairo (v1.5-12.2; Urbanek and Horner 2020), yarrr (v0.1.5; Phillips 2017) and car (v3.0-8; Fox et al., 2020) packages. The full reproducible code is provided in Supplementary Appendix 5, or is available on request to the Lead Author. All other analyses were performed in GraphPad Prism version 8.2.

### Patient and public involvement

Edinburgh Imaging staff were actively involved in the design of the prospective study. Pre-existing patient temperature data helped to inform the selection of most appropriate time points for scanning in healthy participants. TBI patients were not actively involved in study design since their data was anonymised prior to extraction and analysis. Healthy participants were invited to outline reasons for non-willingness to participate; where appropriate, this information was used to refine the recruitment approach. The findings of this study will be disseminated via Open Access publication. Conference presentations and public engagement activities will be used to explain the purpose of the research and its potential future impact. End of Study Information Sheets will be emailed to individual healthy participants who gave consent to receive these, and a lay summary of the study results will be posted online after publication.

## RESULTS

### Spatiotemporal measurements of healthy brain temperature

Of the 77 volunteers screened for eligibility, we recruited 20 males and 20 females (aged 20–40 years) between July and September 2019 (Fig.1A). Participants represented 15 nationalities across five continents, and the last participant was scanned on October 8^th^ 2019. One male attended only for morning scanning and another male volunteer missed afternoon scanning; available data from both of these participants was included in the analysis. Of the females scanned, 11 had natural menstrual cycles, eight were taking a combined contraceptive pill and seven of these took an ‘active’ pill on the day of scanning. The female subject on a ‘pill break’ reported day one of menstruation at their afternoon scan; their *T*_Br_ data was included in the luteal group. Of the females with natural cycles, six were confirmed luteal (urine test), two were in menstruation, and three were in non-menstrual follicular phase at scanning. Five females thus formed a non-luteal group. One female wore a combined contraceptive patch on the scanning day (transiently removed during each scan); their *T*_Br_ data was included in the luteal group (fig.S5).

**Fig. 1.**
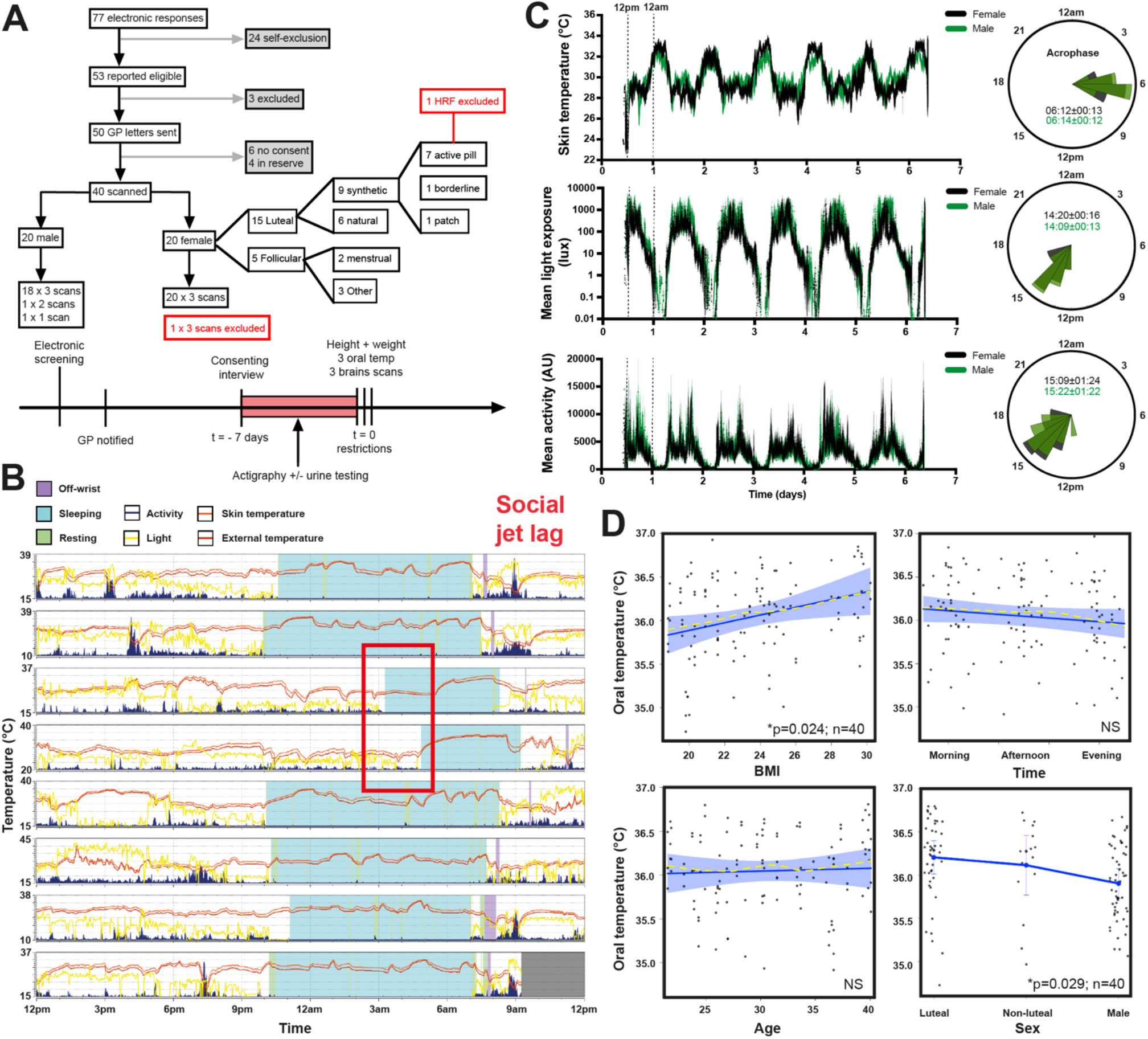
Chronotype-controlled temperature variations. (**A**) Prospective study profile and workflow. (**B**) Representative actogram displaying typical actigraphy over one week from one male volunteer. Horizontal panels represent consecutive days. Note absence of light exposure and activity, with increase in skin temperature during sleep time (activity also absent when device was ‘off-wrist’). Social jet lag refers to large delay in sleeping schedule due to social activities on two consecutive days, highlighted (red box). (**C**) Group-averaged mean±SEM data for distal skin (wrist) temperature, total light exposure, and activity by sex during actigraphy week (left). Females n=20, males n=20. Associated rose plots with circular means (acrophases±SD) displayed (right). For each data type, radial uniformity was rejected for both groups (Rayleigh uniformity test p<0.0001) and there were no significant differences in circular mean between them (Watson’s two-sample test for homogeneity, p>0.1). (**D**) Linear mixed modelling results for oral temperature. Solid blue lines represent model fits, shaded areas and double-ended error bars represent 95% confidence intervals, dark grey circles display residuals (single temperature data points), and smoothed dashed yellow lines represent partial residuals. The x-axis for Time summarizes the continuous variable of time distance since the participant’s MSF_sc_ (proportion of a linearized unit circle, where 0=MSF_sc_ and 1=24 hours). Note time-dependent trend but lack of significant diurnal variation in oral temperature, likely reflecting inherent practical challenges of obtaining accurate oral temperature readings in human subjects.^34^

All subjects exhibited diurnal variation in wrist skin temperature, which was anti-phasic with their rhythm in activity and light exposure, in the week preceding their scans (Fig.1B-C and fig.S1-S2). BMI was marginally higher in males (P=0.014; Table 2). Oral temperature was 0.29°C higher in luteal females relative to males (95% confidence interval 0.03 to 0.58, P=0.029), and 0.04°C higher for a unit increase in BMI (0.005 to 0.083, P=0.024; Fig.1D). There were no differences in oral temperature by age or time of day however, despite daily changes in environmental temperature (Fig.1D, fig.S3). Brain locations for MRS data sampling are shown in Fig.2A. MRS data from one female were excluded due to a HRF; 24 *T*_Br_ data points from a total of 9434 (0.25%) were excluded because they did not meet quality control criteria for MRS spectral fitting (Supplementary Methods, fig.S4, Table S3). The data points that failed quality control derived from 15 of the 40 subjects scanned. Together, these data confirmed that our cohort was representative of healthy adult men and women with respect to basic physiological parameters, chronotype distribution, and sleep patterns. Our novel chronotype-controlled imaging protocol reproducibly obtains time-resolved *T*_Br_ data at high spatial resolution.

**Table 2.** Healthy participant demographics and sleep characteristics. Data presented as arithmetic mean (SD) except for Sleep Onset, Sleep Offset, and Acrophase (where circular mean (SD) is presented) and ‘slept during scan’ where numbers of individuals are presented as definite (possibly). Mean calculated across entire data collection period for each participant prior to calculation of group mean, where applicable. ^†^BMI higher in males than females (P=0.014; unpaired two-tailed t-test with Welch’s correction). Sleep onset = bed time plus latency of sleep onset. Sleep offset = wake up time. Sleep duration = duration between sleep onset and offset. Total sleep time = total duration of sleep period after removing periods of wakefulness. WASO = wake after sleep onset time (refers to periods of wakefulness occurring after defined sleep onset; a reflection of sleep fragmentation). Sleep efficiency = the percentage of time spent asleep while in bed. Calculated by dividing the amount of time spent asleep by the total amount of time in bed. A normal sleep efficiency is considered to be 80% or higher. MSF_sc_ = sleep corrected midpoint of sleep on free days (sleep onset on free days plus half of the average weekly sleep duration for all days). MSW_sc_ = sleep corrected midpoint of sleep on work days (sleep onset on work days plus half of the average weekly sleep duration for all days). PCSM = previous corrected sleep midpoint (sleep corrected midpoint of sleep on the night before scanning). SJL_sc_ = sleep corrected social jetlag (MSF_sc_-MSW_sc_ or absolute difference between sleep onset on free and work days when average sleep duration was longer on free than work days; if average sleep duration was longer on work days than free days, SJL_sc_ was calculated as the absolute difference between sleep offset on free and work days). Note that this parameter was calculated only for participants that reported at least one of each ‘day type’ (free or scheduled) during data collection. CFI = circadian function index; this parameter ranged from 0.43– 0.73 in an age-matched group of healthy volunteers.^35^

|  | **Females (n=20)** | **Males (n=20)** |
| --- | --- | --- |
| **Age (y)** | 29.76 (5.48) | 31.81 (6.16) |
| **BMI** | 22.33 (2.80) | 24.97 (3.65)<sup>†</sup> |
| **No. days actigraphy data** |  |  |
| **Free** | 1.6 (1.23) | 1.65 (1.50) |
| **Scheduled** | 6.0 (1.26) | 5.95 (1.73) |
| **Total** | 7.6 (0.60) | 7.6 (0.94) |
| **Sleep onset** | 23:33 (00:55) | 23:59 (01:07) |
| **Onset latency (min)** | 5.4 (4.26) | 4.4 (2.33) |
| **Sleep offset** | 07:40 (00:50) | 07:50 (01:04) |
| **Sleep duration (min)** | 486.5 (33.47) | 474.2 (39.61) |
| **Total sleep time per night (min)** | 442.4 (34.95) | 424.8 (37.00) |
| **WASO (min)** | 40.05 (17.90) | 43.80 (22.07) |
| **Sleep efficiency (%)** | 89.93 (3.86) | 89.04 (4.99) |
| **MSF<sub>sc</sub>** | 03:56 (01:01) | 03:58 (01:26) |
| **MSW<sub>sc</sub>** | 03:33 (00:50) | 03:53 (01:01) |
| **PCSM** | 03:15 (00:34) | 03:31 (01:17) |
| **SJL<sub>sc</sub> (min)** | 52.27 (49.28) | 38.82 (34.06) |
| **Acrophase** | 15:09 (01:24) | 15:22 (01:22) |
| **CFI** | 0.65 (0.07) | 0.67 (0.08) |
| **Oral temperature (°C)** |  |  |
| **Morning** | 36.18 (0.51) | 36.02 (0.40) |
| **Afternoon** | 36.11 (0.60) | 36.03 (0.48) |
| **Evening** | 36.09 (0.57) | 35.84 (0.43) |
| **MRI room temperature (°C)** |  |  |
| **Morning** | 21.02 (0.67) | 21.36 (0.76) |
| **Afternoon** | 21.98 (0.63) | 21.94 (0.71) |
| **Evening** | 21.30 (0.64) | 21.38 (0.53) |
| **Scan duration (minutes)** |  |  |
| **Morning** | 31.80 (3.82) | 31.10 (3.29) |
| **Afternoon** | 30.50 (6.68) | 29.61 (2.97) |
| **Evening** | 29.55 (2.09) | 28.63 (1.64) |
| **Slept during scan** |  |  |
| **Morning** | 1 (0) | 2 (3) |
| **Afternoon** | 6 (1) | 6 (2) |
| **Evening** | 5 (0) | 5 (1) |

**Fig. 2.**
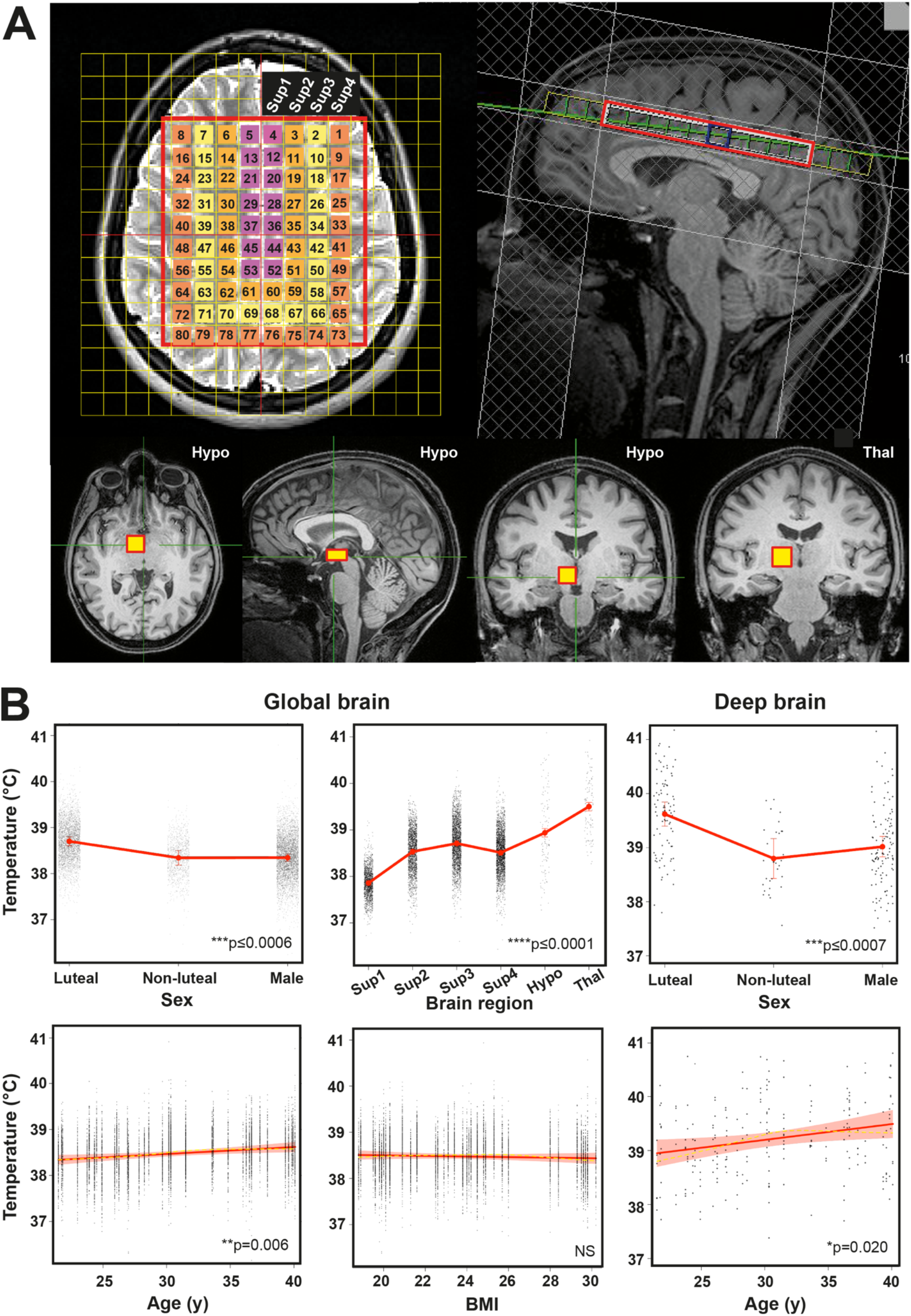
Human *T*_Br_ is spatially heterogenous. (**A**) Representative annotated MR images to show MRS extraction protocol immediately after whole-brain structural acquisition. T2-weighted axial (top left) and T1-weighted mid-sagittal (top right) image showing multivoxel MRS overlay for more superficial brain regions including cerebral grey and white matter; note positioning superior to corpus callosum. From this multivoxel acquisition, MRS data was extracted from each of the numbered voxels individually; for the final statistical model, the whole cerebral region was split into 4 superficial groups of voxels (Sup1–4, depicted as separate colours in the overlay, from medial to lateral). T1-weighted axial, sagittal, and coronal images (bottom three images from left side, respectively) showing orthogonal positioning of single voxel in right hypothalamus (yellow box). T1-weighted coronal image (bottom right) showing positioning of single MRS voxel in right thalamus (yellow box). (**B**) Linear mixed modelling results for global *T*_Br_ by sex, age, brain region, and BMI, and for deep *T*_Br_ (including thalamus and hypothalamus) by sex and age. Solid red lines represent model fits, shaded areas and double-ended error bars represent 95% confidence intervals, dark grey circles display residuals (single temperature data points), and smoothed dashed yellow lines represent partial residuals. For sex, P-value reflects comparisons of each group with luteal females. For brain region, P-value represents comparisons of each region relative to superficial region 1 (parasagittal group of voxels). Sup1–4, superficial brain regions 1–4 from medial to lateral; Hypo, hypothalamus; Thal, thalamus.

### Regional variation in brain temperature by sex and age

Global *T*_Br_ was higher than oral temperature (38.5 SD 0.4°C versus 36.0 SD 0.5°C), and was 0.36°C higher in luteal females relative to follicular females and males (95% confidence interval 0.17 to 0.55, P=0.0006 and 0.23 to 0.49, P<0.0001, respectively). This sex difference appeared to be driven by menstrual cycle phase (fig.S5). Despite age-selective recruitment, we captured an age-dependent increase in *T*_Br_, most notably in deep brain regions (thalamus and hypothalamus; 0.6°C over 20 years; 0.11 to 1.07; P=0.0002). Sex, age, and spatial effects on *T*_Br_ are summarized in Fig.2B and Fig.S6A. The mean maximal spatial *T*_Br_ range (difference between hottest and coolest voxel in an individual at any given time point) was 2.41 SD 0.46°C. In the cerebrum, white matter-predominating areas were relatively warm. The lowest temperatures were observed in cortical grey matter regions lying close to the brain surface and adjacent to a major venous drainage channel (region Sup1, surrounding the superior sagittal sinus). The highest temperatures were observed in the thalamus (1.64°C higher than cortical grey matter, 1.57 to 1.72, P<0.0001; 0.56°C higher than hypothalamus, 0.39 to 0.73, P<0.0001). Eight female and 12 male participants reported having ‘definitely’ or ‘possibly’ fallen asleep during one or more scans; this had no measurable impact on *T*_Br_ within the 30-minute scan time (Appendix 5). Collectively, these data show that normal human *T*_Br_ substantially exceeds oral temperature and varies by sex, age, menstrual cycle, and brain region.

### Diurnal variation in brain temperature

Absolute *T*_Br_ is ultimately determined by a balance between the rate of heat generated by the brain, and its rate of heat loss, mediated principally by CBF.^36,37^ Since blood arrives to the brain from the body at a lower temperature, this temperature gradient should enable effective brain heat removal, as long as cerebral perfusion is maintained.^38^ It follows that *T*_Br_ must be partially determined by *T*_Bo_. Since *T*_Bo_ and CBF both show clear diurnal regulation in humans, with lower temperature and higher CBF at night,^20,21^ we reasoned that human *T*_Br_ should drop in the evening. Our linear mixed model (Fig.3A-B) revealed that global *T*_Br_ varied by 0.57°C (95% confidence interval 0.40 to 0.75, P<0.0001) across time; whereas deep brain varied by 0.86°C (0.37 to 1.26, P=0.0001) and the hypothalamus displayed the greatest temporal variation (1.21 SD 0.65°C, range 0.27 to 2.75°C). Diurnal temperature variation was significantly greater in deep brain regions than in the cerebrum or the body (oral temperature; Fig.3C and fig.S6B-C), and for all brain regions, *T*_Br_ was lowest in the late evening. Robust, approximately sinusoidal, daily *T*_Bo_ rhythms are a very well-characterised aspect of human physiology, and similar temperature rhythms have been extensively documented in other diurnal mammals in the brain and body.^39,40^ Since *T*_Br_ is expected to depend (at least in part) on *T*_Bo_, we used the simplest and most appropriate mathematical model (cosinor analysis) to predict diurnal human *T*_Br_ in a continuous fashion. We interpolated a sinusoidal time series for *T*_Br_ in six brain regions of interest (Fig.3D). The predicted average minimum (anticipated around MSF_sc_, ∼3am) was 38.4°C in luteal females and 38.0°C in males. Importantly, the diurnal *T*_Br_ range across individuals was ∼37.0–40.3°C in healthy cortical white matter—the location measured in patients with moderate-to-severe brain injury. In summary, these data show that normal human *T*_Br_ varies substantially over the day, in a sex- and brain region-dependent fashion.

**Fig. 3.**
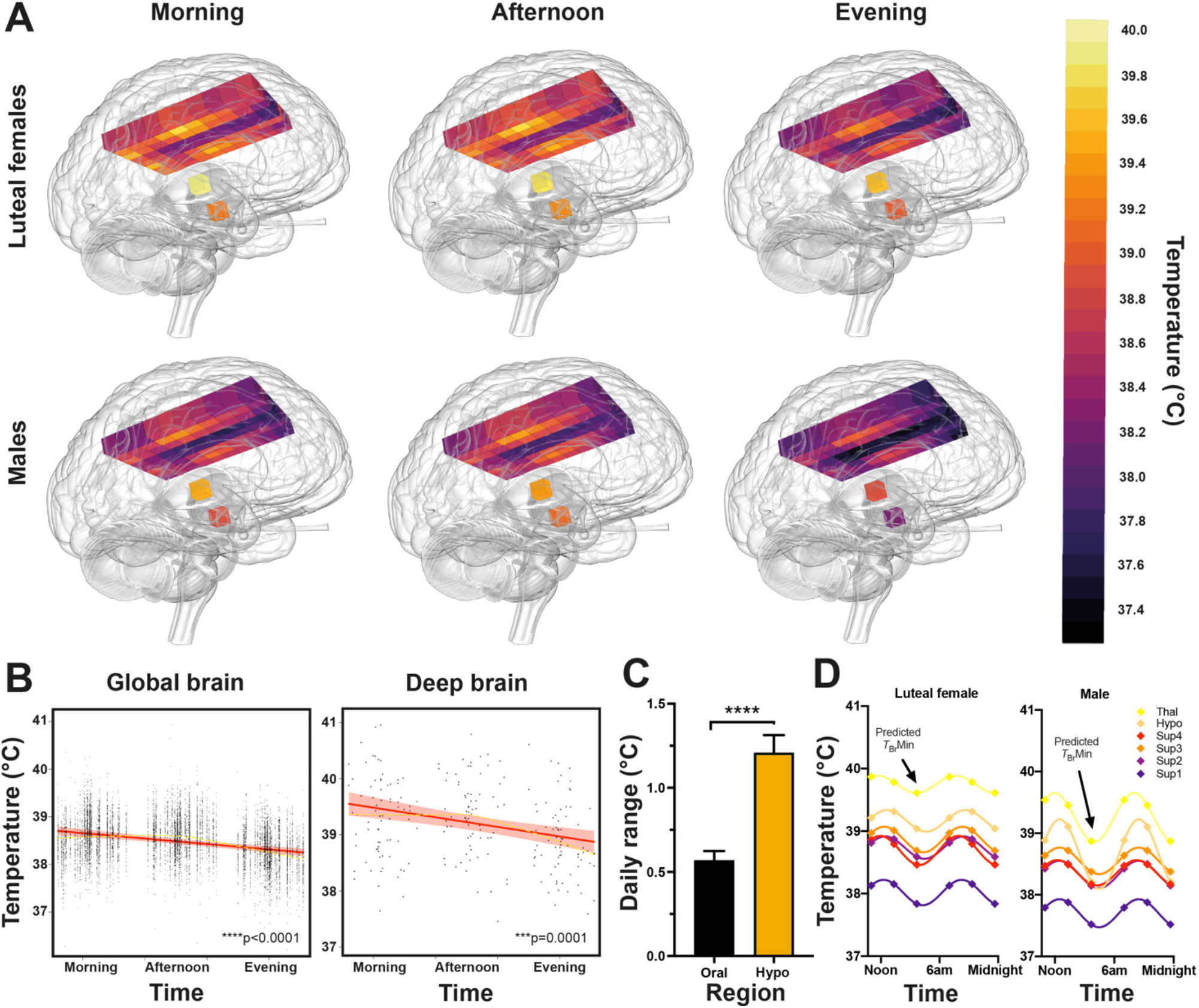
Human *T*_Br_ varies by time of day. (**A**) Snapshot 3D maps of *T*_Br_ at each data collection point. Inferno colour scale is used to assign a temperature to each tissue voxel, to 0.1°C resolution. Aggregate temperatures are displayed in each voxel for luteal females (n=14) and males (n=20) separately. (**B**) Linear mixed modelling results for *T*_Br_ by time of day; results for global *T*_Br_ (left) and deep brain *T*_Br_ (thalamus and hypothalamus, right) are shown. Solid red lines represent model fits, shaded areas represent 95% confidence intervals, dark grey circles display residuals (single temperature data points), and smoothed dashed yellow lines represent partial residuals. The x-axis for Time summarizes the continuous variable of time distance since the participant’s MSF_sc_ (proportion of a linearized unit circle, where 0=MSF_sc_ and 1=24 hours). (**C**) Temperature range (maximum versus minimum across three tested time points) for oral and hypothalamic sites for each healthy participant (n=39). Temperature varied more by time of day in the hypothalamus than orally (repeated measures one-way ANOVA with Sidak’s multiple comparisons test ****P<0.0001; see fig.S6B for other brain regions). (**D**) 24-hour temperature rhythms of the healthy brain, double plotted. Interpolated average *T*_Br_ rhythms in healthy luteal females (n=14) and males (n=20) based on sinusoidal fit using temperatures measured at three time points. Note higher temperatures in all regions in luteal females relative to males and marked variation in deep brain temperatures in males. Arrows point to predicted *T*_Br_ minima around 2–3am (approaching MSF_sc_). Sup1–4, superficial brain regions 1–4 from medial to lateral; Hypo, hypothalamus; Thal, thalamus.

### HEATWAVE—a 4D map of human brain temperature

Combining our spatial and temporal observations, we built HEATWAVE—a 4-dimensional map to model human *T*_Br_ at hourly resolution (Movies S1 and S2). HEATWAVE can be dynamically explored at (https://www2.mrc-lmb.cam.ac.uk/groups/oneill/research/heatwave/). These comparisons highlight the relatively hot deep brain regions and their greater diurnal variation in males than females. The HEATWAVE videos complement the voxel maps in Fig.3A, which represent a reference resource for interpreting human *T*_Br_ at each of the time points tested. Since each data point in each map is an average of data from multiple individuals, it incorporates the range of ages, BMIs, and chronotypes expected for each sex in the demographic tested. Our data collection points also cater for the times (morning and afternoon) when most patients would present for MR-based neuroimaging in the non-acute setting. In addition to modelling diurnal human *T*_Br_ in a continuous fashion, HEATWAVE thus provides the first comprehensive spatially-resolved description of normal human *T*_Br_ at three clinically-relevant time points; a rich reference dataset for future studies in different age groups and patient cohorts.

### Daily brain temperature rhythms predict patient survival

The characterisation of physiological *T*_Br_ variation in humans allows its dysregulation by brain injury to be understood in context for the first time. Of 134 eligible patient records screened, 114 had at least 24h of temperature data recorded (criteria level A). Of these, 110 patients had sufficient temperature data (≥ 36h) for diurnal rhythm analysis (criteria levels A and B; for eight of these patients, sufficient data was available for *T*_Bo_ only). Outcome in intensive care was available for 113/114 patients (criteria levels A and C), and a complete set of injury severity scores (PLR, GCS, and GCSM) was available for 109/114 patients (criteria levels A, C, D). A total of 107 patients met all criteria levels, and 100 patients had sufficient data to test for an association between diurnal *T*_Br_ rhythmicity and outcome (mortality). Summary data are shown in Table 3. As in our healthy cohort, mean *T*_Br_ (38.5 SD 0.8°C) was significantly higher than mean *T*_Bo_ (37.5 SD 0.5°C; P<0.0001, Fig.4A), but the range was much wider (32.6 to 42.3°C). *T*_Br_ was not affected by the site of intracranial probe placement relative to focal injury (fig.S7). We found an approximately daily temperature rhythm in 25/110 patients, of which 23 had a daily *T*_Br_ rhythm (Fig.4B, fig.S8). However, across the cohort, the timings of temperature maxima and minima were poorly aligned with the external day-night cycle. This desynchronization of internal timing from the external solar cycle is a hallmark of circadian rhythms when external timing cues are diminished,^41^ and lies in stark contrast to rectal temperature data from healthy individuals maintained in the presence of daily light/dark and feed/fast cues (Fig.4C).

**Table 3.** TBI patient demographics and summary temperature data. Data presented as mean (SD); Min, minimum; Max, maximum; PLR, presence of pupillary light reflex in one or both eyes; GCS, Glasgow Coma Score; GCSM, Glasgow Coma Score Motor response. Individual patient temperature values and ranges were calculated from all available data present within the data analysis window that met inclusion/exclusion criteria (Table 1); aggregate results for males and females are presented here.

|  | **Females** | **Males** |
| --- | --- | --- |
| Age (y) | 55.1 (14.8) n=21 | 45.7 (17.2) n=94 |
| $T_{Br}$ Mean (°C) | 38.2 (0.85) n=20 | 38.6 (0.85) n=85 |
| $T_{Bo}$ Mean (°C) | 37.2 (0.52) n=17 | 37.5 (0.51) n=78 |
| $T_{Br}$ Range (°C) | 3.38 (1.21) n=20 | 3.21 (1.08) n=85 |
| $T_{Bo}$ Range (°C) | 2.65 (0.93) n=17 | 2.50 (0.87) n=85 |
| $T_{Br}$Max (°C) | 39.8 (1.00) n=20 | 40.3 (1.03) n=85 |
| $T_{Bo}$Max (°C) | 38.4 (0.70) n=17 | 38.8 (0.71) n=85 |
| $T_{Br}$Min (°C) | 36.4 (1.07) n=20 | 37.1 (0.99) n=85 |
| $T_{Bo}$Min (°C) | 35.7 (0.82) n=17 | 36.3 (0.76) n=85 |
| PLR | 1.86 (0.36) n=21 | 1.77 (0.59) n=86 |
| GCS | 7.90 (3.42) n=21 | 8.07 (3.60) n=87 |
| GCSM | 4.10 (1.61) n=21 | 3.71 (1.85) n=87 |

**Fig. 4.**
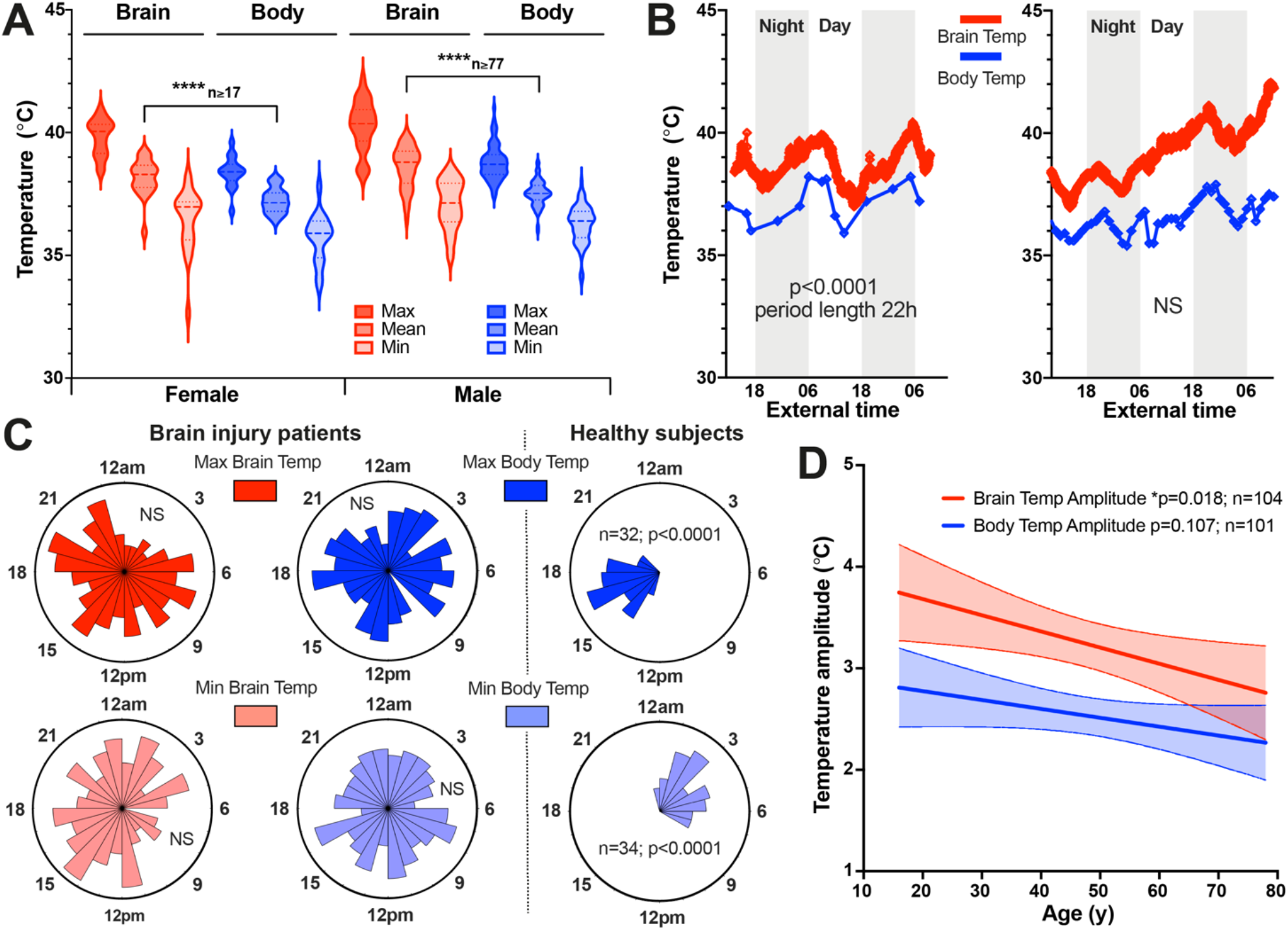
Temperature rhythms in TBI patients. (**A**) Violin plot of patient *T*_Br_ and *T*_Bo_ according to sex. Mean *T*_Br_ significantly greater than mean *T*_Bo_, mixed effects analysis with Tukey’s for multiple comparisons (****P<0.0001, females, n=20 for *T*_Br_ and n=17 for *T*_Bo_; males, n=85 for *T*_Br_ and n=77 for *T*_Bo_). (**B**) Representative raw data from 62 y female patient (left) showing daily variation in *T*_Br_ and *T*_Bo_, with *T*_Br_ consistently higher than *T*_Bo_ and both parameters in same phase. *T*_Br_ sampled once per minute; peak at 05:28 and nadir at 16:12 highlighting inversion of phase relationship with external day-night cycle under intensive care conditions (external time in 24-hour clock format). Representative raw data from 42 y male patient (right) showing lack of a daily rhythm in both *T*_Br_ and *T*_Bo_. (**C**) Rose plots (left) showing timings of temperature maxima and minima in 114 TBI patients (24-hour clock format). For all variables, the null hypothesis of a uniform distribution could not be rejected (Rayleigh test of uniformity; *T*_Br_Max, n=104, P=0.20; *T*_Br_Min, n=104, P=0.16; *T*_Bo_Max, n=101, P=0.99; *T*_Bo_Min, n=101, P=0.86). Contrast with healthy subject rectal temperature data from publicly-available database (right).^42^ (**D**) Linear regression of patient temperature ranges with age; reduction in temperature amplitude significant for brain (slope of -0.016 significantly different from zero; 95% confidence interval -0.029 to 0.003). Shaded areas represent 95% confidence intervals for lines of best fit. Max, maximum; Min, minimum; NS, not significant.

As for healthy adults, there was a relationship between *T*_Br_ and age; *T*_Br_ amplitude was reduced in older patients (P=0.018), dominated by an upward trend in minimum temperature (Fig.4D, fig.S9). Twenty-five patients died in intensive care. Applying a GLMM (Fig.5), we found that lack of a daily *T*_Br_ rhythm, or an age increase of 10 years, increased the odds of death in intensive care 12-fold and 11-fold, respectively (OR for death with rhythm 0.09; 95% confidence interval 0.01 to 0.84, P=0.035 and OR for death with ageing by 1 year 1.10; 1.05 to 1.16, P=0.0002). These relationships could not be explained by a general elevation in *T*_Br_, since mean *T*_Br_ was positively associated with survival (OR for death 0.45 for 1°C increase, 0.21 to 0.96, P=0.040). The presence of a diurnal *T*_Br_ rhythm did not correlate with either age or mean *T*_Br_ (Appendix 5). Together, these data show that daily temperature variation is frequently disrupted or absent in TBI patients and that *T*_Br_ variation is of greater prognostic use than absolute *T*_Br_. Older TBI patients lacking a daily *T*_Br_ rhythm are at greatest risk of death in intensive care, and presence of a daily *T*_Br_ rhythm appears to be the strongest single predictor of survival after TBI.^43^

**Fig. 5.**
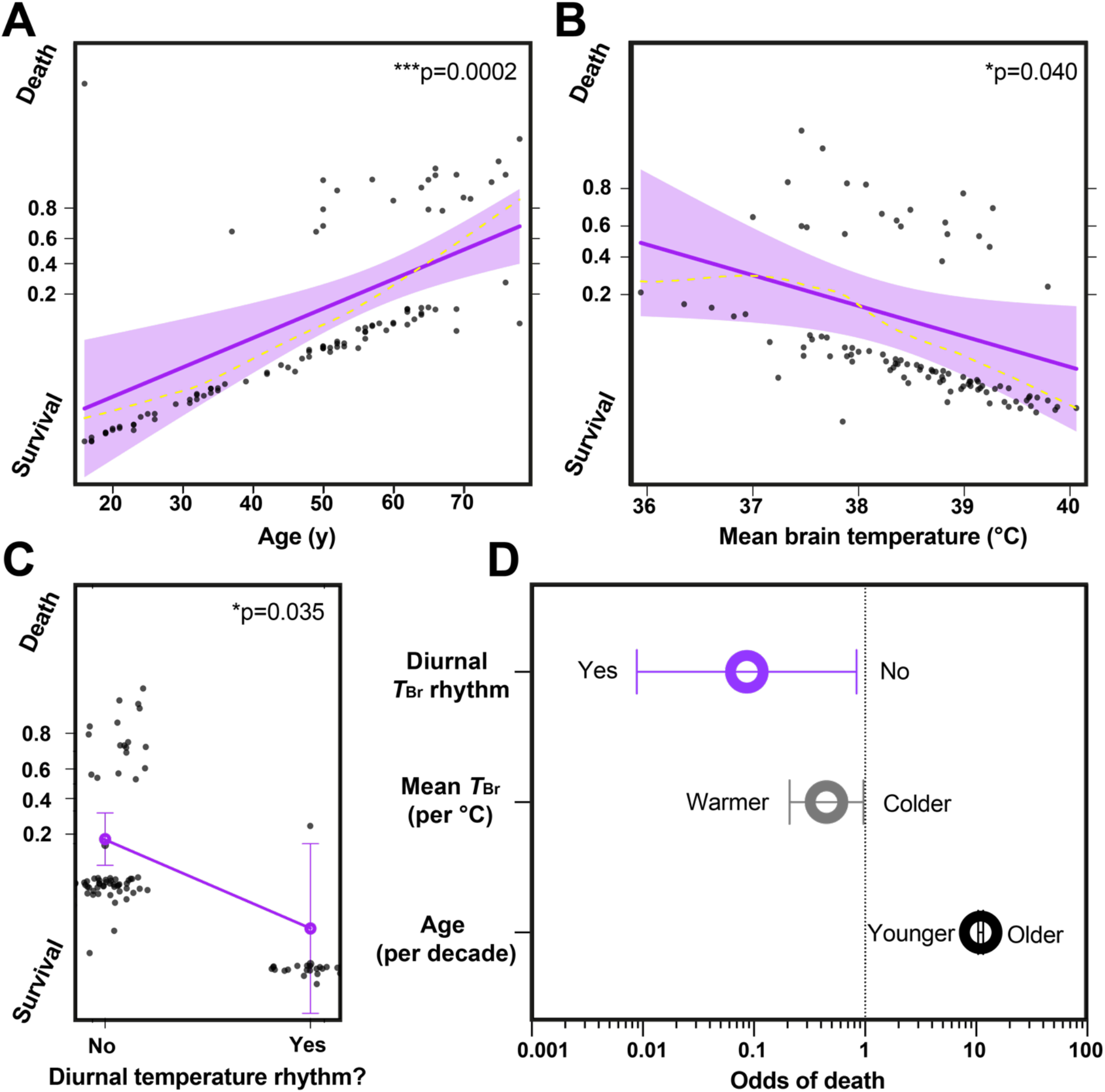
A daily *T*_Br_ rhythm predicts survival after brain injury. **(A-C)** GLMM results for outcome in n=100 TBI patients. Probability of death (‘success’ or ‘hit’ = 1) relative to survival (‘failure’ or ‘miss’ = 0) is depicted on the y-axis. Solid purple lines represent model fits for logit (log of the odds) binomial distribution for a given predictor and dark grey circles display residuals (individual patients). For numerical predictors **(A-B)**, shaded areas represent 95% confidence intervals and smoothed dashed yellow lines represent partial residuals. For the categorical predictor of presence/absence of diurnal temperature rhythm **(C)**, residuals are jittered in the x-axis direction for visibility and 95% confidence intervals are presented as double-ended error bars. **(D)** Odds of death in intensive care transformed from the data in **(A-C)**; the results for these three predictors are significant since the 95% confidence intervals (double-ended error bars) do not include 1. Note also that confidence intervals become numerically asymmetric once transformed from log odds to regular odds. Only factors that demonstrated a statistically significant relationship with mortality are shown. Note logarithmic scale on x-axis and large effect size for presence of a daily rhythm in *T*_Br_ in **(D)**. See Methods and Supplementary Methods for further details on the GLMM, and Appendix 5 for all numerical outputs and related code.

## DISCUSSION

### Principal findings

We have established a 4-dimensional map of human *T*_Br_, and shown how this parameter varies with time of day, brain region, age, and sex in healthy adults. These data provide clinicians with an urgently-needed and readily-accessible reference resource for evidence-based interpretation of *T*_Br_ data in patients. Furthermore, we have found a relationship between the presence of a daily *T*_Br_ rhythm and survival of TBI patients. Our findings demonstrate the high prognostic value of time-resolved *T*_Br_ measurements in neurocritical care, thus empowering a temperature-based prediction of mortality.^22^ Overall, this work reveals marked heterogeneity and dynamism of human *T*_Br_ that must influence neural cell activity, and represents an important correlate of brain health.

### Strengths and limitations

Although time-based human neuroimaging studies are sparse, some morning/afternoon comparisons are consistent with diurnal regulation of brain morphometry,^44,45^ as well as diurnal variation in neural activity and metabolism.^46-48^ However, prior studies were underpowered without consideration of chronotype and a late evening time point, which provides greater insight into healthy brain physiology by incorporating the period approaching habitual sleep time (Fig.3). It was neither practical nor clinically-relevant to deprive our participants of all external timing cues (to derive circadian *T*_Br_ variation), but this diurnal *T*_Br_ variation is almost identical to direct measurements obtained in healthy non-human primates under stringent conditions.^18^ Though unlikely, it is conceivable that gross regional differences in the activity of cellular water impact upon the apparent spatial *T*_Br_ variation we observe within an individual at a given time point. However, we are unaware of any supporting evidence for this, nor can it be attributed to simple grey versus white matter distribution.^49^ Moreover, such differences cannot influence the *T*_Br_ variation we have found in relation to time of day, sex, age, or menstrual cycle stage. This is illustrated well when we limit our model to a subset of deep brain regions of more homogenous tissue structure, where *T*_Br_ variation persists with respect to all of the aforementioned fixed effects (Fig.2B, 3B, fig.S6). Crucially, our robust statistical approach caters for multiple physiologically-relevant confounders within and between individuals that would have prevented the detection of significant *T*_Br_ variation in previous studies.^49,50^ Alongside the patient data (Fig.5), and multiple parallel methods of temperature measurement in healthy subjects by us and others,^22,51-53^ our results offer compelling evidence of a daily temperature rhythm throughout the normal human brain (Supplementary Text *‘Temperature rhythms and sleep’*).

The within-brain temperature gradient is remarkable (Fig.2B). As an ‘open’ thermodynamic system performing no mechanical work, aerobic metabolism of the brain releases heat at ∼0.66 J/min/g of tissue which is primarily removed by CBF.^38,54-56^ It is therefore highly likely that regional variation in neurovascular anatomy plays the chief role in creating spatial *T*_Br_ gradients (Supplementary Text *‘Temperature gradients’*). Although we cannot completely exclude a contribution from regional differences in water content,^49,57^ these are unlikely to explain the temperature difference between the thalamus and hypothalamus (both grey matter structures devoid of cerebrospinal fluid). We suggest that the lower temperature of the hypothalamus might reflect its closer proximity to major vascular networks such as the Circle of Willis. In principle, technical limitations (Supplementary Text *‘Technical limitations of brain thermometry’*) could potentially exaggerate MRS-derived temperature differences at the extreme edges of regions of interest (cerebral layer Sup4). The spatial distribution we have found is however very similar to non-human primates, excepting a larger gradient magnitude that is entirely consistent with the difference in brain volume between humans and rhesus monkeys.^19^ Unlike previous studies,^49,50^ we make no baseline assumption that temperature should be homogeneous across brain regions, nor between different tissue types within the brain. Importantly, we did not apply a post-acquisition correction to our MRS data to equalize temperatures between grey and white matter,^49,50^ since this would perpetuate the above assumption, and overlooks the clear tissue temperature differences observed in non-human primates and normothermic human patient brains.^14,16,19^ Indeed, higher temperatures in white matter-rich areas concur with predictions based on modelling perfusion, blood volume fraction, and heat generation in different brain tissues.^13,58-60^

### Possible mechanisms and implications

An increase in mean *T*_Br_ (Fig.2B) and a trend upwards in minimum *T*_Br_ (fig.S9) with age suggests that overnight brain cooling becomes less efficient in older people, leading to a damped *T*_Br_ rhythm. This age-dependent reduction in *T*_Br_ amplitude is consistent with studies of *T*_Bo_ and may contribute to the disrupted sleep patterns and ‘sundowning’ symptoms of dementia patients.^24,61-63^ Cerebral blood supply is considered so efficient that heat removal is achieved without the need for other mechanisms under most circumstances,^36,37,64^ which seems intuitive for the young, healthy brain. However, the vast literature linking neurodegeneration to cerebrovascular compromise indicates that our key brain cooling mechanism progressively deteriorates with age (Supplementary Text *‘Internal rhythms and health’*).^65,66^ Neuronal activity is highly sensitive to temperature change, with a Q_10_ of ∼2.3, although this is generally considered to be most problematic in the acute setting.^38,55,67^ In a study of 1130 epilepsy patients, 80–92% showed a 24-hour cycle of seizure rates, with events most common at ∼8am, when *T*_Br_ should increase most steeply (Fig.3D).^68^ Given that cooling can terminate epileptic discharges,^69^ diurnal changes in *T*_Br_ may well contribute to diurnal variation in the incidence of seizures and cluster headache.^70,71^

*T*_Bo_ increases, and its overnight drop is blunted, in luteal versus follicular phase women.^52,72^ This menstrual variation predicts that *T*_Bo_ is ∼0.4°C higher in the early luteal phase.^72^ For the first time, we report a parallel luteal-phase increase in global *T*_Br_ by ∼0.36°C, and of deep *T*_Br_ by 0.82°C (95% confidence interval 0.37 to 1.28, P=0.0006). This may contribute to the reported variable sleep patterns and changes in cognition across different stages of the menstrual cycle.^52,73^ A thermogenic effect of progesterone is well-recognized, and may involve direct stimulation of preoptic/anterior hypothalamic thermoregulatory neurons, or the suprachiasmatic nucleus.^52,53,74,75^ Despite its postulated neuroprotective effects, large clinical trials have failed to show any benefit of progesterone therapy for TBI—one reason for this could be damping of the daily *T*_Br_ rhythm (Fig.3D).^76^ It is widely accepted that BMI positively correlates with *T*_Bo_ as found here (Fig.1).^22,77^ Since BMI was slightly higher in males relative to females in our healthy cohort, a difference in BMI cannot explain the higher *T*_Br_ observed in luteal females and, notably, there was no relationship between BMI and *T*_Br_ overall (Fig.2B; Appendix 5). This supports our conclusion that *T*_Br_ cannot be solely dependent on, nor predicted from, *T*_Bo_ since brain heat removal also occurs through routes that are unaffected by adipose deposition.^36,37^

Essential to clinical diagnostics is the comparison of patient data with reference ranges from healthy individuals; MRS-thermometry now makes this possible for *T*_Br_. We have validated our core MRS findings using multiple complementary methods of temperature measurement. This is most pressing for *T*_Br_, where such methods are effectively mutually exclusive in healthy individuals and neurocritical care patients. TTM is the mainstay of neuroprotection subsequent to out-of-hospital cardiac arrest.^78^ Here, the objective is to reduce *T*_Br_, which is rarely measured directly in trials that test the therapeutic value of TTM in the context of brain injury. Cooling adults at the ‘wrong’ biological time or fixing patient temperatures at a constant target value may further compromise thermoregulation by abolishing physiologically-important, health-critical temperature variation. The highest temperature we observed in any healthy individual was 40.9°C in the thalamus of a luteal female in the afternoon; whilst the perception exists that a *T*_Br_ of this value would cause brain damage, there is no direct evidence for this, and similar deep brain temperatures are observed physiologically in other mammalian species.^79^ Furthermore, the *T*_Br_ range in our volunteers raises doubt over whether *T*_Br_ was abnormally high in some patient reports.^15^ Current temperature management guidelines do not consider physiological differences by sex or time of day,^80^ and whether adults should be cooled at all in neurocritical care remains controversial. A clear understanding of how and why *T*_Br_ varies in health and disease is thus imperative. Here we report a healthy cortical white matter maximum *T*_Br_ of 40.3°C, but we caution strongly against overinterpreting single *T*_Br_ values or transitory trends. Rather, we recognize the need for technological solutions that allow individualized target temperature ranges to be determined, facilitating decision making that incorporates chronotype, age, sex, menstrual cycle, and time of day.

### Unanswered questions and future research

Prospective controlled trials are needed to confirm the predictive power of *T*_Br_ rhythmicity in relation to patient outcome, as well as the clinical utility of TTM protocols in brain-injured patients. There may be high clinical value in exploiting *T*_Br_ variation to detect or monitor focal pathologic processes such as neoplasia, trauma, vascular insults, and epileptogenesis, but also more distributed inflammatory, metabolic, and neuropsychiatric diseases.^54,81-87^ In particular, future work should be directed to address whether abnormal *T*_Br_ rhythmicity may serve as an early biomarker of neurodegeneration; a mechanistically opaque process for which early diagnostics are notably deficient.^26,65^ Beyond brain injury and disease (Supplementary Text *‘Clinical applications’*), our results further question the value of single-point temperature measurements using peripheral thermometers.^88^ We have shown that more sophisticated analyses can better exploit temperature as a clinical tool. Wearable devices now permit easy and convenient recording of daily rhythms in many physiological parameters. Algorithm-based temperature profiling will help accomplish the goals of precision medicine, not just for individuals,^89^ but at scale. For example, in an infectious disease outbreak, real-time screening for fever development could rapidly identify high-risk individuals by deviation from their own temperature rhythm, rather than a population ‘mean’ or by random testing. Personalized, digital, round-the-clock temperature monitoring would thus advance remote health tracking and evidence-based enforcement of global health policy in the context of emerging disease. Whilst providing excellent spatial resolution, MRS brain thermometry is clearly impractical for routine use in most clinical settings. Since core *T*_Bo_ is not a faithful proxy for *T*_Br,_^90^ our work highlights an urgent need for cost-effective, non-invasive technologies that can capture longitudinal variations in *T*_Br_, alongside core body and peripheral temperatures.

## Data Availability

Data sharing: Individual patient data contained within the CENTER-TBI database are not publicly available but permissions for access can be requested at https://www.center-tbi.eu/data. We are committed to sharing all other anonymised individual participant and patient data that would support the clinical community. All shareable items are available immediately upon publication and indefinitely, or ending 5 years following article publication, by reasonable request from the Lead Author at. Shareable items will be available to anyone who wishes to access them and for any purpose. Code for statistical modelling is provided in Supplementary Appendix 5.

## Acknowledgements

We gratefully acknowledge CiBraT Study Radiographers at the Edinburgh Imaging (Royal Infirmary of Edinburgh) Facility (Gayle Barclay, Annette Cooper, Lucy Kesseler, Donna McIntyre, Isla Mitchell, and Maddy Murphy) and CiBraT Study Participants; administrative support from Dawn Cardy, Joanne Douglas, and Duncan Martin; data management support from Dominic Job and Aidan Hutchison; governance support from the Academic and Clinical Central Office for Research and Development (ACCORD); recruitment support from Edinburgh Imaging, Edinburgh Neuroscience, and the Centre for Clinical Brain Sciences; graphics support from Lesley McKeane and Matthew Fry (MRC Laboratory of Molecular Biology Visual Aids); writing support from Ramanujan Hegde (MRC Laboratory of Molecular Biology), and valuable discussion from Rachel Edgar (Imperial College London) and O’Neill Lab members. CENTER-TBI data extraction was supported by Erta Beqiri, University of Milan.

## Contributors

NMR conceived the idea for the work, designed and orchestrated the study, conducted recruitment and data collection for the prospective study, analyzed and interpreted all data, prepared all tables and figures, and wrote and revised the manuscript. JSON made conceptual contributions to the prospective study and contributions to the overall study design, interpreted data, contributed to figure preparation, and revised the manuscript. IM and MJT collected, analyzed, and interpreted spectroscopy data, contributed to figure preparation, and revised the manuscript. FMC contributed to study design, devised the statistical analysis plan for the prospective study, and revised the manuscript. AE and MC contributed to study design, CENTER-TBI High Resolution ICU data curation and extraction, statistical analysis, and revised the manuscript. GM analyzed, interpreted, and reported on prospective structural MRI data, and revised the manuscript. JR collected and developed extraction methods for retrospective patient data, contributed to data analysis, and revised the manuscript. All authors contributed to the literature search. JR, IM, and JSON contributed equally to this work (are joint last authors). NMR and JSON are guarantors and joint corresponding authors; NMR and JSON both attest that all listed authors meet authorship criteria and that no others meeting the criteria have been omitted. Investigators for The CENTER-TBI High HR ICU Sub-Study include: Audny Anke, Department of Physical Medicine and Rehabilitation, University hospital Northern Norway; Ronny Beer, Department of Neurology, Neurological Intensive Care Unit, Medical University of Innsbruck, Innsbruck, Austria; Bo-Michael Bellander, Department of Neurosurgery & Anesthesia & intensive care medicine, Karolinska University Hospital, Stockholm, Sweden; Andras Buki, Department of Neurosurgery, University of Pecs and MTA-PTE Clinical Neuroscience MR Research Group and Janos Szentagothai Research Centre, University of Pecs, Hungarian Brain Research Program, Pecs, Hungary; Giorgio Chevallard, NeuroIntensive Care, Niguarda Hospital, Milan, Italy; Arturo Chieregato, NeuroIntensive Care, Niguarda Hospital, Milan, Italy; Giuseppe Citerio, NeuroIntensive Care Unit, Department of Anesthesia & Intensive Care, ASST di Monza, Monza, Italy; and School of Medicine and Surgery, Università Milano Bicocca, Milano, Italy; Endre Czeiter, Department of Neurosurgery, University of Pecs and MTA-PTE Clinical Neuroscience MR Research Group and Janos Szentagothai Research Centre, University of Pecs, Hungarian Brain Research Program (Grant No. KTIA 13 NAP-A-II/8), Pecs, Hungary; Bart Depreitere, Department of Neurosurgery, University Hospitals Leuven, Leuven, Belgium; George Eapen, Shirin Frisvold, Department of Anesthesiology and Intensive Care, University Hospital Northern Norway, Tromso, Norway; Raimund Helbok, Department of Neurology, Neurological Intensive Care Unit, Medical University of Innsbruck, Innsbruck, Austria; Stefan Jankowski, Neurointensive Care, Sheffield Teaching Hospitals NHS Foundation Trust, Sheffield, UK; Daniel Kondziella, Departments of Neurology, Clinical Neurophysiology and Neuroanesthesiology, Region Hovedstaden Rigshospitalet, Copenhagen, Denmark; Lars-Owe Koskinen, Department of Clinical Neuroscience, Neurosurgery, Umea University Hospital, Umea, Sweden; Geert Meyfroidt, Intensive Care Medicine, University Hospitals Leuven, Leuven, Belgium; Kirsten Moeller, Department Neuroanesthesiology, Region Hovedstaden Rigshospitalet, Copenhagen, Denmark; David Nelson, Department of Neurosurgery & Anesthesia & intensive care medicine, Karolinska University Hospital, Stockholm, Sweden; Anna Piippo-Karjalainen, Helsinki University Central Hospital, Helsinki, Finland; Andreea Radoi, Department of Neurosurgery, Vall d’Hebron University Hospital, Barcelona, Spain; Arminas Ragauskas, Department of Neurosurgery, Kaunas University of technology and Vilnius University, Vilnius, Lithuania; Rahul Raj, Helsinki University Central Hospital, Helsinki, Finland; Jonathan Rhodes, Department of Anaesthesia, Critical Care & Pain Medicine NHS Lothian & University of Edinburgh, Edinburgh, UK; Saulius Rocka, Department of Neurosurgery, Kaunas University of technology and Vilnius University, Vilnius, Lithuania; Rolf Rossaint, Department of Anaesthesiology, University Hospital of Aachen, Aachen, Germany; Juan Sahuquillo, Department of Neurosurgery, Vall d’Hebron University Hospital, Barcelona, Spain; Oliver Sakowitz, Klinik für Neurochirurgie, Klinikum Ludwigsburg, Ludwigsburg, Germany; Department of Neurosurgery, University Hospital Heidelberg, Heidelberg, Germany; Ana Stevanovic, Department of Anaesthesiology, University Hospital of Aachen, Aachen, Germany; Nina Sundström, Department of Radiation Sciences, Biomedical Engineering, Umea University Hospital, Umea, Sweden; Riikka Takala, Perioperative Services, Intensive Care Medicine, and Pain Management, Turku University Central Hospital and University of Turku, Turku, Finland; Tomas Tamosuitis, Neuro-intensive Care Unit, Kaunas University of Health Sciences, Kaunas, Lithuania; Olli Tenovuo, Rehabilitation and Brain Trauma, Turku University Central Hospital and University of Turku, Turku, Finland; Peter Vajkoczy, Neurologie, Neurochirurgie und Psychiatrie, Charité–Universitätsmedizin Berlin, Berlin, Germany; Alessia Vargiolu, NeuroIntensive Care Unit, Department of Anesthesia & Intensive Care, ASST di Monza, Monza, Italy; Rimantas Vilcinis, Department of Neurosurgery, Kaunas University of Health Sciences, Kaunas, Lithuania; Stefan Wolf, Interdisciplinary Neuro Intensive Care Unit, Charité– Universitätsmedizin Berlin, Berlin, Germany; Alexander Younsi, Department of Neurosurgery, University Hospital Heidelberg, Heidelberg, Germany.

## Funding

The prospective CiBraT Study was funded by a Medical Research Council Clinician Scientist Fellowship awarded to NMR (MR/S022023/1). JSON is supported by the Medical Research Council (MC_UP_1201/4). Some of the data contributing to the retrospective analysis were obtained in the context of CENTER-TBI, a large collaborative project (EC grant 602150) supported by the European Union’s 7th Framework program (FP7/2007-2013). Additional funding for patient data collection was obtained from the Hannelore Kohl Stiftung (Germany), from OneMind (USA) and from Integra LifeSciences Corporation (USA). MJT acknowledges funding from the NHS Lothian Research and Development Office. This research was independent from funders. Funders had no role in the study design; in the collection, analysis, and interpretation of data; in the writing of the report; or in the decision to submit the article for publication.

## Competing interests

all authors have completed the Unified Competing Interest form (available on request from the corresponding author) and declare: no support from any organisation for the submitted work; no financial relationships with any organisations that might have an interest in the submitted work in the previous three years, no other relationships or activities that could appear to have influenced the submitted work.

## Ethical approval

The prospective study was co-sponsored by the University of Edinburgh and NHS Lothian (R&D Project Number 2019/0133). Ethics approval was obtained from the Academic and clinical office for research support (ACCORD) Medical Research Ethics Committee (AMREC; Study Number 18-HV-045). All participants provided written informed consent to participate. For TBI patient data analysis, approval was provided by NHS Scotland (14/SS/1086, R&D Department, University Hospitals Division NHS Lothian 2015/0171) for data collected at the Intensive Care Unit, Western General Hospital, Edinburgh, UK. For data extracted from other clinical sites via the CENTER-TBI database, the CENTER-TBI study was conducted in accordance with all relevant laws of the EU if directly applicable or of direct effect and all relevant laws of the country where the Recruiting sites were located, including but not limited to, the relevant privacy and data protection laws and regulations (the “Privacy Law”), the relevant laws and regulations on the use of human materials, and all relevant guidance relating to clinical studies from time to time in force including, but not limited to, the ICH Harmonised Tripartite Guideline for Good Clinical Practice (CPMP/ICH/135/95) (“ICH GCP”) and the World Medical Association Declaration of Helsinki entitled “Ethical Principles for Medical Research Involving Human Subjects”. Informed Consent by the patients and/or the legal representative/next of kin was obtained, accordingly to the local legislations, for all patients recruited in the Core Dataset of CENTER-TBI and documented in the e-CRF. Ethical approval was obtained for each recruiting site. The list of sites, Ethical Committees, approval numbers and approval dates can be found on the website: https://www.center-tbi.eu/project/ethical-approval.

## Data sharing

Individual patient data contained within the CENTER-TBI database are not publicly available but permissions for access can be requested at https://www.center-tbi.eu/data. We are committed to sharing all other anonymised individual participant and patient data that would support the clinical community. All shareable items are available immediately upon publication and indefinitely, or ending 5 years following article publication, by reasonable request from the Lead Author at. Shareable items will be available to anyone who wishes to access them and for any purpose. Code for statistical modelling is provided in Supplementary Appendix 5.

## Transparency

The lead author (NMR) affirms that the manuscript is an honest, accurate, and transparent account of the study being reported; that no important aspects of the study have been omitted; and that any discrepancies from the study as planned have been explained.

## Copyright

The Corresponding Author has the right to grant on behalf of all authors and does grant on behalf of all authors, an exclusive licence (or non-exclusive for government employees) on a worldwide basis to the BMJ Publishing Group Ltd to permit this article (if accepted) to be published in BMJ editions and any other BMJPGL products and sublicences such use and exploit all subsidiary rights, as set out in our licence.

## Notes

### Competing Interest Statement

The authors have declared no competing interest.

### Clinical Trial

UK CRN NIHR CPMS 42644; ClinicalTrials.gov number, NCT02210221

### Author Declarations

Ethical approval: The prospective study was co-sponsored by the University of Edinburgh and NHS Lothian (R&D Project Number 2019/0133). Ethics approval was obtained from the Academic and clinical office for research support (ACCORD) Medical Research Ethics Committee (AMREC; Study Number 18-HV-045). All participants provided written informed consent to participate. For TBI patient data analysis, approval was provided by NHS Scotland (14/SS/1086, R&D Department, University Hospitals Division NHS Lothian 2015/0171) for data collected at the Intensive Care Unit, Western General Hospital, Edinburgh, UK. For data extracted from other clinical sites via the CENTER-TBI database, the CENTER-TBI study was conducted in accordance with all relevant laws of the EU if directly applicable or of direct effect and all relevant laws of the country where the Recruiting sites were located, including but not limited to, the relevant privacy and data protection laws and regulations (the 'Privacy Law'), the relevant laws and regulations on the use of human materials, and all relevant guidance relating to clinical studies from time to time in force including, but not limited to, the ICH Harmonised Tripartite Guideline for Good Clinical Practice (CPMP/ICH/135/95) ('ICH GCP') and the World Medical Association Declaration of Helsinki entitled 'Ethical Principles for Medical Research Involving Human Subjects'. Informed Consent by the patients and/or the legal representative/next of kin was obtained, accordingly to the local legislations, for all patients recruited in the Core Dataset of CENTER-TBI and documented in the e-CRF. Ethical approval was obtained for each recruiting site. The list of sites, Ethical Committees, approval numbers and approval dates can be found on the website: https://www.center-tbi.eu/project/ethical-approval.

## REFERENCES

1 Grodzinsky E, Levander MS. History of the thermometer. Understanding Fever and Body Temperature 2019;23–25. doi: 10.1007/978-3-030-21886-7_3

2 Phillips MA, Burrows JN, Manyando C, Hooft van Huijsduijnen R, Van Voorhis WC, Wells TNC. Malaria. Nat Rev Dis Primers 2017;3:17050

3 Busto R, Dietrich WD, Globus MY, Valdes I, Scheinberg P, Ginsberg MD. Small differences in intraischemic brain temperature critically determine the extent of ischemic neuronal injury. J Cereb Blood Flow Metab 1987;7:729–738

4 Childs C, Vail A, Protheroe R, King AT, Dark PM. Differences between brain and rectal temperatures during routine critical care of patients with severe traumatic brain injury. Anaesthesia 2005;60:759–765

5 Abu-Arafeh A, Rodriguez A, Paterson RL, Andrews PJD. Temperature variability in a modern targeted temperature management trial. Crit Care Med 2018;46:223–228

6 Dehkhargani S, Fleischer CC, Qiu D, Yepes M, Tong F. Cerebral temperature dysregulation: MR thermographic monitoring in a nonhuman primate study of acute ischemic stroke. AJNR Am J Neuroradiol 2017;38:712–720

7 Rumana CS, Gopinath SP, Uzara M, Valadka AB, Robertson CS. Brain temperature exceeds systemic temperature in head-injured patients. Crit Care Med 1998;26:562–567

8 Karaszewski B, Wardlaw JM, Marshall I et al. Early brain temperature elevation and anaerobic metabolism in human acute ischaemic stroke. Brain 2009;132:955–964

9 Karaszewski B, Carpenter TK, Thomas RGR et al. Relationships between brain and body temperature, clinical and imaging outcomes after ischemic stroke. J Cereb Blood Flow Metab 2013;33:1083–1089

10 Moser E, Mathiesen I, Andersen P. Association between brain temperature and dentate field potentials in exploring and swimming rats. Science 1993;259:1324–1326

11 Henker RA, Brown SD, Marion SW. Comparison of brain temperature with bladder and rectal temperatures in adults with severe head injury. Neurosurgery 1998;42:1071–1075

12 Rossi S, Zanier ER, Mauri I, Columbo A, Stocchetti N. Brain temperature, body core temperature, and intracranial pressure in acute cerebral damage. J Neurol Neurosurg Psychiatry 2001;71:448–454

13 Thrippleton MJ, Parikh J, Harris B et al. Reliability of MRSI brain temperature mapping at 1.5 and 3 T. NMR Biomed 2014;27:183–90

14 Mariak Z. Intracranial temperature recordings in human subjects. The contribution of the neurosurgeon to thermal physiology. J Therm Biol 2002; 27:219–228. https://doi.org/10.1016/S0306-4565(01)00087-0

15 Mellergård P. Changes in human intracerebral temperature in response to different methods of brain cooling. Neurosurgery 1992;31:671–677.

16 Mellergård P. Monitoring of rectal, epidural, and intraventricular temperature in neurosurgical patients. Acta Neurochir. Suppl. (Wien) 1994;60:485–487

17 Shiraki K, Sagawa S, Tajima F, Yokota A, Hashimoto M, Brengelmann GL. Independence of brain and tympanic temperatures in an unanaesthetised human. J Appl Physiol (1985) 1988;65:482–486

18 Maloney SK, Mitchell D, Mitchell G, Fuller A. Absence of selective brain cooling in unrestrained baboons exposed to heat. Am J Physiol Regul Integr Comp Physiol 2007;292: R2059–2067

19 Hayward JN, Baker MA. Role of cerebral arterial blood in the regulation of brain temperature in the monkey. Am J Physiol 1968;215:389–403

20 Hastings MH, O’Neill JS, Maywood ES. Circadian clocks: regulators of endocrine and metabolic rhythms. J Endocrinol 2007;195:187–98

21 Conroy DA, Spielman AJ, Scott RQ. Daily rhythm of cerebral blood flow velocity. J Circadian Rhythms 2005;3:3. doi:10.1186/1740-3391-3-3

22 Obermeyer Z, Samra JK, Mullainathan S. Individual differences in normal body temperature: longitudinal big data analysis of patient records. BMJ 2017;359:j5468. doi: 10.1136/bmj.j5468

23 Morris Z, Whiteley WN, Longstreth et al. Incidental findings on brain magnetic resonance imaging: systematic review and meta-analysis. BMJ 2009;339:b3016

24 Logan RW, McClung CA. Rhythms of life: circadian disruption and brain disorders across the lifespan. Nat Rev Neurosci 2019;20:49–65

25 Takahashi JS. Transcriptional architecture of the mammalian circadian clock. Nat Rev Genet 2017;18:164–179

26 Kondratova AA, Kondratov RV. The circadian clock and pathology of the ageing brain. Nat Rev Neurosci 2012;13:325–335

27 Roenneberg T, Merrow M. The circadian clock and human health. Curr Biol 2016; 26:R432–443

28 Roenneberg T, Pilz LK, Zerbini G, Winnebeck EC. Chronotype and Social Jetlag: A (Self-) Critical Review. Biology 2019;8:54. doi:10.3390/biology8030054

29 Cady EB, D’Souza PC, Penrice J, Lorek A. The estimation of local brain temperature by in vivo 1H magnetic resonance spectroscopy. Magn Reson Med 1995;33:862–867

30 Carney N, Totten AM, O’Reilly C et al. Guidelines for the management of severe traumatic brain injury, Fourth Edition. Neurosurgery 2017;80:6–15

31 Zielinski T, Moore AM, Troup E, Halliday KJ, Millar AJ. Strengths and limitations of period estimation methods for circadian data. PLoS ONE 2014;9:e96462

32 Lueck S, Thurley K, Thaben PF, Westermark PO. Rhythmic degradation explains and unifies circadian transcriptome and proteome data. Cell Rep 2015;9:741–751

33 Babyak MA. What you see may not be what you get: a brief, non-technical introduction to overfitting in regression-type models. Psychosom Med 2004;66:411–421. doi: 10.1097/01.psy.0000127692.23278.a9

34 Erikson R. Oral temperature differences in relation to thermometer and technique. Nurs Res 1980;29:157–64

35 Ortiz-Tudela E, Martinez-Nicolas A, Campos M, Rol MA, Madrid JA. A new integrated variable based on thermometry, actimetry and body position (TAP) to evaluate circadian system status in humans. PLoS Comput Biol 2010;6:e1000996. doi:10.1371/journal.pcbi.1000996

36 Brengelmann GL. Specialised brain cooling in humans? FASEB J 1993;7:1148–1153

37 Cabanac M. Selective brain cooling in humans: “fancy” or fact? FASEB J 1993;7:1143–1147

38 Wang H, Wang B, Normoyle KP et al. Brain temperature and its fundamental properties: a review for clinicians and neuroscientists. Front Neurosci 2014;8:307. doi:10.3389/fnins.2014.00307

39 Dunlap JC, Loros JJ, DeCoursey PJ. Chronobiology: biological timekeeping. Sinauer Associates, Sunderland, Mass, 2004

40 Baud MO, Magistretti PJ, Petit J-P. Sustained sleep fragmentation affects brain temperature, food intake, and glucose tolerance in mice. J Sleep Res 2013;22:3–12

41 Skene DJ, Skornyakov E, Chowdhury NR et al. Separation of circadian- and behaviour-driven metabolite rhythms in humans provides a window on peripheral oscillators and metabolism. PNAS 2018;115:7825–7830. https://doi.org/10.1073/pnas.1801183115

42 Kemp B, Zwinderman AH, Tuk B, Kamphuisen HAC, Oberye JJL. Analysis of a sleep-dependent neuronal feedback loop: the slow-wave microcontinuity of the EEG. IEEE-BME 2000;47:1185–1194. Data set located at PhysioNet: Goldberger AL, Amaral LA, Glass et al. PhysioBank, PhysioToolkit, and PhysioNet: Components of a new research resource for complex physiological signals. Circulation 2000;101:pp.e215–e220

43 Steyerberg EW, Mushkudiani N, Perel P et al. Predicting outcome after traumatic brain injury: development and international validation of prognostic scores based on admission characteristics. PLoS Med 2008;5:e165. doi:10.1371/journal.pmed.0050165

44 Trefler A, Sadeghi N, Thomas AG, Pierpaoli C, Baker CI, Thomas C. Impact of time-of-day on brain morphometric measures derived from T1-weighted magnetic resonance imaging. Neuroimage 2016;133:41–52

45 Nakamura K, Brown RA, Narayanan S, Collins DL, Arnold DL. Alzheimer’s Disease Neuroimaging Initiative. Diurnal fluctuations in brain volume: statistical analyses of MRI from large populations. Neuroimage 2015;118:126–132

46 Jiang C, Yi L, Su S et al. Diurnal variations in neural activity of healthy human brain decoded with resting-state blood oxygen level dependent fMRI. Front Hum Neurosci 2016;10:634.

47 Arm J, Al-ledani O, Lea R, Lechner-Scott J, Ramadan S. Diurnal variability of metabolites in healthy human brain with 2D localized correlation spectroscopy (2D L-COSY). J Magn Reson Imaging 2019;50:592–601

48 Soreni N, Noseworthy MD, Cormier T, Oakden WK, Bells S, Schachar R. Intraindividual variability of striatal 1H-MRS brain metabolite measurements at 3 T. Magn Reson Imaging 2006;24:187–194

49 Maudsley AA, Goryawala MZ, Sheriff S. Effects of tissue susceptibility on brain temperature mapping. Neuroimage 2017;146:1093–1101

50 Chadzynski GL, Bender B, Groeger A, Erb M, Klose U. Tissue specific resonance frequencies of water and metabolites within the human brain. J Magn Reson 2011;212:55–63.

51 Aschoff VJ, Wever R. Spontanperiodik des menschen bei ausschluß aller zeitgeber. Naturwissenschaften 1962;49:337–342

52 Baker FC, Driver HS, Paiker J, Rogers GG, Mitchell D. Acetaminophen does not affect 24-h body temperature or sleep in the luteal phase of the menstrual cycle. J Appl Physiol (1985) 2002;92:1684–1691

53 Baker FC, Waner JI, Vieira EF, Taylor SR, Driver HS, Mitchell D. Sleep and 24 hour body temperatures: a comparison in young men, naturally cycling women and women taking hormonal contraceptives. J Physiol 2001;530:565–574

54 Yablonskiy DA, Ackerman JJ, Raichle ME. Coupling between changes in human brain temperature and oxidative metabolism during prolonged visual stimulation. Proc Natl Acad Sci USA 2000;97:7603–7608

55 Rango M, Arighi A, Bonifati C, Del Bo R, Comi G, Bresolin N. The brain is hypothermic in patients with mitochondrial diseases. J Cereb Blood Flow Metab 2014;34: 915–920

56 Kaupinnen RA, Vidyasagar R, Childs C, Balanos GM, Hiltunen Y. Assessment of human brain temperature by 1H MRS during visual stimulation and hypercapnia. NMR Biomed 2008;21:388–395

57 Birkl C, Langkammer C, Haybaeck J et al. Temperature dependency of T1 relaxation time in unfixed and fixed human brain tissue. Biomed Tech (Berl) 2013;58 Suppl 1 .pii/j/bmte.2013.58.issue-s1-L/bmt-2013-4290/bmt-2013-4290.xml. doi: 10.1515/bmt-2013-4290

58 Blowers S, Marshall I, Thrippleton M, et al. How does blood regulate cerebral temperatures during hypothermia? Sci Rep 2018;8:7877.

59 Neimark MA, Konstas A-A, Choi J-H, Pile-Spellman J. Brain cooling maintenance with cooling cap following induction with intracarotid cold saline infusion: a quantitative model. J Theor Biol 2008;253:333–44. doi: 10.1016/j.jtbi.2008.03.025

60 Larsson HBW, Courivaud F, Rostrup E, Hansen AE. Measurement of brain perfusion, blood volume, and blood-brain barrier permeability, using dynamic contrast-enhanced T(1)-weighted MRI at 3 tesla. Magn Reson Med 2009;62:1270–81. doi:10.1002/mrm.22136

61 Czeisler CA, Dumont M, Duffy JF et al. Association of sleep-wake habits in older people with changes in output of circadian pacemaker. Lancet 1992;340:933–936

62 Ferrari E, Magri F, Locatelli M et al. Chrono-neuroendocrine markers of the aging brain. Aging (Milano) 1996;8:320–327

63 Sumida K, Sato N, Ota M et al. Intraventricular temperature measured by diffusion-weighted imaging compared with brain paremchymal temperature measured by MRS in vivo. NMR Biomed 2016;29:890–895

64 Simon E. Tympanic temperature is not suited to indicate selective brain cooling in humans: a re-evaluation of the thermophysiological basics. Eur J Appl Physiol 2007;101:19–30

65 Wardlaw JM, Smith EE, Biessels GJ et al. Standards for Reporting Vascular Changes on Neuroimaging (STRIVE v1). Neuroimaging standards for research into small vessel disease and its contribution to ageing and neurodegeneration. Lancet Neurol 2013;12:822–838

66 Nedergaard M, Goldman SA. Glymphatic failure as a final common pathway to dementia. Science 2020;370:50–56

67 Rango M, Bonifati C, Bresolin N. Post-activation brain warming: a thermometry study. PLoS ONE 2015;10:e0127314

68 Karoly PJ, Goldenholz DM, Freestone DR et al. Circadian and circaseptan rhythms in human epilepsy: a retrospective cohort study. Lancet Neurol 2018;17:977–985

69 Nomura S, Kida H, Hirayama Y et al. Reduction of spike generation frequency by cooling in brain slices from rats and from patients with epilepsy. J Cereb Blood Flow Metab 2019;39: 2286–2294

70 May A, Bahra A, Büchel C, Frackowiak RS, Goadsby PJ. Hypothalamic activation in cluster headache attacks. Lancet 1998;352:275–278

71 Lodi R, Pierangeli G, Tonon C et al. Study of hypothalamic metabolism in cluster headache by proton MR spectroscopy. Neurology 2006;66:1264–1266

72 Baker FC, Driver HS, Rogers GG, Paiker J, Mitchell D. High nocturnal body temperatures and disturbed sleep in women with primary dysmenorrhea. Am J Physiol 1999;277:E1013–E1021

73 Grant LK, Gooley JJ, St Hilaire MA et al. Menstrual phase-dependent differences in neurobehavioural performance: the role of temperature and the progesterone/estradiol ratio. Sleep 2020;43:zsz227. doi: 10.1093/sleep/zsz227

74 Nakayama T, Suzuki M, Ishizuka N. Action of progesterone on preoptic thermosensitive neurons. Nature 1975;258:80

75 Kruijver FP, Swaab DF. Sex hormone receptors are present in the human suprachiasmatic nucleus. Neuroendocrinology 2002;75:296–305

76 Ma J, Huang S, Qin S, You C, Zeng Y. Progesterone for acute traumatic brain injury. Cochrane Database Syst Rev 2016;12:CD008409

77 Geneva II, Cuzzo B, Fazili T, Javaid W. Normal body temperature: a systematic review. Open Forum Infect Dis 2019;6:ofz032. doi: 10.1093/ofid/ofz032

78 Hassager C, Nagao K, Hildick-Smith D. Out-of-hospital cardiac arrest: in-hospital intervention strategies. Lancet 2018;391:989–998

79 Fuller A, Carter RN, Mitchell D. Brain and abdominal temperatures at fatigue in rats exercising in the heat. J App Physiol 1998;84:877–883. doi: 10.1152/jappl.1998.84.3.877

80 Cariou A, Payen J-F, Asehnoune K et al. Targeted temperature management in the ICU: Guidelines from a French expert panel. Anaesth Crit Care Pain Med 2018;37:481–491.

81 Madden LK, Hill M, May TL et al. The implementation of targeted temperature management: an evidence-based guideline from the Neurocritical Care Society. Neurocrit Care 2017;27:468–487

82 Nanba T, Nishimoto H, Yoshioka Y et al. Apparent brain temperature imaging with multivoxel proton magnetic resonance spectroscopy compared with cerebral blood flow and metabolism imaging on positron emission tomography in patients with unilateral chronic major artery steno-occlusive disease. Neuroradiology 2017;59:923–935

83 Tsutsui S, Nanba T, Yoshioka Y et al. Preoperative brain temperature imaging on proton magnetic resonance spectroscopy predicts hemispheric ischemia during carotid endarterectomy for unilateral carotid stenosis with inadequate collateral blood flow. Neurol Res 2018;40:617–623

84 Marshall I, Karaszewski B, Wardlaw JM et al. Measurement of regional brain temperature using proton spectroscopic imaging: validation and application to acute ischemic stroke. Magn Reson Imaging 2006;24:699–706

85 Sone D, Ikegaya N, Takahashi A et al. Noninvasive detection of focal brain hyperthermia related to continuous epileptic activities using proton MR spectroscopy. Epilepsy Res 2017; 138:1–4

86 Shiloh R, Kushnir T, Gilat Y et al. In vivo occipital-frontal temperature-gradient in schizophrenia patients and its possible association with psychopathology: a magnetic resonance spectroscopy study. Eur Neuropsychopharmacol 2008;18:557–564.

87 Nikitopoulou G, Crammer JL. Change in diurnal temperature rhythm in manic-depressive illness. BMJ 1976;1:1311–1314

88 Niven DJ, Gaudet JE, Laupland KB, Mrklas KJ, Roberts DJ, Stelfox HT et al. Accuracy of peripheral thermometers for estimating temperature: a systematic review and meta-analysis. Ann Intern Med 2015;163:768–777

89 Papaionnou VE, Chouvarda IG, Maglaveras NK, Pneumatikos IA. Temperature variability analysis using wavelets and multiscale entropy in patients with systemic inflammatory response syndrome, sepsis, and septic shock. Crit Care Lond Engl 2012;16:R51

90 Childs C, Lunn KW. Clinical review: brain-body temperature differences in adults with severe traumatic brain injury. Critical Care 2013;17:222. http://ccforum.com/content/17/2/222

