## Supplementary Information for "Diurnal brain temperature rhythms and mortality after brain injury: a prospective and retrospective cohort study"

**Contents**

Supplementary Methods
Supplementary Text
Figures S1 to S9
Tables S1 to S3
Captions for Movies S1 to S2
Supplementary references
Supplementary Appendices 1-5

**Other Supplementary information for this manuscript includes:**

Movies S1 to S2 (also located at https://www2.mrc-lmb.cam.ac.uk/groups/oneill/research/heatwave/).

### SUPPLEMENTARY METHODS

#### Actigraphy and chronotyping

Chronotype varies between sexes in an age-dependent manner,[1] as well as between individuals of the same sex. Actigraphy is a validated, non-invasive, and objective tool for studying sleep and circadian patterns.[2-5] We chose the ActTrust2 Wrist Actimeter (Condor Instruments, Sao Paulo, Brazil), sampling the following parameters at a frequency of 60 seconds: external temperature, skin temperature, rest/activity/sleep patterns, and various wavelengths of light (infrared, red, green, blue, UVA and UVB). All scanned participants wore their Actimeter for a minimum of 5 days and nights; they were asked to press an event button on the device once just before trying to sleep each night and again when they first awoke each morning. These reported sleep and wake times were validated against the actigraphy data and confirmed verbally with each participant on their scanning day. Data was transferred using the ActTrust Dock, then processed and analysed using ActTrust Studio software and an in-built Condor Instruments algorithm which uses Proportional Integral Mode (PIM) and Zero Crossing Mode normalized per second (ZCMn) data.[6] The light/dark phase was programmed to reflect the latitude and longitude (time zone with Coordinated Universal Time (UTC) offset of +1h) in Edinburgh, United Kingdom. Devices were retrieved, and the data downloaded and analysed, on the day of scanning; participants were asked to report which (if any) main sleep periods in the preceding week ended naturally ('free') or with an alarm or other disturbance ('scheduled'). Free and scheduled assignments were validated against the data. Sleep scoring parameters were enabled to include one or more main periods of sleep at night. Periods of 'off-wrist' time were excluded from the analysis. Sleep scoring was performed using the Cole-Kripke method (ActStudio software) to extract seven key sleep parameters for each night of sleep.[6] ActTrust2 data underwent further processing to derive eleven diurnal characteristics, as well as $MSF_{sc}$ and sleep-corrected social jetlag ($SJL_{sc}$).[7-9]

#### Scanning protocol

The scanning protocol was adapted from Thrippleton et al*.*, 2013.[10] Localizers and structural sequences were acquired prior to MRS to plan voxel placement. Structural acquisition included whole-brain axial T2-weighted (T2w; three-dimensional fast spin-echo; TR/TE = 3200/408 ms; 1-mm isotropic resolution) and T1-weighted (T1w; three-dimensional inversion recovery-prepared gradient echo; TR/TI/TE = 2500/1100/4.37 ms, flip angle, 7°; 1-mm isotropic resolution) sequences. Structural imaging was repeated at each scanning session and the last-acquired structural images were used for HRF screening. Magnetic resonance spectroscopic imaging (MRSI; semi-LASER sequence, TR/TE = 1200/144ms)[11] of the cerebrum was acquired from a 10-mm thick axial slice located at the level of the centrum semiovale, generating multiple 10 × 10 × 10 mm voxels (Fig.2A). Six saturation bands were applied to suppress scalp lipid and other signals on all faces of the volume of interest. Automated shimming and partial water suppression were applied. For each phase-encoding step, a 512-ms free induction decay (FID) was obtained. Since a single-voxel semi-LASER MRS sequence was not available at the time of the study, single-voxel PRESS MRS acquisition (TR/TE = 1200/144 ms) was used to sample the thalamus (voxel size 15 × 15 × 15 mm) and hypothalamus (voxel size 20 × 10 × 10 mm) (Fig.2A). Single voxels were positioned to avoid interference from cerebrospinal fluid and/or large blood vessels; spectral quality was visually assessed in real-time to ensure that the water and NAA peaks were of a sufficient signal-to-noise ratio and the acquisition was repeated if necessary. Voxel position and orientation was matched between scanning sessions for each participant by a single radiographer who acquired all data for that individual at each time point. To maximise consistency of voxel positioning between participants, an illustrated placement protocol, specified by a senior neuroradiologist, was used. Foam pads were wedged between the skull and the head coil to eliminate any movement during acquisition. The whole procedure was optimized through pilot scanning, achieving a total scan time of ~30min, including shimming. All structural brain images were reviewed by a consultant neuroradiologist (GM). Further details on the scanning protocol are in Appendix 3.

**MRS data processing**

MRS data were processed as described previously.[10] Briefly, free induction decays were apodised (Gaussian function, 4 Hz full width at half height) and the NAA and water chemical shifts determined by time-domain fitting of both resonances. For each voxel, four quality control filters were applied: a cut off for NAA linewidth (13 Hz for multivoxel data; 15.5 Hz for single voxel data), an R-squared measure of the NAA fit quality (0.80), a cut off for $H_2O$ linewidth (13 Hz for multivoxel data; 15.5 Hz for single voxel data), and an R-squared measure of $H_2O$ fit quality (0.93). Whilst the consensus recommendation threshold for $H_2O$ linewidth is 0.1 ppm = 12 Hz,[12] the linewidth cut-offs used here included the effect of the Gaussian apodization applied. Hypothalamus and thalamus are challenging to shim, and the single voxels were larger than those acquired with the multivoxel sequence. For these reasons, the single-voxel data was expected to be of lower quality with a slightly increased linewidth (reflected in the higher linewidth cut-off). Data from a voxel was discarded if it failed one or more of the set cut-offs and if the resultant data point affected the range of $T_{Br}$ values for that participant at that time point, and/or the range for that voxel at that time point across a given sex group. The means and standard deviations for linewidths for NAA and $H_2O$ are reported in Table S3; spectral curve fitting and peak annotation are depicted in fig.S4. Spectral fitting was performed in batch mode without investigator intervention. Processing of MRS data was fully automated.

**Data management**

Source data for the prospective study included electronic screening questionnaire responses (hosted by Jisc online surveys, Bristol, UK), actigraphy data logs (electronic format) and Study Participant Data Forms (Appendix 4; hard and electronic format, hosted by the Centre for Clinical Brain Sciences, University of Edinburgh), DICOM files and other raw imaging data (stored at Edinburgh Imaging, hosted by the University of Edinburgh). Clinical imaging reports used for HRF screening were hosted by NHS Lothian; no members of the Study research team had access to these reports apart from the designated Neuroradiologist (GM). Data derived from human participants was managed and shared according to terms described in the Consent to Participate Form (Appendix 1) and Data Protection Information Sheet, in compliance with the revised GDPR (https://www.eugdpr.org). Circular analyses as well as generalized linear and linear mixed modelling were performed using R version 3.6.3 (R Core Team, 2020) and the circular (v0.4–93; Lund et al., 2017); cosinor (v1.1; Sachs 2015), cosinor2 (v0.2.1; Mutak 2018), lme4 (v1.1–23; Bates et al. 2020), effects (v4.1-4; Fox et al. 2019), afex (Singmann et al., 2020), Matrix (v1.2-18; Bates et al., 2019), Cairo (v1.5-12.2; Urbanek and Horner 2020), yarrr (v0.1.5; Phillips 2017) and car (v3.0-8; Fox et al., 2020) packages. The full reproducible code is provided in Appendix 5, or is available on request to the Lead Author. All other analyses were performed in GraphPad Prism version 8.2.

**Rationale for generalized linear mixed model**

A GLMM requires that (1) the outcome is not normally distributed but the distribution is known (here, our outcome variable is binomial 'death or survival'), and (2) that there is more than one source of random error. The outcome itself is not linearly related to the predictor(s) but a function of it (logit for logistic regression) is. In our model, death is defined as a success or 'hit', and survival as a 'miss'. Our observations of 'death' or 'survival' are equal to '1' or '0' and the probability of death *p* lies between these two values. The logit (log of the odds) is thus a way of linking our GLMM to a non-normal distribution of one of two outcomes. In Fig.5A-C, plots are provided for those readers who prefer to have a graphical presentation of GLMM outputs that enables visualization of all data points. However, since log of the odds is hard to interpret, and for greater accessibility, Fig.5D presents the regular odds of death for each predictor. The odds of death = probability of death/(1-probability of death) = $p/(1-p)$. We transform from log odds back to regular odds using the exponential function; for this we also have to transform the 95% confidence intervals and in doing so, we are most interested in whether the confidence interval contains 1 (a confidence interval that contains 0 in log odds will always contain 1 when transformed to regular odds).

Our GLMM is designed as a predictive model (what x's predict y), not an explanatory one (what x's are truly related to y). For a predictive model, we are not concerned about confounders if the model makes accurate predictions using the variable of interest. To this end, based on our results, we might consider $T_{Br}$ rhythmicity as a proxy variable; in reality, this variable is most probably directly relevant to outcome, but our model is not designed to test this. There are many other factors that may have influenced temperature and/or outcome in our TBI cohort,[13] however for the question we are asking of our retrospective data, we are not interested in how or why the temperature rhythm exists or does not exist—we are simply asking whether the presence/absence of a temperature rhythm correlates with odds of death in intensive care.

### SUPPLEMENTARY TEXT

**Internal rhythms and health**

The body clock—our internal circadian rhythm—anticipates the day-night cycle, thus optimizing every aspect of our physiology to solar time.[14] Circadian rhythms are fine-tuned by environmental cues, which feed into a multi-oscillator system; a 'master' clock within the suprachiasmatic nuclei (SCN) of the hypothalamus synchronizes cell-autonomous clocks throughout the brain and periphery, coupling systemic processes to light-dark transitions.[23] With age, this timekeeping becomes less robust—a situation compounded by modern living, which dissociates our body clocks from natural cues.[15,16] Shift-work, travel, and 'social jet-jag' all disrupt our sleep and circadian health.[9,17] Globally, neurological conditions are the primary cause of disability and second leading cause of death.[18] Several brain disorders including traumatic and vascular events, neurodevelopmental and mood problems, and neurodegenerative diseases are associated with circadian disruption; others, such as epilepsy, display symptomatic coupling to the body clock.[14,19-27] A mechanistic link between circadian and brain dysfunction is yet to be established, but a poorly explored factor is $T_{Br}$.[14] This is despite the fact that elevated temperature and increased temperature variability are considered prognostic indicators after brain injury.[28-34]

**Technical limitations of brain thermometry**

Direct $T_{Br}$ measurements from human patients are confounded by variable brain pathology, anaesthesia or sedation, TTM protocols, and/or sampling from subdural or intraventricular sites. MRS brain thermometry is attractive, but controversy remains over the ideal calibration method. Common approaches use phantoms to mimic brain tissue, or directly-measured $T_{Br}$ curves from other species under anaesthesia (typically neonates), often reporting $T_{Br}$ values that are substantially lower than direct normothermic $T_{Br}$ measurements. Patient MRS studies have focused on relative $T_{Br}$ changes in response to injury or other pathology.[32,35-41] At the time of writing, we are aware of 25 published original research articles that used MRS for brain thermometry in healthy adults (Table S1). Sex was reported in 22 of these studies; 21 of them included <36 participants per experiment—a total of 329 participants including 105 women. Only two studies made sex comparisons for $T_{Br}$, both reporting no statistical difference.[42,43] One retrospective study (n=150 with 90 females) found a 0.1°C higher cerebral $T_{Br}$ in females;[44] smaller than the sex difference found in our cohort. However, no prior studies controlled for menstrual cycle phase, which has enabled us to reveal physiologically-relevant differences between the sexes in all brain regions measured. In addition to menstrual cycle, the large retrospective study of Maudsley et al.[44] did not control for age, time of day, chronotype, or medications, all of which may have contributed to high variance in their dataset. Eight MRS brain thermometry studies reported time period of scanning, but only 6 of these arguably controlled for time of day (scanning window 2–3 hours).

Recent studies have emphasized the need for more accurate calibration and potentially brain region-specific temperature coefficients to account for differences in tissue content, microstructure and orientation.[10,44-46] However, using methods described by Marshall[38] and Thrippleton et al.[10] we have now obtained data that are absolutely consistent with published direct $T_{Br}$ measurements obtained from healthy, conscious, non-human primates, and normothermic human patients.[47-50] The limitations of MRS thermometry dictate that 'absolute' temperatures cannot be estimated with the same confidence as relative temperature differences. That said, the mean global temperatures we have observed lie, as expected, between average human core $T_{Bo}$[51,52] and direct brain temperatures obtained from other species and human patients with brain injury (fig.S8).[47,50,53,54] To avoid concerns arising from potential inaccuracy of the calibration intercept, we have focused our analysis on relative $T_{Br}$ changes, for which MRS thermometry is exquisitely well-suited.[10] Given the propensity for major brain structural changes with age and neurodegeneration, exploring time of day variation at different life stages, rather than single time-point variation over months or years, would circumvent some of the inherent limitations of the technique. Ultimately, physiological and clinical understanding should be informed by experimental observation. MRS thermometry has high spatial resolution and is extremely sensitive to temperature change within a tissue, but the technique can only ever have low temporal resolution. Moreover, it cannot be used for repeated measurements of brain-injured patients; whereas, direct temperature probes measure absolute temperature with very high temporal resolution, but cannot be used in healthy individuals. We have found these two approaches of $T_{Br}$ measurement to be mutually supportive, with the temperature probe data and repeated measurements of individuals allowing us to be much more confident about the fidelity of the MRS data than in any previous study. Temperature variations were further supported by high temporal resolution actigraphy data in our healthy cohort, and core $T_{Bo}$ data in TBI patients.

**Temperature gradients**

In our healthy cohort, $T_{Br}$ was always higher than oral temperature. The direction of this brain-body temperature gradient contradicts some studies,[55,56] but agrees with others.[42,55,57] Our findings are corroborated by a case report documenting parallel oesophageal, tympanic, intraventricular, and cortical white matter temperature in an unanaesthetised, normothermic patient.[58] The relationship between sites was fixed, with cortical $T_{Br}$ > ventricular > oesophageal > tympanic.[58] During an overnight recording, cortical $T_{Br}$ declined from ~38.1°C at 9pm to a minimum of ~36.7°C at 3am, as expected during sleep. Considering this 1.4°C decrease over 6 hours, and extrapolating back to the predicted maximum $T_{Br}$ for this male patient (~3pm), maximum $T_{Br}$ in this region would have been ~38.7°C—practically indistinguishable from the mean $T_{Br}$ we found in healthy male parietal white matter in the afternoon (Fig.3). In the report, $T_{Br}$ exceeded oesophageal temperature by ~0.7°C, and so our measured difference between oral temperature and $T_{Br}$ is realistic. A similar brain-body temperature relationship was found in healthy volunteers with a mean parietal MRS-derived $T_{Br}$ of 38.1°C; 1.3°C higher than rectal temperature.[43] Our retrospective analysis indicates that a positive $T_{Br}$-$T_{Bo}$ gradient cannot be attributed entirely to local injury (fig.S7) and so, as for non-human primates,[47,48] it is likely a normal physiological phenomenon in humans. Indeed, maintenance of this gradient was associated with a better outcome in severe head injury.[59]

MRS-derived estimates of absolute $T_{Br}$ often conflict with known $T_{Br}$-$T_{Bo}$ gradients in other species, including non-human primates.[47,48] A recent 7T MRS-based $T_{Br}$ study in healthy sheep found a brain-regional $T_{Br}$ gradient, but global $T_{Br}$ was 0.7°C lower than $T_{Bo}$ (38.5 versus 39.2°C).[54] This inversion of the brain-body temperature gradient might be explained by several factors: (i) that measurements were conducted under general anaesthesia (which inverts temperature gradients in some species), (ii) the species under test has defined neurovascular anatomy to effect selective brain cooling (SBC; a carotid *rete mirabile* is absent in primates and the existence of other SBC mechanisms in humans and non-human primates is highly contentious),[47,50,60,61] and (iii) it remains possible that absolute mean global $T_{Br}$ is the key defended parameter across mammals, whilst $T_{Bo}$ can vary more widely in a species-specific manner ($T_{Bo}$ of most domestic livestock is higher than human $T_{Bo}$). In seminal work by Fuller et al., diurnal variations in colonic and hypothalamic temperatures were explored simultaneously in unsedated male squirrel monkeys.[50] At various ambient temperatures ($T_a$ of 20, 26, 32, and 36°C), there was no change in the phase or period length of either temperature rhythm, and hypothalamic always exceeded colonic temperature except at $T_a$ 36°C. Mean temperatures at both sites increased with increasing $T_a$, and there was a clear reduction in amplitude in both temperature rhythms, primarily resulting from an increase in temperature minima during the night. Importantly, hypothalamic temperatures ranged from 37.5–39.1°C between the afternoon and late evening at $T_a$ 26°C, remarkably similar to the hypothalamic temperature range observed by MRS in male subjects here (Fig.3).[50] Compelling evidence for the absence of SBC in primates comes from a study in unrestrained baboons, where hypothalamic temperature was consistently higher than carotid arterial or abdominal temperature, ranged between 37.5 and 38.8°C under cycling $T_a$ conditions (15–35°C), and from 37.5–40.5°C when water was restricted.[47] Our results for this brain region are thus entirely consistent with direct temperature measurements from conscious healthy animals of closely related species.

### Temperature rhythms and sleep

Temperature rhythms emerge immediately after birth and peak in amplitude during childhood as the brain matures.[14] $T_{Bo}$ rhythms then phase-delay in adulthood, and gradually reduce in amplitude and phase-advance with ageing.[14] Daily variations in heat loss (rather than heat production) are considered to drive these daily changes in temperature;[62] it follows that an increase in minimum $T_{Br}$ (and consequently, a reduced $T_{Br}$ rhythmic amplitude) with ageing must represent a failure of heat loss. It has been estimated that $T_{Br}$ would increase by ~0.28°C/min if the key routes of heat dissipation did not exist,[63] so even modest cerebrovascular compromise is likely to result in net heat gain. This is not necessarily at odds with findings in ischaemic stroke, where $T_{Br}$ is higher in the lesion penumbra than the lesion core.[33,38] At the core, total loss of perfusion would lead to a $T_{Br}$ drop as observed during brain death.[64] A vascular-driven disruption of $T_{Br}$ rhythm reconciles various risk factors for non-convulsive seizures during sleep in dementia patients with some of the temperature-sensitive molecular aberrations in animal models of these disorders,[65,66] and potentially even disease-specific distribution of protein pathology. Rather than an inherent vulnerability of specific, synaptically-connected neurons, or an age-related decline in glymphatic clearance,[17] the predictable 'spread' of protein aggregates through the brain in neurodegenerative disease may simply reflect age-related progression of cooling failure through vascular compromise. This is conceptually straightforward for chronic traumatic encephalopathy, where tau pathology is concentrated at cerebral sulci surrounding repetitive microvascular trauma.[67] The propensity of mutant protein forms to aggregate in familial disease may signify a heightened sensitivity to temperature-driven effects on biochemical modification in regions of greatest diurnal $T_{Br}$ variability.[68,769] Notably, the hypothalamus (and in particular the SCN) is one of the most densely vascularized regions of the mammalian brain.[70] This is the first study to measure $T_{Br}$ in the human hypothalamus; future studies should consider whether the position and vascularity of this structure optimizes its circadian and thermoregulatory roles, whilst shielding it from high absolute temperatures. Epilepsy is bimodally distributed but the highest incidence is in adults >75 years, where cerebrovascular disease causes more than a third of cases.[65,71] Alzheimer's disease and late-onset epilepsy may be pathologically linked through vascular changes, tau pathology, or both, and sleep deprivation can unmask a life-long tendency to seizures.[65] Given the temperature-sensitivity of tau protein modifications, the role of brain vasculature in effecting $T_{Br}$ changes, and the importance of temperature in sleep, impaired $T_{Br}$ rhythms might contribute to this pathological link.[68,72] In everyday clinical terms however, cerebrovascular impairment is not the only route to a $T_{Br}$ increase; nasal airflow is clearly inhibited during prolonged wearing of facemasks,[73,74] or indeed bypassed all together in intubated patients. That temperature variation is such as strong predictor of mortality (Fig.5) underpins an urgent need for further chronotype-controlled research in this area.[28,51]

Superimposed on daily temperature rhythms is a daily oscillation in thermoregulatory capacity which is relatively impaired during sleep.[62] This may have contributed to the increased variability in $T_{Br}$ in participants that reportedly slept during their scans (Fig.2B). Whilst the preoptic area and anterior hypothalamus are the main loci for $T_{Bo}$ regulation, their thermoregulatory function appears to suppress, rather than coordinate, the circadian organization of $T_{Bo}$—a tenet of the SCN.[14] Studies in several mammalian species including humans have suggested that circadian $T_{Br}$ and sleep-wake cycles are not interdependent, but that under free-running conditions, the period length of the sleep-wake cycle is negatively correlated with average core $T_{Bo}$.[75-79] In rats, ultradian (<24-hour) $T_{Br}$ rhythms are superimposed on circadian $T_{Br}$ rhythms.[80] These high-frequency rhythms are closely associated with sleep-wake states, and persist in the absence of SCN input which abolishes the circadian rhythm in $T_{Br}$.[80] Rodents however are not small humans; there are critical differences in thermoregulation strategy and rate of heat gain/loss across mammals which make it difficult to translate temperature-related phenomena between species of vastly different sizes.[81] Such caveats aside, it is clear that changes in $T_{Br}$ can affect synaptic structure, function, and plasticity in mammals,[82,83] and may even influence the diurnal variation in glymphatic clearance of 'brain waste'.[17,84] This clearance in awake rats seems to be greatest during the rest phase, at a time when $T_{Br}$ would be lowest and CBF increased.[84-86] On the one hand, increased CBF might simultaneously enhance the removal of brain heat whilst favouring pulsatile glymphatic clearance; on the other it could reduce the perivascular space available for glymphatic flow through hydrostatic means, particularly in the presence of hypertensive cardiovascular disease.[17] We found no significant correlation between $T_{Br}$ and subject-reported sleep during scanning (Appendix 5). However, we did not employ a gold standard method for sleep measurement (polysomnography) and since the scan time was around only 30 minutes (and the MRS acquisition much less than this), we would not expect to observe a significant effect. Irrespectively, our late evening $T_{Br}$ data confirms that sleep onset coincides with a reduction in both core $T_{Bo}$ and $T_{Br}$, assisted by a selective redistribution of blood flow (Fig.1B-C), and presumably modulated by melatonin.[62,87,88-90] Moving forward, it will be important to consider the role of this hormone, and whether its rhythmic disruption in intensive care patients affects its rhythmic differential modulation of regional vasculature.[90,91]

**Limitations of retrospective TBI analysis**

There are many factors (internal and external to the patient) that might impact upon $T_{Bo}$, $T_{Br}$, or mortality risk in TBI patients.[13] We could only test for relationships among the limited number of parameters for which data were available in our patient cohort, and that could be accommodated by our sample size without overfitting the outcome model. Our model thus posed a very simple question of whether the presence or absence of a $T_{Br}$ rhythm correlates with the odds of death, without implying causality in either direction. Despite our inability to adjust for several more conventional parameters such as injury severity, our analysis suggests that loss of a diurnal $T_{Br}$ rhythm is a very powerful predictor of mortality compared to prognostic factors reported historically, either alone, or in combination.[13] One interpretation is that diurnal $T_{Br}$ rhythm together with age might be a very simple way to accurately predict ICU outcome after brain injury—a rigorous prospective clinical study is of course needed test this hypothesis. The fact that presence of a daily $T_{Br}$ rhythm did not correlate with either age or mean $T_{Br}$ indicates that this parameter has additional predictive value in its own right.

**Clinical applications**

Impaired core $T_{Bo}$ rhythmicity correlates with injury severity and worsened behavioural measures for patients in vegetative or minimally conscious states.[19,20] One study reported that only 29% of trauma patients had a $T_{Bo}$ rhythm period of 22–26h, and circadian disruption has been associated with poor outcome after both head trauma and subarachnoid haemorrhage.[92,93] $T_{Br}$ was not measured in these patients, but our findings (Fig.5) suggest that relationships between temperature rhythm and outcome would be clearer if $T_{Br}$ was included for monitoring and prognostication. Indeed, functional scoring of comatose patients varies by time of day, with better scores around maximum $T_{Bo}$.[92] Based on our new data, we recommend review of existing guidelines for management of TBI patients in intensive care. Specifically, we propose that direct $T_{Br}$ (in addition to core $T_{Bo}$) becomes the clinical standard for temperature monitoring in these patients, and that, alongside other clinical parameters, diurnal *temperature variation* should be considered a core element of treatment plans and prognostication. At the very least, time of day should be considered when scheduling assessments, and ideally, patient-specific temperature rhythm and chronotype should be accounted for when interpreting results that influence life-changing clinical decisions. Future studies should standardize the method of $T_{Bo}$ measurement, and the location of intracranial probes with respect to focal brain pathology. By contrast, HEATWAVE would find greatest practical utility for chronic brain disease, since our voxel maps give an approximation of the $T_{Br}$ expected in each brain location at three clinically-relevant times of day, and how much $T_{Br}$ should vary between voxels at a given time point. Since each data point in each map is averaged from multiple healthy individuals, it incorporates the range of ages, BMIs, and chronotypes for each sex in the demographic tested.

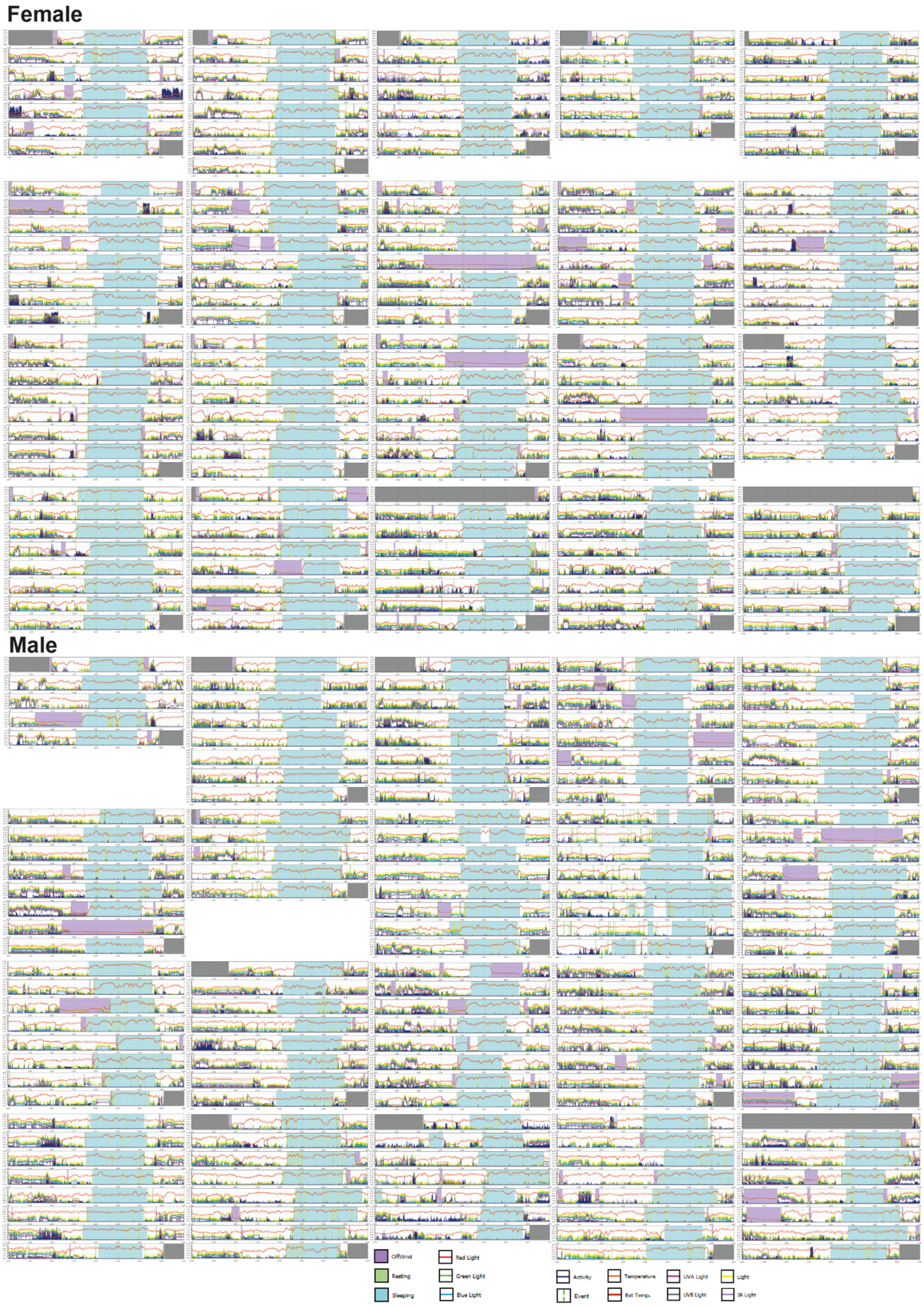
Female
Male
OffWrist
Resting
Sleeping
Red Light
Green Light
Blue Light
Activity
Event
Temperature
Ext Temp.
UVA Light
UVB Light
Light
IR Light

**Fig. S1. Actograms in healthy volunteers.** Montage of actograms in 40 healthy volunteers. Most participants wore an ActTrust2 device for eight days. Actograms are presented in order of acrophase (earliest top left, latest bottom right) in females and males separately. Note increase in participant wrist skin temperature during sleep phase, reflecting peripheral vasodilation (core body temperature is expected to drop in anti-phase to a skin temperature increase during sleep).[94]

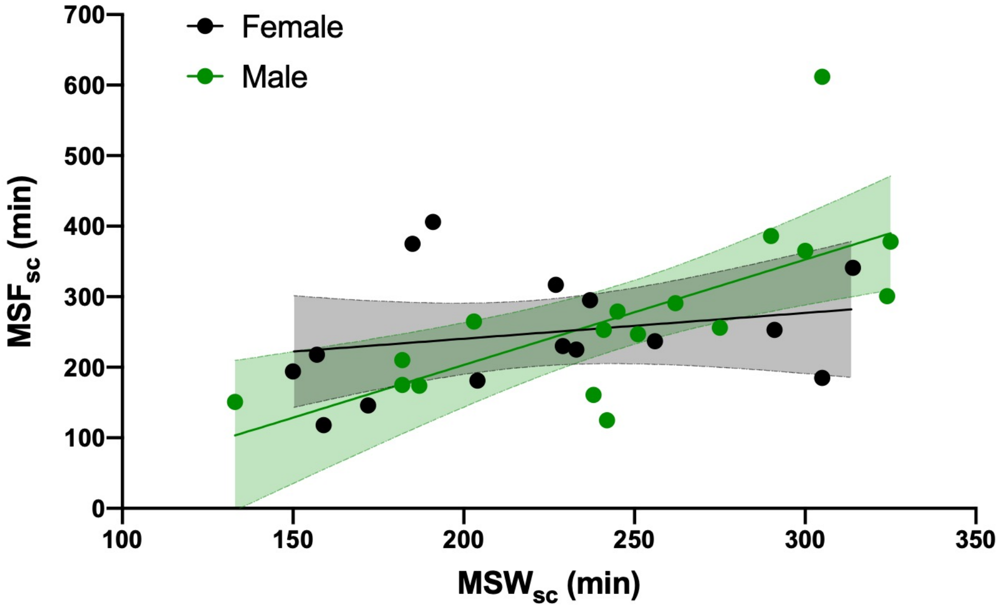


**Fig.S2. Interpolation of $MSF_{sc}$.** Five females and three males reported no free days during actigraphy sampling; their $MSF_{sc}$ was interpolated by linear regression with the sleep-corrected midpoint of sleep on work days ($MSW_{sc}$). Regression was performed for males and females separately.

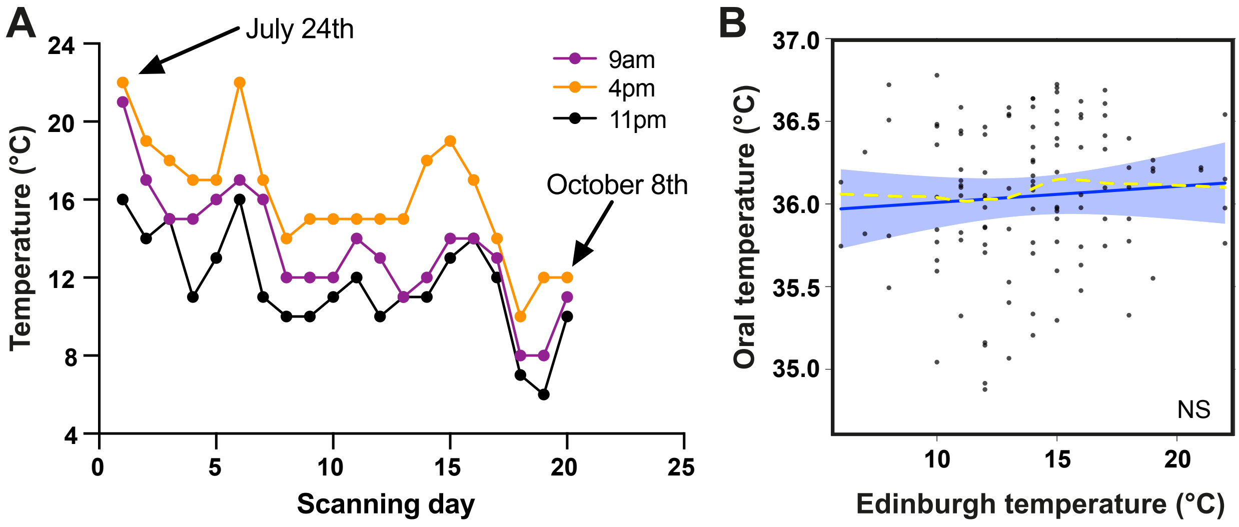


**Fig. S3. Environmental temperature variations.** (**A**) Recorded outdoor air temperatures at 1.5m above ground level in Edinburgh at times of scanning sessions, on each day of scanning during data collection period. Collection period limited to 14 weeks to minimize any effect of seasonal variation in environmental temperature and light. For actigraphy data extraction from each participant, daily changes in sunrise and sunset times were adjusted according to date of device retrieval using date and location function in ActStudio. (**B**) Linear mixed model results for oral temperature by environmental temperature. Solid blue line represents model fit, shaded areas represent 95% confidence intervals, dark grey circles display residuals (single temperature data points), and smoothed dashed yellow line represents partial residuals.

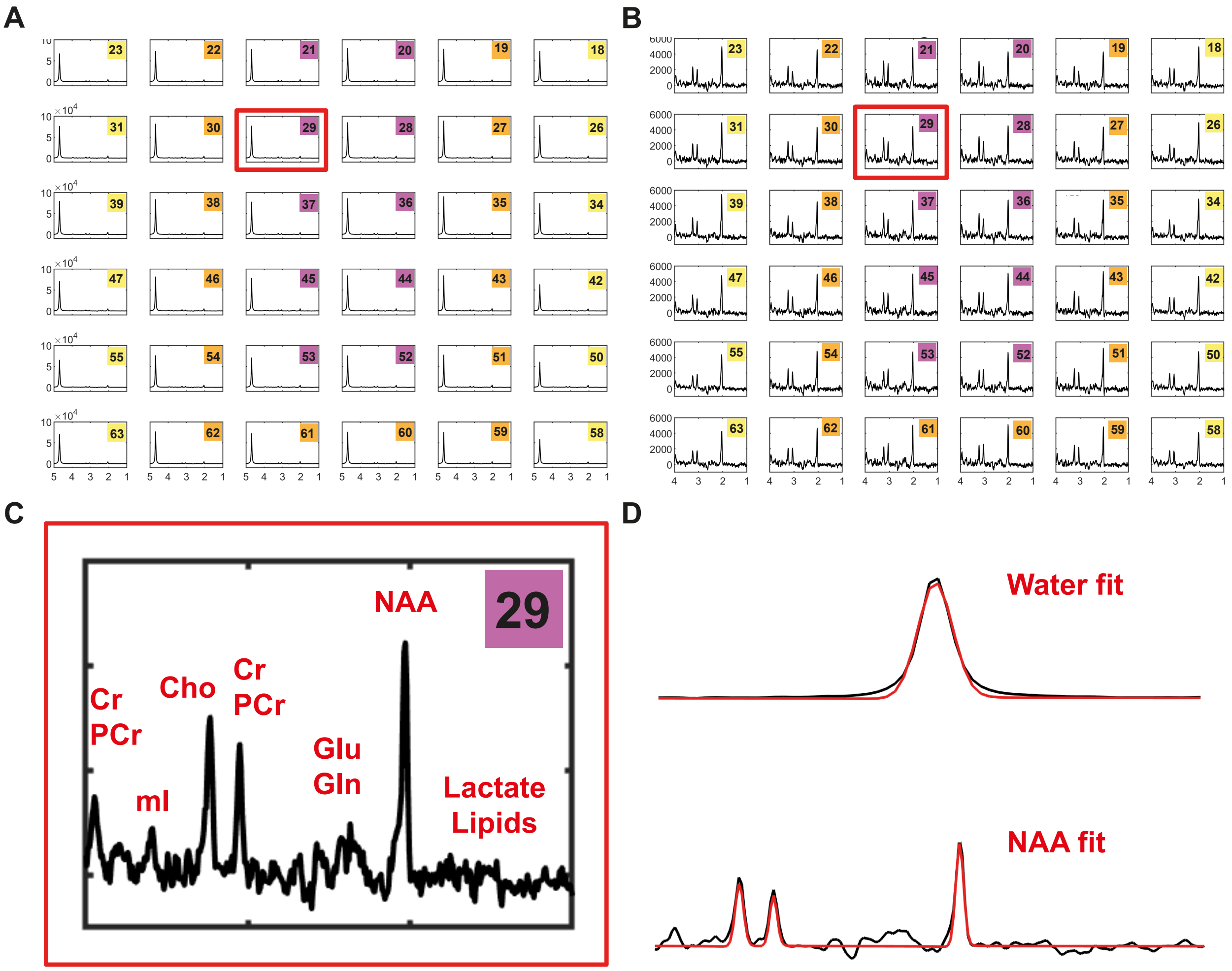


**Fig. S4. MRS data processing.** (**A**) Example spectra from central 36 voxels from cerebral region; voxel numbering as in Fig.2. At this scale, only the large water peak to the left of each spectrum is clearly visible; smaller peaks to the right of this are barely visible. Selected voxel (29) shown in red box. (**B**) Zoomed-in representation of voxel 29, focusing on smaller peaks; the water peak is off-chart (and off-scale) to the left. The largest peak visible in the spectrum is now NAA. (**C**) Enlargement of voxel from (**B**) to show peak annotation; Cr, creatine; PCr, phosphocreatine; mI, myo-inositol; Cho, choline; Glu, glutamate; Gln, glutamine; NAA, N-acetylaspartate. (**D**) Representative examples of spectral fitting of water and NAA peaks for voxel 29. All but one of the 24 rejected $T_{\mathrm{Br}}$ data points were obtained from the most rostro-lateral voxel of the right frontal lobe.

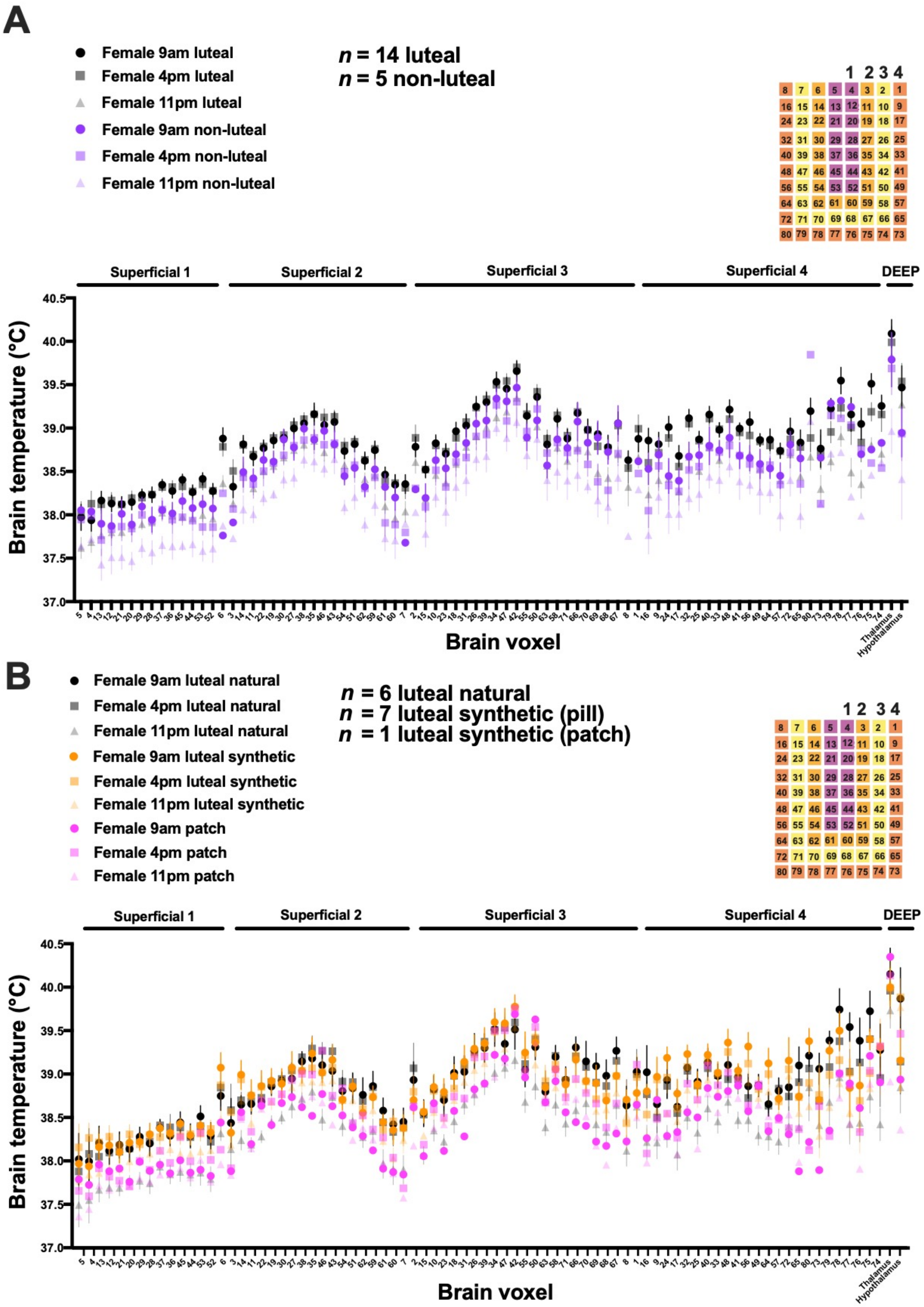
A
Female 9am luteal
Female 4pm luteal
Female 11pm luteal
Female 9am non-luteal
Female 4pm non-luteal
Female 11pm non-luteal
n = 14 luteal
n = 5 non-luteal
Superficial 1
Superficial 2
Superficial 3
Superficial 4
DEEP
Brain temperature (°C)
Brain voxel
Thalamus
Hypothalamus
B
Female 9am luteal natural
Female 4pm luteal natural
Female 11pm luteal natural
Female 9am luteal synthetic
Female 4pm luteal synthetic
Female 11pm luteal synthetic
Female 9am patch
Female 4pm patch
Female 11pm patch
n = 6 luteal natural
n = 7 luteal synthetic (pill)
n = 1 luteal synthetic (patch)
Superficial 1
Superficial 2
Superficial 3
Superficial 4
DEEP
Brain temperature (°C)
Brain voxel
Thalamus
Hypothalamus

**Fig. S5. Female brain temperature by menstrual cycle phase. (A)** Female $T_{Br}$ data split into luteal and non-luteal groups. Menstrual cycle phase explained 15.2% (P=0.0006) and time of day explained 21.2% (P=0.0004) of the variation in $T_{Br}$, respectively (two-way ANOVA). The interaction between these two factors was not significant. (**B**) Luteal-phase female $T_{Br}$ plotted according to contraception type. $T_{Br}$ did not differ between luteal females with natural menstrual cycles and those taking the combined contraceptive pill (mixed-effects analysis with Tukey's multiple comparisons test). Synthetic steroid hormones influence $T_{Bo}$ differently from endogenous forms; women taking oral contraceptives have persistently raised $T_{Bo}$, similar to naturally cycling women in the luteal phase.[95] We suggest this is also true for $T_{Br}$. Only one participant was using the combined patch; note that $T_{Br}$ for this participant was generally lower (more consistent with non-luteal females and males). For each voxel in (**A**) and (**B**), data are plotted as mean ±SEM. Voxel numbering on x-axis refers to numbers shown in overlay (top right); voxels were grouped into 4 concentric U-shaped regions labelled 1–4 based on $T_{Br}$ variation. Note relatively high $T_{Br}$ in cerebral white matter ('Superficial 3') and thalamus, and low $T_{Br}$ in grey matter surrounding the sagittal sulcus ('Superficial 1').

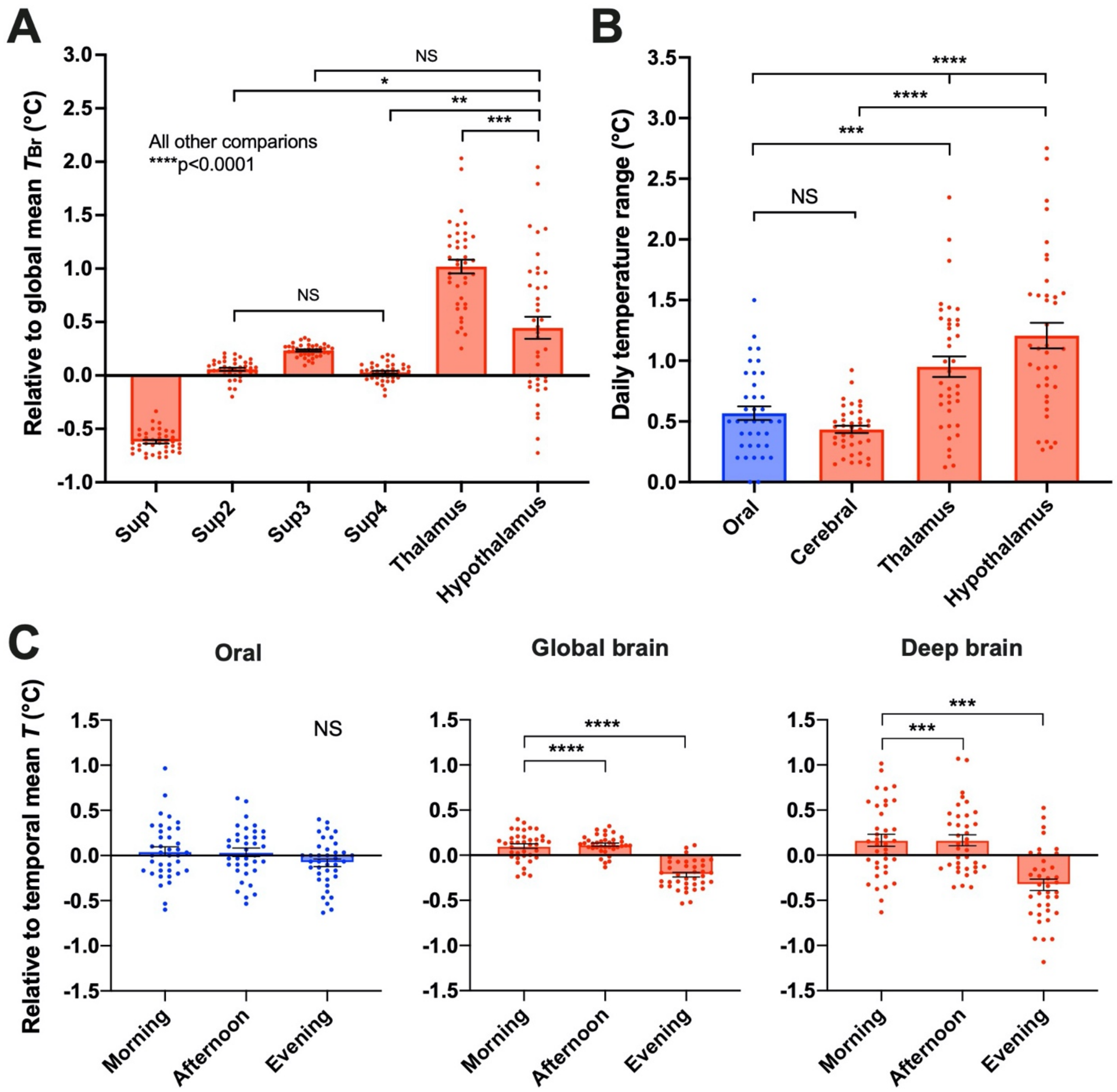


**Fig. S6. Daily temperature ranges by location.** (**A**) Time-averaged spatial variation in $T_{Br}$ by brain region; temperature deviations relative to global temporal mean $T_{Br}$ were calculated for each individual. Sup1-4, superficial cerebral layers 1-4. Comparisons between regions were significant unless specified as NS (n=39, repeated measures one-way ANOVA with Tukey's multiple comparisons test, *P=0.01, **P=0.004, ***P=0.0002, ****P<0.0001). (**B**) Daily temporal temperature ranges (maximum versus minimum across the three tested time points) are plotted by measurement site (n=39). Note that temperature varied more by time of day in the thalamus and hypothalamus than in the cerebrum or orally (repeated measures one-way ANOVA with Sidak's multiple comparisons test ***P=0.0004, ****P<0.0001). 'Cerebral' refers to mean $T_{Br}$ from all cerebral voxels combined. (**C**) Temporal variations in temperature; temperature deviations relative to temporal mean were calculated for each participant for three sites (body, global brain, deep brain) at each time point. Note drop in $T_{Br}$ in late evening (n=38, mixed-effects analysis (REML) with Tukey's multiple comparisons test, ****P<0.0001, ***P=0.0004 for morning versus evening and 0.0001 for afternoon versus evening). NS, non-significant. Note that analyses in (**A**) to (**C**) were performed without controlling for sex, age, menstrual cycle stage, or chronotype. Irrespectively, the effects of brain region and time of day on $T_{Br}$ remain significant.

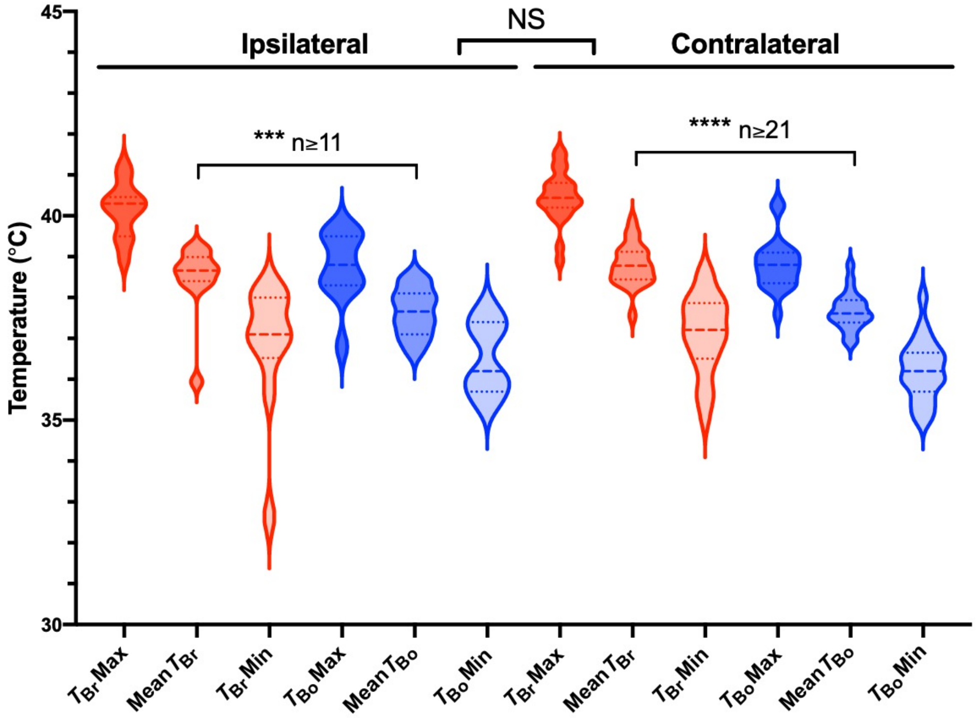


**Fig. S7. Patient temperatures according to probe site.** Violin plot of $T_{Br}$ and $T_{Bo}$ in TBI patients with focal brain injury. The nature of brain injury was available for 46 patients (34 focal, 12 diffuse); of the focal injury cases, the brain probe was placed ipsilateral and contralateral to the site of injury in 12 and 22 cases, respectively. Mean $T_{Br}$ was greater than mean $T_{Bo}$, regardless of site of probe placement (mixed effects analysis with Tukey's for multiple comparisons ***P=0.0007, ****P<0.0001, n refers to number of individual patients; one patient in each probe site group lacked $T_{Bo}$ data). There were no significant differences in temperature according to site of probe placement.

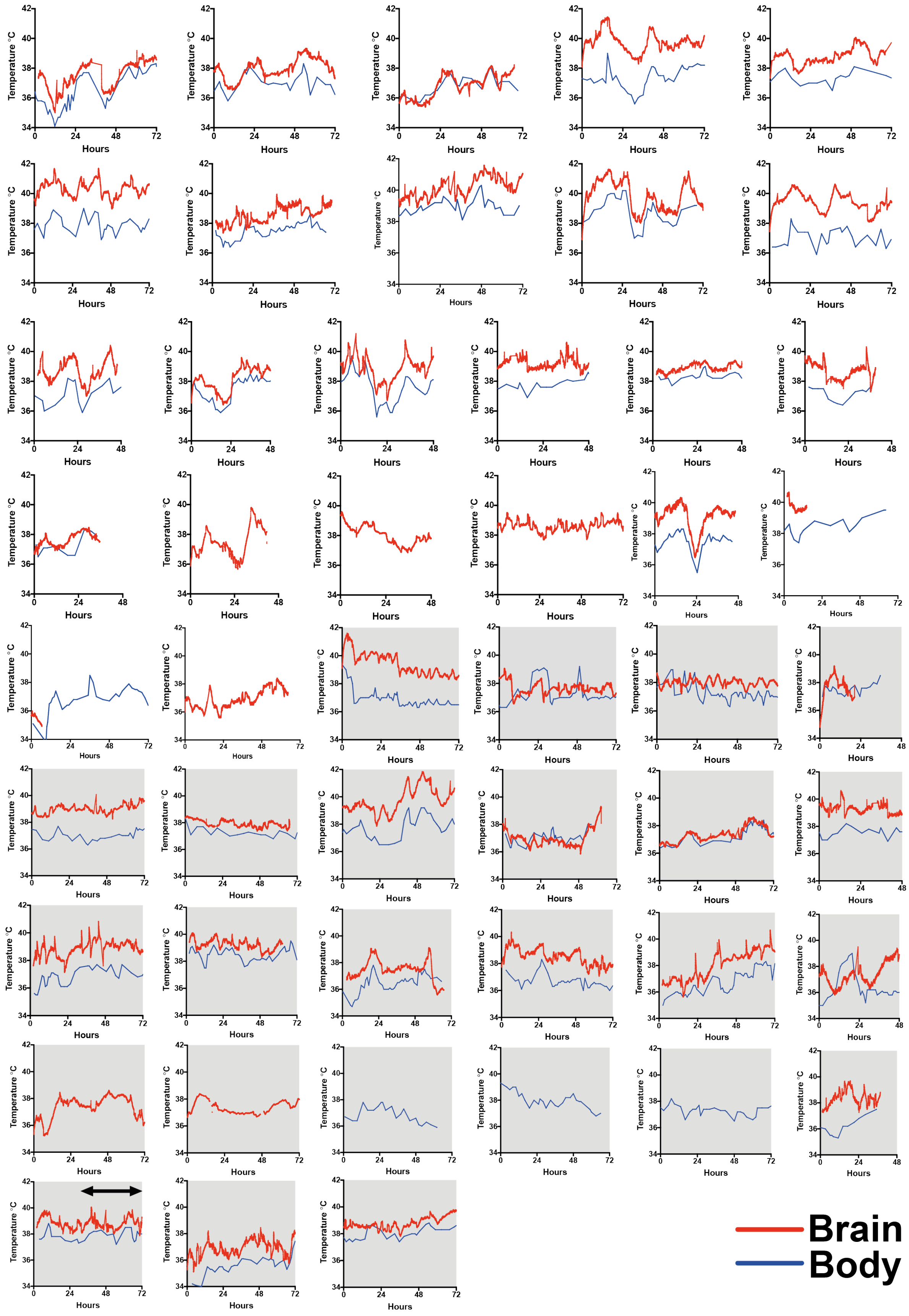
Temperature °C
Hours
Brain
Body

**Fig. S8. Raw temperatures in non-survivors and patients with daily rhythms.** Upper panel (white plots) show raw $T_{Br}$ and/or $T_{Bo}$ data from 24 patients that survived intensive care and were deemed to have evidence of a diurnal/daily/circadian rhythm in one or both temperature parameters at some point during recording. Lower panel (grey plots) show the 25 patients that did not survive in intensive care after TBI. All but one of these patients (lower left) showed a complete lack of a daily temperature rhythm during recording. Rhythmicity was apparent in some of these patients but it did not meet our assigned diurnal/circadian/daily cut-off for period length of 22–26h. Note that in the patients who died, transitory inversion of the $T_{Br}$–$T_{Bo}$ gradient was more commonly observed than in those that survived (in which $T_{Br}$ was almost always higher than $T_{Bo}$). The patient in the lower left corner was classified as 'diurnal/daily' based on the time period highlighted by the horizontal black arrow, although it is clear that the period length was >26h earlier on in the recording.

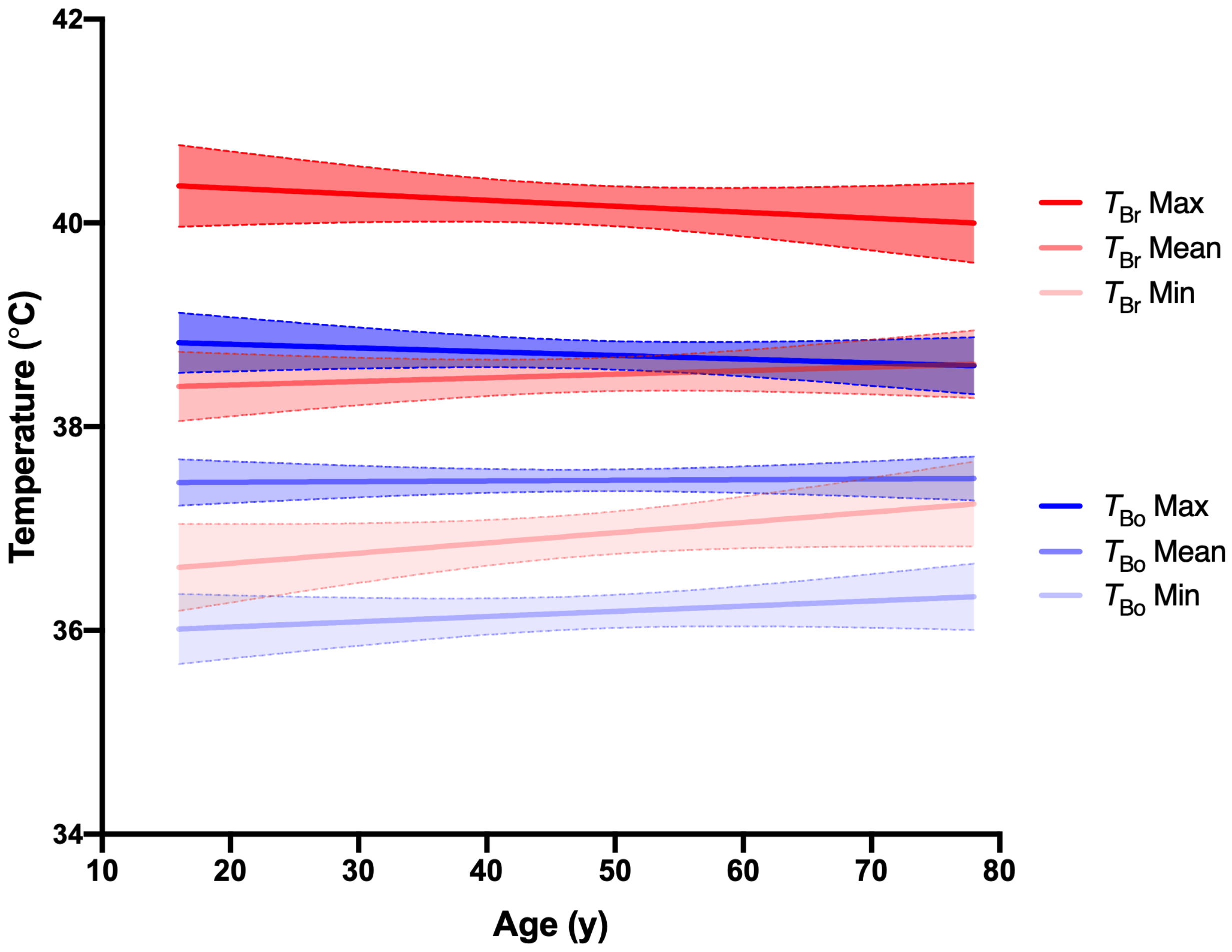


**Fig. S9. Minimum brain temperature trends upwards with age in TBI patients.** Linear regression of patient temperatures with age (n=105 for $T_{Br}$Mean, n=104 for $T_{Br}$Max and $T_{Br}$Min, n=101 for $T_{Bo}$Max and $T_{Bo}$Min, n=94 for $T_{Bo}$Mean); the apparent upward trend for minimum $T_{Br}$ was not significantly different from zero (slope 0.010, 95% confidence interval -0.002 to 0.022, P=0.096). Shaded areas represent 95% confidence intervals.

| Study | Subject age | Females/total | TOD control | $T_{Br}$ | Calibration method |
|---|---|---|---|---|---|
| Childs et al. (2007)[56] | 23–52y | 3/8 | No | 36.5 | Anaesthetised piglets[57] |
| Corbett et al. (1997)[57] | NR | 5/10 | No | 37.2/37.7* | Anaesthetised piglets[57] |
| Corbett & Laptook (1998)[96] | 22–47y | NR/10 | No | 37.0/36.6** | Anaesthetised piglets[57] |
| Curran et al. (2017)[97] | Mean 33y | NR/6 | No | NA | Phantom†[98] |
| Covaciu et al. (2010)[43] | 18–57y | 5/18 | No (10:30–17:30) | 38.1 | Phantom‡[42] |
| Covaciu et al. (2011)[98] | 21–62y | 2/10 | No | NA | Anaesthetised adult pigs[99] |
| Fujiwara et al. (2016)[100] | 21–50y | 3/15 | No | 38.3*** | Anaesthetised piglets[101] |
| Harris et al. (2008)[102] | 31–48y | 2/5 | No | NA | Phantom§[38] |
| Inoue et al. (2013)[103] | NR | NR/5 | No | 37.1 | NR |
| Kaupinnen et al. (2008)[104] | 21–51y | 5/14 | No | 37.4 | Anaesthetised piglets[101] |
| Kickhefel et al. (2010)[105] | 25–43y | 5/9 | No | NA | Ex vivo swine muscle |
| Marshall et al. (2006)[38] | 23–38y | 0/4 | No | 36.5 | Phantom referenced§[38] |
| Maudsley et al. (2017)[44] | 18–84y | 90/150 | No | 37.5 | Combination[101,46,106,107] |
| Onitsuka et al. (2018)[108] | Mean 26.9 | 0/8 | No | ~37.2 | Anaesthetised piglets[101] |
| Posporelis et al. (2018)[42] | Mean 23.1y | 7/20 | Yes (12:00–14:00) | NA | Anaesthetised piglets[101] |
| Rango et al. (2012)[39] | 49–78y | 5/10 | Yes (14:00–16:00) | 36.8 | Anaesthetised piglets[57] |
| Rango et al. (2014)[63] | Mean 43y | 6/14 | Yes (14:00–16:00) | 37.6 | Anaesthetised piglets[57] |
| Rango et al. (2015)[109] | 23–53y | 10/20 | Yes (14:00–16:00) | 37.38 | Anaesthetised piglets/rats[57,109] |
| Sharma et al. (2020)[110] | 23-46y | 7/18 | No | 37.2 | Combination[101,46,106,107] |
| Shiloh et al. (2008)[111] | Mean 30.6y | 0/10 | No | 37.7 | Anaesthetised piglets[101] |
| Sumida et al. (2016)[112] | 25–78y | 18/35 | Yes (17:00–19:00) | 36.04 | Anaesthetised piglets[101] |
| Thrippleton et al. (2014)[10] | 22–40y | 0/51 | Yes (afternoon) | 37.4 | Referenced phantom§[38] |
| Verius et al. (2019)[35] | 22–37y | 15/30 | No (08:00 to 15:50) | 37.78 | Phantom††[35] |
| Weis et al. (2012)[98] | 20–61y | 2/10 | No | NA | Anaesthetised adult pigs[113] |
| Zhang (2020)[114] | 19-49y | 5/10 | No | 36.9 | Combination[101,46,106,107] |

**Table S1. Published studies using MRS brain thermometry in healthy adults.** Subject age is reported as available in publication; either mean or range. Females/total states number of female subjects included relative to total number of subjects, where this information was available. NR, not reported; NA, not applicable; TOD, time of control. 'No' means there was no reported control for TOD when scanning subjects; time period during which scans were performed are given in brackets if reported. $T_{Br}$ = mean/median absolute $T_{Br}$ (if reported) in °C.
*frontal lobe/thalamus
**superficial cortex/thalamus
***defined as upper threshold in healthy control subjects
†Water and N-acteylaspartate (NAA) solution
‡Aqueous solutions of glycero-phosphocoline (GPC), creatine (Cr), and NAA
§Homogenous solution of metabolites
††pH-buffered aqueous solution of NAA, Cr, methyl protons of Cr (Cr2), dimethyl silapentane sulfonic acid (DSS), and sodium formate (NaFor)

| Criteria for inclusion | Criteria for exclusion |
|---|---|
| • Healthy men<br>• Healthy women with regular natural menstrual cycles for a minimum of 6 months, or women that have taken monophasic hormonal contraception (fixed dose oestradiol and synthetic progestin) for a minimum of 3 months prior to scanning and will continue to do so during the month of scanning.<br>• Age 20–40 years<br>• Body mass index 18.5–29.9<br>• Live within 5 mile radius of Edinburgh Imaging (Royal Infirmary of Edinburgh) Facility<br>• Capacity to understand written and verbal information provided in English, and able to provide valid written informed consent to participate<br>• Able to wear an actigraphy wristband for a week prior to scanning<br>• Able to commit to 3 x 45min scanning protocol within selected 24h period<br>• Able to commit to fixed times of food and caffeine consumption on the day of scanning<br>• Able to avoid alcohol and excessive physical activity on the day of scanning | • Pregnancy<br>• Early menopause, irregular menstrual cycles, premenstrual syndrome<br>• Lack of luteinizing hormone surge prior to two scheduled scanning days (naturally cycling women only)<br>• MRI contraindications (e.g. cardiac pacemaker, cochlear implant, claustrophobia, any prior accident in which metal penetrated one or both eyes)<br>• Oral temperature outside of normal range (33.2–38.1°C) prior to any scan<br>• Medical history that might limit activity or alter cerebral blood flow (hypothyroidism, stroke, severe arthritis, Parkinson's disease, dementia, history of brain trauma, brain tumour or epilepsy, significant mental illness besides clinical depression, spinal cord injuries, recent serious burn, diabetes, dehydration)<br>• Taking medications (except seasonal allergy medication, over-the-counter NSAIDs, or contraceptives), paracetamol, drug abuse<br>• Known neurodevelopmental, neuropsychiatric or neurodegenerative disorder<br>• Known sleep or chronotype disorder (delayed sleep phase syndrome, familial advanced sleep phase syndrome)<br>• Known family history of cardiovascular disease at < 40 years of age<br>• Failure to attend, or late attendance at, two independently scheduled morning scans<br>• Participant-reported ill-health on the day before or the morning of scanning |

**Table S2. Inclusion and exclusion criteria for prospective study**

| | NAA | | | $H_2O$ | | |
|---|---|---|---|---|---|---|
| | Morning | Afternoon | Evening | Morning | Afternoon | Evening |
| **Females** | | | | | | |
| Cerebrum | 7.44 (0.19) | 7.31 (0.16) | 7.31 (0.18) | 9.70 (0.22) | 9.45 (0.19) | 9.40 (0.20) |
| Thalamus | 8.64 (0.64) | 8.46 (0.78) | 8.46 (0.75) | 11.5 (0.98) | 11.3 (0.84) | 11.3 (0.73) |
| Hypothalamus | 10.8 (1.63) | 10.0 (0.67) | 9.86 (1.19) | 14.1 (1.42) | 14.0 (1.38) | 13.9 (1.39) |
| **Males** | | | | | | |
| Cerebrum | 7.42 (0.29) | 7.36 (0.27) | 7.35 (0.28) | 9.68 (0.34) | 9.57 (0.35) | 9.52 (0.31) |
| Thalamus | 8.65 (0.58) | 8.61 (0.62) | 8.44 (0.71) | 11.5 (0.92) | 11.7 (0.85) | 11.3 (0.69) |
| Hypothalamus | 10.4 (1.44) | 10.3 (1.57) | 9.83 (0.88) | 13.6 (1.48) | 13.7 (1.27) | 13.5 (1.48) |

**Table S3. MRS linewidths by brain region, time of day, and sex.** Spectral linewidth data presented for NAA and $H_2O$ as mean (SD) for n=19 females and n=20 males. All data for the female participant with an HRF was excluded.

**Movie S1. Introduction to HEATWAVE.** Dynamic 3D brain temperature map showing aggregate data from luteal females at one time point and highlighting brain regions from which data was extracted in healthy volunteers. Inferno colour scale is used to assign a temperature to each tissue voxel, to 0.1°C resolution. The colour of each voxel represents the mean raw temperature of that voxel calculated from the data obtained from all luteal females (n=14). For enhanced accessibility, the mapping reverses the radiological convention used in Fig.2A such that from all perspectives, the data presented on the right side of the brain in HEATWAVE are from the right side of the human brain. Note that the thalamus is illuminated as a relatively hot, core brain structure.

**Movie S2. HEATWAVE by sex.** Comparison of daily brain temperature variation in luteal females and males (aggregate data from each group), enabling both time-of-day and sex differences to be visualized in accelerated time, side-by-side. Two days' worth of data are presented in an accelerated looped fashion at 1 hour resolution. At the data collection points (9am, 4pm, 11pm), the colour of each voxel represents the mean raw temperature of that voxel calculated from the data obtained from either all luteal females (n=14), or all males (n=20). Between data collection points, data were interpolated via non-linear regression of the aggregate data in GraphPad Prism version 8.2, fitting to a sine function.

### Supplementary references

1. Fischer D, Lombardi DA, Marucci-Wellman H, Roenneberg T. Chronotypes in the US ± Influence of age and sex. *PLoS ONE* 2017;12:e0178782
2. Cambras T, Castro-Marrero J, Zaragoza MC, Diez-Noguera A, Alegre J. Circadian rhythm abnormalities and autonomic dysfunction in patients with Chronic Fatigue Syndrome/Myalgic Encephalomyelitis. *PLoS ONE* 2018;13:e0198106. doi:10.1371/journal.pone.0198106
3. Blackwell T, Redline S, Ancoli-Israel S et al. Comparison of sleep parameters from actigraphy and polysomnography in older women: the SOF study. *Sleep* 2008;31:283-291. doi:10.1093/sleep/31.2.283
4. Marino M, Rueschman MN, Winkelman JW et al. Measuring sleep: accuracy, sensitivity and specificity of wrist actigraphy compared to polysomnography. *Sleep* 2013;36:1747-1755. doi:10.5665/sleep.3142
5. Albu S, Umemura G, Forner-Cordero A. Actigraphy-based evaluation of sleep quality and physical activity

in individuals with spinal cord injury. *Spinal Cord Series and Cases* 2019; 5:7. doi:10.1038/s41394-019-0149-0
6. Cole RJ, Kripke DF, Gruen W, Mullaney DJ, Gillin JC. Automatic sleep/wake identification from wrist activity. *Sleep* 1992;15:461-469
7. Roenneberg T, Pilz LK, Zerbini G, Winnebeck EC. Chronotype and Social Jetlag: A (Self-) Critical Review. *Biology* 2019;8:54. doi:10.3390/biology8030054
8. Beale AD, Pedrazzoli M, Goncalves BdSB et al. Comparison between an African town and a neighbouring village shows delayed, but not decreased, sleep during the early stages of urbanisation. *Sci Rep* 2017;7:5697. DOI:10.1038/s41598-017-05712-3
9. Jankowski KS. Social jet lag: Sleep-corrected formula. *Chronobiol Int* 2017;34:531-535. doi: 10.1080/07420528.2017.1299162
10. Thrippleton MJ, Parikh J, Harris B et al. Reliability of MRSI brain temperature mapping at 1.5 and 3 T. *NMR Biomed* 2014;27:183-90
11. Scheenen TWJ, Klomp DWJ, Wijnen JP, Heerschap A. Short echo time H-1-MRSI of the human brain at 3T with minimal chemical shift displacement errors using adiabatic refocusing pulses. *Magn Reson Med* 2008;59:1-6
12. Wilson M, Andronesi O, Barker PB et al. Methodological consensus on proton MRS of the brain: Review and recommendations. *Magn Resn Med* 2019;82:527-550. doi: 10.1002/mrm.27742
13. Steyerberg EW, Mushkudiani N, Perel P et al. Predicting outcome after traumatic brain injury: development and international validation of prognostic scores based on admission characteristics. *PLoS Med* 2008;5:e165. doi:10.1371/journal.pmed.0050165
14. Logan RW, McClung CA. Rhythms of life: circadian disruption and brain disorders across the lifespan. *Nat Rev Neurosci* 2019;20:49-65
15. Kondratova AA, Kondratov RV. The circadian clock and pathology of the ageing brain. *Nat Rev Neurosci* 2012;13:325-335
16. Roenneberg T, Merrow M. The circadian clock and human health. *Curr Biol* 2016; 26:R432-443
17. Nedergaard M, Goldman SA. Glymphatic failure as a final common pathway to dementia. *Science* 2020;370:50-56
18. GBD 2016 Neurology Collaborators. Global, regional, and national burden of neurological disorders, 1990–2016: a systematic analysis for the Global Burden of Disease Study 2016. *Lancet Neurol* 2019;18:459-480
19. Bekinschtein TA, Golombek DA, Simonetta SH, Coleman MR, Manes FF. Circadian rhythms in the vegetative state. *Brain Inj* 2009;23:915-919
20. C. Kirkness J, Burr RL, Thompson HJ, Mitchell PH. Temperature rhythm in aneurysmal subarachnoid haemorrhage. *Neurocrit Care* 2008;8:380-390
21. Paul T, Lemmer B. Disturbance of circadian rhythms in analgosedated intensive care unit patients with and without craniocerebral injury. *Chronobiol Int* 2007;24:45-61
22. Takekawa H, Miyamoto M, Miyamoto T, Yokota N, Hirata K. Alteration of circadian periodicity in core body temperature of patients with acute stroke. *Psychiatry Clin Neurosci* 2002;56:221-222
23. Lyall LM, Wyse CA, Graham N et al. Association of disrupted circadian rhythmicity with mood disorders, subjective wellbeing, and cognitive function: a cross-sectional study of 91 105 participants from the UK Biobank. *Lancet Psychiatry* 2018;5:507-514
24. Bunney BG, Walsh DM, Stein R et al. Circadian dysregulation of clock genes: clues to rapid treatments in major depressive disorder. *Mol Psychiatry* 2015;20:48-55
25. Leng Y, Musiek ES, Hu K, Cappuccio FP, Yaffe K. Association between circadian rhythms and neurodegenerative diseases. *Lancet Neurol* 2019;18:307-318
26. Khan S, Nobili L, Khatami R, Loddenkemper T et al. Circadian rhythm and epilepsy. *Lancet Neurol* 2018;17:1098-1108
27. Nikitopoulou G, Crammer JL. Change in diurnal temperature rhythm in manic-depressive illness. *BMJ* 1976;1:1311-1314
28. Busto R, Dietrich WD, Globus MY, Valdes I, Scheinberg P, Ginsberg MD. Small differences in intraischemic brain temperature critically determine the extent of ischemic neuronal injury. *J Cereb Blood Flow Metab* 1987;7:729-738
29. Childs C, Vail A, Protheroe R, King AT, Dark PM. Differences between brain and rectal temperatures during routine critical care of patients with severe traumatic brain injury. *Anaesthesia* 2005;60:759-76

30. Abu-Arafeh A, Rodriguez A, Paterson RL, Andrews PJD. Temperature variability in a modern targeted temperature management trial. *Crit Care Med* 2018;46:223-228
31. Dehkhargani S, Fleischer CC, Qiu D, Yepes M, Tong F. Cerebral temperature dysregulation: MR thermographic monitoring in a nonhuman primate study of acute ischemic stroke. *AJNR Am J Neuroradiol* 2017;38:712-720
32. Rumana CS, Gopinath SP, Uzara M, Valadka AB, Robertson CS. Brain temperature exceeds systemic temperature in head-injured patients. *Crit Care Med* 1998;26:562-567
33. Karaszewski B, Wardlaw JM, Marshall I et al. Early brain temperature elevation and anaerobic metabolism in human acute ischaemic stroke. *Brain* 2009;132:955-964
34. Karaszewski B, Carpenter TK, Thomas RGR et al. Relationships between brain and body temperature, clinical and imaging outcomes after ischemic stroke. *J Cereb Blood Flow Metab* 2013;33:1083-1089
35. Verius M, Frank F, Gizewski E, Broessner G. Magnetic resonance spectroscopy thermometry at 3 Tesla: importance of calibration measurements. *Ther Hypothermia Temp Manag* 2019;9: 146-155
36. Nanba T, Nishimoto H, Yoshioka Y et al. Apparent brain temperature imaging with multi-voxel proton magnetic resonance spectroscopy compared with cerebral blood flow and metabolism imaging on positron emission tomography in patients with unilateral chronic major artery steno-occlusive disease. *Neuroradiology* 2017;59:923-935
37. Tsutsui S, Nanba T, Yoshioka Y et al. Preoperative brain temperature imaging on proton magnetic resonance spectroscopy predicts hemispheric ischemia during carotid endarterectomy for unilateral carotid stenosis with inadequate collateral blood flow. *Neurol Res* 2018;40:617-623
38. Marshall I, Karaszewski B, Wardlaw JM et al. Measurement of regional brain temperature using proton spectroscopic imaging: validation and application to acute ischemic stroke. *Magn Reson Imaging* 2006;24:699-706
39. Rango M, Arighi A, Bonifati C, Del Bo R, Comi G, Bresolin N. The brain is hypothermic in patients with mitochondrial diseases. *J Cereb Blood Flow Metab* 2014;34: 915-920
40. Sone D, Ikegaya N, Takahashi A et al. Noninvasive detection of focal brain hyperthermia related to continuous epileptic activities using proton MR spectroscopy. *Epilepsy Res* 2017; 138:1-4
41. Shiloh R, Kushnir T, Gilat Y et al. In vivo occipital-frontal temperature-gradient in schizophrenia patients and its possible association with psychopathology: a magnetic resonance spectroscopy study. *Eur Neuropsychopharmacol* 2008;18:557-564
42. Posporelis S, Coughlin JM, Marsman A et al. Decoupling of brain temperature and glutamate in recent onset of schizophrenia: a 7T proton Magnetic Resonance Spectroscopy study. *Biol Psychiatry Cogn Neurosci Neuroimaging* 2018;3:248-254
43. Covaciu L, Rubertsson S, Ortiz-Nieto F, Ahlström H, Weis J. Human brain MR spectroscopy thermometry using metabolite aqueous-solution calibrations. *J Magn Reson Imaging* 2010;31:807-814
44. Maudsley AA, Goryawala MZ, Sheriff S. Effects of tissue susceptibility on brain temperature mapping. *Neuroimage* 2017;146:1093-1101
45. Chadzynski GL, Bender B, Groeger A, Erb M, Klose U. Tissue specific resonance frequencies of water and metabolites within the human brain. *J Magn Reson* 2011;212:55-63
46. Brown MA. Time-domain combination of MR spectroscopy data acquired using phased-array coils. *Magn Reson Med* 2004;52:1207-1213
47. Maloney SK, Mitchell D, Mitchell G, Fuller A. Absence of selective brain cooling in unrestrained baboons exposed to heat. *Am J Physiol Regul Integr Comp Physiol* 2007;292: R2059-2067
48. Hayward JN, Baker MA. Role of cerebral arterial blood in the regulation of brain temperature in the monkey. *Am J Physiol* 1968;215:389-403
49. Prakash KN, Verma SK, Marchenko Y et al. Echo planar spectroscopic imaging based temperature calibration at 7T and 3T for whole brain temperature measurement in rodents and humans. *International Society of Magnetic Resonance in Medicine, Milan 2014* (conference paper)
50. Fuller CA, Baker MA. Selective regulation of brain and body temperatures in the squirrel monkey. *Am J Physiol* 1983;245:R293-R297
51. Baker FC, Waner JI, Vieira EF, Taylor SR, Driver HS, Mitchell D. Sleep and 24 hour body temperatures: a comparison in young men, naturally cycling women and women taking hormonal contraceptives. *J Physiol* 2001;530:565-574
52. Papaionnou VE, Chouvarda IG, Maglaveras NK, Pneumatikos IA. Temperature variability analysis using wavelets and multiscale entropy in patients with systemic inflammatory response syndrome, sepsis, and septic shock. *Crit Care Lond Engl* 2012;16:R51

53. Mariak Z. Intracranial temperature recordings in human subjects. The contribution of the neurosurgeon to thermal physiology. *J Therm Biol* 2002; 27:219-228. https://doi.org/10.1016/S0306-4565(01)00087-0.
54. Walsh JJ, Huang Y, Simmons JW et al. Dynamic thermal mapping of localized therapeutic hypothermia in the brain. *J Neurotrauma* 2020;37:55-65
55. Erikson R. Oral temperature differences in relation to thermometer and technique. *Nurs Res* 1980;29:157-64
56. Childs C, Hiltunen Y, Vidyasagar R, Kaupinnen RA. Determination of regional brain temperature using proton magnetic resonance spectroscopy to assess brain-body temperature differences in healthy human subjects. *Magn Reson Med* 2007;57:59-66
57. Corbett R, Laptook A, Weatherall P. Noninvasive measurements of human brain temperature using volume-localized proton magnetic resonance spectroscopy. *J Cereb Blood Flow Metab* 1997;17:363-369
58. Shiraki K, Sagawa S, Tajima F, Yokota A, Hashimoto M, Brengelmann GL. Independence of brain and tympanic temperatures in an unanaesthetised human. *J Appl Physiol (1985)* 1988;65:482-486
59. Awaya S. A study of brain temperature in patients with severe head injuries. *Nihon Ika Daigaku Zasshi* 1993;60:37-43
60. Brengelmann GL. Specialised brain cooling in humans? *FASEB J* 1993;7:1148-1153
61. Cabanac M. Selective brain cooling in humans: “fancy” or fact? *FASEB J* 1993;7:1143-1147
62. Refinetti R. The circadian rhythm of body temperature. *Front Biosci (Landmark Ed)* 2010;15:564-594
63. Rango M, Arighi A. Bonifati C, Del Bo R, Comi G, Bresolin N. The brain is hypothermic in patients with mitochondrial diseases. *J Cereb Blood Flow Metab* 2014;34:915-920
64. Orita T, Izumihara A, Tsurutani T, Kajiwara K. Brain temperature before and after brain death. *Neurol Res* 1995;17:443-444
65. Sen A, Jette N, Husain M, Sander JW. Epilepsy in older people. *Lancet* 2020;395:735-748
66. Peretti D, Bastide A, Radford H et al. RBM3 mediates structural plasticity and protective effects of cooling in neurodegeneration. *Nature* 2015;518:236-239
67. Sandsmark DK, Bashir A, Wellington CL, Diaz-Arrastia R. Cerebral microvascular injury: a potentially treatable endophenotype of traumatic brain injury-induced neurodegeneration. *Neuron* 2019;103:367-379
68. Rzechorzek NM, Connick P, Livesey MR et al. Hypothermic preconditioning reverses tau ontogenesis in human cortical neurons and is mimicked by protein phosphatase 2A inhibition. *EBioMedicine* 2015;3:141-154
69. Spillantini MG, Goedert M. Tau pathology and neurodegeneration. *Lancet Neurol* 2013;12: 609-622
70. Mirmiran M, Bernardo L, Jenkins SL, Ma XH, Brenna JT, Nathanielsz PW. Growth, neurobehavioural and circadian rhythm development in newborn baboons. *Pediatr Res* 2001; 49:673-677
71. Tjijs RD, Surges R, O’Brien TJ, Sander W. Epilepsy in adults. *Lancet* 2019;393:689-701
72. Arendt T, Stieler J, Strijkstra AM et al. Reversible paired helical filament-like phosphorylation of tau is an adaptive process associated with neuronal plasticity in hibernating animals. *J Neurosci* 2003;23:6972-6981
73. Mariak Z, White MD, Lewko J, Lyson T, Piekarski P. Direct cooling of the human brain by heat loss from the upper respiratory tract. *J Appl Physiol (1985)* 1999;87:1609-1613
74. Roberge RJ, Kim JH, Coca A. Protective facemask impact on human thermoregulation: an overview. *Ann Occup Hyg* 2012;56:102-112
75. Kuwabara N, Seki K, Aoki K. Circadian, sleep and brain temperature rhythms in cats under sustained daily light-dark cycles and constant darkness. *Physiol Behav* 1986;38,283-289
76. J. Aschoff, Circadian rhythms in man. *Science* 1965;148:1427-1432
77. Daan S, Honma S, Honma K. Body temperature predicts the direction of internal desynchronization in humans isolated from time cues. *J Biol Rhythms* 2013;28:403-411
78. Archer SN, Oster H. How sleep and wakefulness influence circadian rhythmicity: effects of insufficient and mistimed sleep on the animal and human transcriptome. *J Sleep Res* 2015;24: 476-493
79. Dijk DJ. Circadian variation of EEG power spectra in NREM and REM in humans: dissociation from body temperature. *J Sleep Res* 1999;8:198-195
80. Baker FC, Angara C, Szymusiak R, McGinty D. Persistence of sleep-temperature after suprachiasmatic nuclei lesions in rats. *Am J Physiol Integr Comp Physiol* 2005;289:R927-838
81. V. Skop V, Guo J, Liu N et al. Mouse thermoregulation: introducing the concept of the thermoneutral point. *Cell Rep* 2020;31:107501. doi: 10.1016/j.celrep.2020.03.065
82. Frank MG. Circadian regulation of synaptic plasticity. *Biology (Basel)* 2016;5:pii:E31 doi: 10.3390/biology5030031
83. Deboer T. Brain temperature dependent changes in the electroencephalogram power spectrum of humans

and animals. *J Sleep Res* 1998;7:254-262
84. Cai X, Qiao J, Kulkarni P, Harding IC, Ebong E, Ferris CF. Imaging the effect of the circadian light-dark cycle on the glymphatic system in awake rats. *Proc Natl Acad Sci USA* 2020;117:668-676
85. Hastings MH, O'Neill JS, Maywood ES. Circadian clocks: regulators of endocrine and metabolic rhythms. *J Endocrinol* 2007;195:187-98
86. Conroy DA, Spielman AJ, Scott RQ. Daily rhythm of cerebral blood flow velocity. *J Circadian Rhythms* 2005;3:3. doi:10.1186/1740-3391-3-3
87. Aschoff VJ, Wever R. Spontanperiodik des menschen bei ausschluß aller zeitgeber. *Naturwissenschaften* 1962;49:337-342
88. Harding EC, Franks NP, Wisden W. The temperature dependence of sleep. *Front Neurosci* 2019;23:336.
89. Raymann RJ, Swaab DF, Van Someren EJ. Cutaneous warming promotes sleep onset. *Am J Physiol Regul Integr Comp Physiol* 2005;288:R1589-R1597
90. Cook JS, Sauder CL, Ray CA. Melatonin differentially affects vascular blood flow in humans. *Am J Physiol Heart Circ Physiol* 2011;300:H670-H674
91. Maas MB, Lizza BD, Abbott AM. Factors disrupting melatonin secretion rhythms during critical illness. *Crit Care Med* 2020;48:854-861
92. Blume C, Lechinger J, Santhi N et al. Significance of circadian rhythms in severely brain-injured patients: a clue to consciousness? *Neurology* 2017;88:1933-1941
93. Culver A, Coiffard B, Antonini F et al. Circadian disruption of core body temperature in trauma patients: a single-center retrospective observational study. *J Intensive Care* 2020;8:4. doi: 10.1186/s40560-019-0425-x
94. Van Someren EJ. More than a marker: interaction between the circadian regulation of temperature and sleep, age-related changes, and treatment possibilities. *Chronobiol Int* 2000;17:313-354. doi: 10.1081/cbi-100101050
95. Yablonskiy DA, Ackerman JJ, Raichle ME. Coupling between changes in human brain temperature and oxidative metabolism during prolonged visual stimulation. *Proc Natl Acad Sci USA* 2000;97:7603-7608.
96. Corbett RJ, Laptook AR. Failure of localized head cooling to reduce brain temperature in adult humans. *Neuroreport* 1998;9:2721-2725
97. Curran EJ, Wolfson DL, Watts R, Freeman K. Cold blooded: evaluating brain temperature by MRI during surface cooling of human subjects. *Neurocrit Care* 2017;27:214-219
98. Covaciu L. Brain temperature in volunteers subjected to intranasal cooling. *Intensive Care Med* 2011;37:1277-1284
99. Weis J, Covaciu L, Rubertsson S, Allers M, Lunderquist A, Ahlstrom et al. Noninvasive monitoring of brain temperature during mild hypothermia. *Magn Reson Imaging* 2009;27:923-932
100. Fujiwara S, Yoshika Y, Matsuda T et al. Brain temperature measured by 1H-magnetic resonance spectroscopy in acute and subacute carbon monoxide poisoning. *Neuroradiology* 2016;58:27-32
101. Cady EB, D'Souza PC, Penrice J, Lorek A. The estimation of local brain temperature by in vivo 1H magnetic resonance spectroscopy. *Magn Reson Med* 1995;33:862-867
102. Harris BA, Andrews PJ, Marshall I, Robinson TM, Murray GD. Forced convective head cooling device reduces human cross-sectional brain temperature measured by magnetic resonance: a non-randomized healthy volunteer pilot study. *Br J Anaesth* 2008;100:365-72
103. Inoue T, Shimizu H, Fujimura M et al. Noninvasive measurement of human brain temperature adjacent to arteriovenous malformation using 3.0T magnetic resonance spectroscopy. *Clin Neurol Neurosurg* 2013;115:445-449
104. Birkl C, Langkammer C, Haybaeck J et al. Temperature dependency of T1 relaxation time in unfixed and fixed human brain tissue. *Biomed Tech (Berl)* 2013;58 Suppl 1.pii/j/bmte.2013.58.issue-s1-L/bmt-2013-4290/bmt-2013-4290.xml. doi: 10.1515/bmt-2013-4290
105. Kickhefel A, Roland J, Weiss C, Schick F. Accuracy of real-time MR temperature mapping in the brain: a comparison of fast sequences. *Phys Med* 2010;26:192-201
106. Cady EB, Penrice J, Robertson NJ. Improved reproducibility of MRS regional brain thermometry by 'amplitude-weighted combination'. *NMR Biomed* 2011;24:865-872
107. Zhu M, Bashir A, Ackerman JJ, Yablonskiy DA. Improved calibration technique for in vivo proton MRS thermometry for brain temperature measurement. *Magn Reson Med* 2008;60:536-541
108. Onitsuka S, Nakamura D, Onishi T, Arimitsu T, Takahashi H, Hasegawa H. Ice slurry ingestion reduces human brain temperature measured using non-invasive magnetic resonance spectroscopy. *Sci Rep* 2018;8:2757. doi: 10.1038/s41598-018-21086-6

109. Karoly PJ, Goldenholz DM, Freestone DR et al. Circadian and circaseptan rhythms in human epilepsy: a retrospective cohort study. *Lancet Neurol* 2018;17:977-985
110. Sharma AA, Nenert R, Mueller C, Maudsley AA, Younger JW, Szaflarski JP. Repeatability and reproducibility of in-vivo brain temperature measurements. *Front Hum Neurosci* 2020;14:598435. doi: 10.3389/fnhum.2020.598435
111. Trefler A, Sadeghi N, Thomas AG, Pierpaoli C, Baker CI, Thomas C. Impact of time-of-day on brain morphometric measures derived from T1-weighted magnetic resonance imaging. *Neuroimage* 2016;133:41-52
112. Simon E. Tympanic temperature is not suited to indicate selective brain cooling in humans: a re-evaluation of the thermophysiological basics. *Eur J Appl Physiol* 2007;101:19-30
113. Weis J, Covaciu L, Rubertsson S et al. Phase-difference and spectroscopic imaging for monitoring of human brain temperature during cooling. *Magn Reson Imaging* 2012;30:1505-1511
114. Zhang Y, Taub E, Mueller C, Younger J, Uswatte G, DeRamus TP, Knight DC. Reproducibility of whole-brain temperature mapping and metabolite quantification using proton magnetic spectroscopy. *NMR Biomed* 2020;33:e4313. https://doi.org/10.1002/nbm.4313

### Supplementary Appendices

Appendix 1 - Participant Information Sheet and Consent to Participate Form
Appendix 2 - Prospective Study Protocol
Appendix 3 - MRI Protocol
Appendix 4 - Study Participant Data Form
Appendix 5 - Analytic code for statistical models
