## Supplementary Appendix 1 for "Diurnal brain temperature rhythms and mortality after brain injury: a prospective and retrospective cohort study"

### Participant Information Sheet

Diurnal brain temperature mapping by Magnetic Resonance Spectroscopy (MRS) in healthy volunteers

**You are being invited to take part in a research study. Before you decide whether or not to take part, it is important for you to understand why the research is being done and what it will involve. Please take time to read the following information carefully. Talk to others about the study if you wish. Contact us if there is anything that is not clear, or if you would like more information. Take time to decide whether or not you wish to take part.**

#### What is the purpose of the study?

##### Background and rationale

Circadian (daily) rhythms co-ordinate many biological functions. These rhythms are driven by molecular 'clocks' that exist in every cell of your body. Good timekeeping in brain cell clocks is essential for brain health and for maintaining body temperature. Research suggests that brain cell clocks fail to keep time in neurodegenerative diseases such as dementia. Risk of these diseases increases with age, and elderly people struggle to regulate their own body temperature.

In healthy people, body temperature varies with a daily rhythm, but we do not know whether brain temperature also varies in this way. Previous studies suggest that brain temperature is higher than body temperature, but we do not know if this difference persists throughout the day. Body temperature is higher during some parts of the menstrual cycle in women, relative to men, but we do not know if this is the case for brain temperature. There is some evidence that hormones in contraceptive pills are associated with an increased body temperature throughout the menstrual cycle, but whether this affects brain temperature is unknown.

Brain temperature can be measured using a tiny thermometer inserted into the surface of the brain. This invasive method can only be justified in critically ill patients, and it measures only a tiny part of the brain. MRS uses a magnetic resonance imaging (MRI) scanner and is currently the only way to measure human brain temperature non-invasively. A recent study at Edinburgh Imaging showed that MRS can measure temperature in many brain regions in healthy male volunteers.

##### What is the purpose of this study?

Our main objective is to determine how human brain temperature varies according to time of day. To achieve this we will measure brain temperature in healthy volunteers at three different time points in the 24-hour cycle using MRS. **This study will provide critical information about daily variation in human brain temperature in healthy adults.**

Our study may also determine:

- How much brain temperature varies in the course of a day relative to oral temperature
- Whether there is a difference in temperature between different parts of the brain
- Whether there is a difference in brain temperature between healthy men and women

**How many healthy volunteers will be involved?**

36 volunteers (18 men and 18 women) are needed to answer our main research question. Four ‘extra’ volunteers will be recruited for scanning. This will account for potential losses due to participant-elected withdrawal from the study, unexpected technical failure of the scanner, and exclusion of participants after scanning (see below).

**What areas are being studied?**

We will use a non-invasive method called Magnetic Resonance Spectroscopy (MRS) to scan your brain. MRS can be used to estimate temperature in different parts of your brain. Because MRS is based on MRI, we will also obtain limited information about the structure of your brain.

**Where will the study be conducted?**

Scanning will take place at the Edinburgh Imaging (Royal Infirmary of Edinburgh) Facility, Little France. Scans will be conducted using an MRI scanner in a temperature-controlled room. Most image processing and analysis will occur on-site. Some secondary analysis of anonymised brain temperature data will take place at the MRC Laboratory of Molecular Biology in Cambridge. Anonymised data cannot be linked back to you as an individual.

**When will the study start and end?**

We started recruiting volunteers in Spring 2019. Scanning will take place between June and September 2019. We expect to submit the results of the study for publication in Spring 2020.

**What will it mean to take part?**

Taking part in this study requires that you meet the criteria for inclusion, that you understand the information provided, and that you have the capacity to provide valid, written, informed consent to participate.

Overall, your involvement will likely span 14-17 days. This includes today’s consenting interview, a 7-10 day period during which you will wear a special wristband, one day of scanning, and up to 7 days after scanning. During the week after scanning, you must report any unexpected abnormal events to the Chief Investigator by email (even if these are completely unrelated to the scans). The total duration of your involvement could be longer (up to a maximum of 2 months) if we need to reschedule your scanning day.

#### Why have I been invited to take part?

You are a healthy volunteer for a study investigating the normal daily variation in human brain temperature.

#### Do I have to take part?

No, it is up to you to decide whether or not to take part. If you do decide to take part you will be given this information sheet to keep and you will be asked to sign a 'Consent to Participate' Form. If you decide to take part you are still free to withdraw at any time and without giving a reason. Deciding not to take part or withdrawing from the study will not affect the healthcare that you receive, or your legal rights. Before participating you should consider if this will affect any insurance you have and seek advice if necessary

#### What will happen if I take part?

Today the Chief Investigator will go through this Information Sheet and the Consent to Participate Form with you to check you have understood the information provided, and that you remain willing to take part. You will be invited to initial each statement on the Form and then sign it to give your consent to take part. You will then be given a special wristband and asked to wear this until your scanning date. The wristband will monitor your rest, sleep, and activity patterns, as well as your skin temperature and light exposure. If you decide to take part in the scanning, the data collected by the wristband will be downloaded via USB connector to a secure University of Edinburgh computer for processing. If you decide not to take part in the scanning, your wristband data will be safely destroyed without further processing.

In 7-10 days' time, you will be scheduled to attend the Edinburgh Imaging (Royal Infirmary of Edinburgh) Facility in the morning. The Chief Investigator will remove your wristband and then measure your height, weight, and oral temperature (under your tongue). A Radiographer (scanning specialist) will then **check that it is safe for you to be scanned**. A changing cubicle will be provided. You will be asked to place any metal objects (keys, phones, credit cards) in a locker. **Please do not wear any make-up or talcum powder and be prepared to remove contact lenses if you use them.** You will be asked to change into clean hospital clothing and/or a hospital gown (provided) before entering the scanning room.

You will be positioned in the scanner for your first scan in the morning, and asked to return 15 minutes in advance of your second and third scans in the afternoon and late evening, respectively on the same day. **It is important that you are not late for your scans, which will each last around 45 minutes.** The scanner makes quite loud noises whilst it operates. You will be provided with headphones during scanning. If at any stage during a scan you become worried, or wish to ask a question, you will be able to speak to the Radiographer via an intercom.

During each scan, your whole brain will be imaged using standard MRI to collect information about the structure of your brain. An MRS scan will then be run across a section of your brain at its surface, and again in a deeper region, to collect information on brain temperature.

**It is not essential to remain at the hospital between scans, but it may be convenient for you to do so.**

No adverse effects are expected as a result of scanning, however you must report to us any abnormalities experienced during the week before, and the week after, scanning. **We cannot provide any information to you about the results of your scans on the day of scanning.** Data from the wristband will be used to determine your normal rest/activity pattern so that we can effectively interpret your brain temperature data. **To safeguard your anonymity when publishing our findings, we cannot provide you with your individual wristband data** (although commercially available products can provide you with very similar data). No adverse effects are expected as a result of wearing the wristband. Although extremely unlikely, as with any new wearable item, an allergic reaction is possible; **you should contact your GP and the Chief Investigator immediately if you notice any itching or irritation in the contact area of the wristband with the skin.**

**Screening and exclusion**

Your eligibility to take part will be confirmed on the day of scanning. Any of the following would mean that you <u>cannot take part</u>:

- The wristband was not worn for two or more days preceding the scanning day
- Your body mass index (BMI) sits above or below the range stated in the inclusion criteria
- Your oral temperature sits above or below the normal range before the first scan (we will invite you to reschedule your scanning day).
- You report feeling unwell on the day before, or the morning of, scanning
- You are a woman with natural menstrual cycles, and you cannot confirm ovulation in the preceding 7-10 days (we will invite you to reschedule your scanning day for the following month).

The following circumstances would result in <u>exclusion of your data from our study</u>:

- Your oral temperature sits above or below the normal range before your second or third scans
- Detection of a significant health-related finding (HRF) in your brain scans
- Consumption of food or caffeine during prohibited time periods on the day of scanning
- Consumption of alcohol on the day of scanning
- Participation in excessive physical activity on the day of scanning
- Failure to attend, or late attendance at two independently scheduled morning scans

If you or your data are excluded, you will still be reimbursed for any expenses incurred as a result of taking part in our study.

**Health-related findings (HRFs)**

When apparently healthy volunteers take part in an imaging study, there is always a small chance that an abnormality (unrelated to the purpose of the study) may be found. For standard MRI scans of the brain, **this is occurs in roughly 1 in 20 people**. Some abnormalities have no known health implications; such findings are considered harmless in medical terms, but knowing about them may harm an individual by causing unnecessary anxiety. **Potentially serious HRFs are those that indicate the possibility of a health condition which, if confirmed, would likely impact upon lifespan, major body functions, or life quality.** For standard MRI scans of the brain, potentially serious HRFs are **noted in around 1 in 70 people**. However, based on current literature, serious diagnoses are confirmed only in 1 in 350 people.

Your scans will be reviewed by an expert in brain imaging. **If any significant HRFs are noted, these will be reported back to you and your GP in strictest confidence. A significant HRF is defined as one that could have health implications for you, and where the potential benefits of knowing about this HRF outweigh any potential harm caused to you.** Please note that diagnostic MRI studies include the injection of a chemical into the bloodstream to highlight some types of brain abnormality. **No injections will be administered during this study, and so some types of brain abnormality will not be detected.** Only data obtained from structurally normal brains can be included in our brain temperature analysis. The Chief Investigator will be notified if your data must be excluded on the basis of an HRF, but they will not be given any specific information about the reasons for exclusion.

**Will my GP be involved?**

By providing the details of your registered GP, you agreed to us contacting them to notify them of your interest in taking part in our study. If you report any adverse events to us during the study, we will report these back to your GP (in confidence). **If any significant HRFs are noted on your scans, these will be discussed with you and your GP. If recommended, your GP will arrange any further tests or specialist referrals for you.**

**Expenses and payments**

You will not be substantially out of pocket as a result of taking part in our study. We will reimburse travel expenses, and meal vouchers will be provided for Royal Infirmary of Edinburgh catering facilities on the day of scanning. Reimbursement for expenses incurred for childcare or loss of earnings will be considered on a case-by-case basis upon discussion with the Chief Investigator.

#### Is there anything I need to do or avoid?

Please wear the wristband provided until your scanning date and proceed with your normal daily routine. **The band is splash proof so it can be worn during hand washing, however please remove it when taking a bath or shower, or if swimming, and reapply it immediately afterwards. On the day of scanning, you must adhere to strict times for consumption of food** (6am-8am, 12 noon to 2pm and 6pm-8pm), **and caffeine** (6am-8am, and 12 noon to 2pm). You must **not consume alcohol or undergo any excessive physical activity on the day of scanning** (e.g. gym workout, running, cycling).

**Additional requirements for women with natural menstrual cycles:**

If you are a woman and not taking oral contraceptives, **your scans must be scheduled during the post-ovulation phase of your menstrual cycle**. We will confirm this with urine testing kits (ClearBlue). A kit with several test strips is provided today (on or just before your expected ovulation date). You can test your urine on up to three consecutive days (if needed) to confirm ovulation. **You must record the date and time of a positive result and notify the Chief Investigator.** Alternatively you can take a digital photograph of the result (i.e. on a mobile phone) and bring this with you on the morning of scanning (the photograph will be viewed by the Chief Investigator but not transferred or stored). If you are unable to confirm ovulation, further test strips will be provided, and you will be invited to reschedule your scanning day for the following month.

#### What are the possible benefits of taking part?

There are no direct financial or health benefits associated with taking part in this study. We do not know what the outcome will be, which is why we are conducting the research. Some volunteers may experience indirect benefits as follows:

- Better awareness of BMI and menstrual pattern (where applicable)
- Potential identification of an HRF that might be treated at an earlier stage **(please note that not all HRFs have health implications, and those that do, may not have treatment options)**
- Better engagement with research, researchers, and health professionals
- Contributing data from an under-researched group (women)
- Helping to address a fundamental knowledge gap that may help patients in the future

#### What are the possible disadvantages of taking part?

MRS scans are completely safe as long as routine MRI safety procedures are followed. MRI uses a magnet and radio waves to generate brain images. MRI and MRS scans do not involve any radiation, so there is no additional risk to having multiple scans. In the event of an unexpected medical emergency, the Imaging Facility has easy access to emergency medical care on-site. It is not possible to eliminate all risks of taking part in this study. You should be aware that stress or anxiety may result from the following:

- Reviewing the Participant Information Sheet or completing the Consent to Participate Form
- Attending in person for the consenting procedure and scanning day
- Wearing the wristband for 7-10 days
- BMI and oral temperature checks
- Urine testing (for some women)
- Attending 3 scans on time including a late evening scan
- Being positioned within the MRI scanner
- Restricted food and caffeine consumption on the day of scanning
- Prohibition of alcohol consumption on the day of scanning
- Restricted activity on the day of scanning

- Requirement to report any adverse events
- Identification of a HRF (especially for lesions that have no effective treatment options)
- Exclusion from the study after consenting to participate

**Please be aware that any HRFs identified in your brain scans might affect insurance policies you may have, or plan to take out.**

#### What if there are any problems?

If you have a concern about any aspect of this study, please contact Dr Nina Rzechorzek (Chief Investigator) who will do her best to answer your questions. You may also speak to a Consultant Radiologist (imaging specialist) who is not involved in the study. Contact details are provided at the end of this Information Sheet.

If you have a medical problem that occurs during the study and you cannot consent to take part any longer, or if you become unable to make decisions for yourself, we will exclude you from the study but we will use the information we have collected up to that point.

In the unlikely event that something goes wrong, and you are harmed during the study due to someone's negligence, then you may have grounds for legal action for compensation against the NHS or the University of Edinburgh. You may however have to pay your own legal costs. The normal NHS complaints mechanisms will still be available to you (if appropriate). The University of Edinburgh is liable for its employees' actions (undertaken as part of their job) and is insured against the risk of claims relating to research studies that their staff design and undertake. This insurance covers both negligence and no-fault compensation.

#### What will happen if I don't want to carry on with the study?

You can withdraw from our study at any stage, without giving a reason. Your decision to withdraw will be notified to your GP, but will not have any impact on your future care. There will be no penalty or loss of benefits (i.e. you will still be reimbursed for any expenses incurred as a result of being part of the study).

If you wear the wristband but then decide not to proceed with the scans, the data already collected from you will be safely destroyed. If you withdraw after your scans have been completed, but before analysis is complete, your data will no longer be included in the study. **However, any significant HRFs identified on your brain scans must still be reported to you and your GP (in confidence)**. You must also understand that once anonymised, it will not be possible to exclude your brain temperature data from the analysis and publication of the results, or from future research studies that use this anonymised data. The reason for this is that once the data is anonymised, we cannot identify which data belongs to you.

#### What happens when the study is finished?

##### The use of your data in further research

The data from your wristband and the brain temperature data generated from your scans will be anonymised prior to further analysis. Anonymised data will be shared between members of the Study Team (including Dr O'Neill, an expert in circadian biology at the MRC Laboratory of Molecular Biology Cambridge). Anonymised data will also be made available for use by other researchers internationally via deposit in a secure online database and will be retained indefinitely. The data collected during this study will be used as a standard against which to compare similar data from individuals of other age groups, and also patients with neurological disorders.

Some of the data collected for this study will be linked to you as an individual and will be retained securely for a minimum of 20 years. Such data will be used only for communicating with you and your GP as outlined above. With your consent, your email address will be used to notify you when the study results are published, and to inform you of future imaging studies that might interest you. **You can decline to be contacted for these purposes using the Consent to Participate Form.**

#### Will my taking part be kept confidential?

All the information we collect during the research will be kept confidential and there are strict laws that safeguard your privacy at every stage. Wristband data, brain images, BMI results, oral temperatures, and urine test results (where applicable) will only be identifiable by your participant number for the study and will not be labelled with any person-identifiable information.

With your consent, we have notified your GP that you are interested in taking part in this study. We will ask your permission to use your Community Health Index (CHI) number to communicate with your GP. The CHI is a population register, which is used in Scotland for healthcare purposes. The CHI number uniquely identifies a person on the index. Should the need arise, we will notify your GP of any significant HRFs. To ensure that the study is being run correctly, responsible representatives from the University of Edinburgh and/or NHS Lothian may need to access your medical records and data collected during the study, where it is relevant to you taking part. These institutions are Co-Sponsors for this study and are together responsible for its management and for providing insurance and indemnity.

#### What will happen to the results of the study?

You will not be provided with any specific rest/activity or brain temperature results relating to you as an individual. We will analyse the results from all participants as a group, and publish our findings in publically-accessible scientific journals and at scientific conferences. We will also make the results available on University of Edinburgh (Edinburgh Imaging) and MRC Laboratory of Molecular Biology websites. All data are anonymised so it will not be possible to identify you when we make our results public.

The results of this study will be used to design studies in the lab. These studies will use human stem cell-derived brain cells to understand how brain temperature interacts with the molecular clock inside brain cells. Your data may also be used to design future studies looking at how brain temperature changes with healthy ageing and shift work, and in neurological diseases such as dementia, brain cancer, mental illness, and sleep disorders.

#### Who is organising and funding the research?

This study has been organised by an Edinburgh Imaging Study Team, led by Dr Nina Rzechorzek (Chief Investigator) and supervised by Professor Ian Marshall at the University of Edinburgh. The study is being co-sponsored by the University of Edinburgh and NHS Lothian. The work is being funded by the Medical Research Council (MRC).

#### Who has reviewed the study?

The study proposal underwent independent external review as part of a MRC Clinician Scientist Fellowship application, led by Dr Rzechorzek. All research conducted using NHS facilities is reviewed by an independent group of people, called a Research Ethics Committee (REC), to protect your interests. This study has been reviewed and given a favourable opinion by the Academic and Clinical Central Office for Research & Development (ACCORD) medical research ethics committee (AMREC). NHS management approval has also been given.

#### Researcher Contact Details

If you would like further information about the study, or if you need to change your scheduled scan date, please contact the **Chief Investigator:** Dr Nina Rzechorzek, MRC Clinician Scientist Fellow,, or, telephone: 07900 815450 (mobile).

#### Independent Contact Details

Alongside the Edinburgh Imaging Radiologist team, Dr Dilip Patel or Dr Graham McKillop, Consultant Radiologists at the Royal Infirmary of Edinburgh (Tel 0131 536 1000) are happy to answer any further questions you may have. These persons are not directly involved in this study, and so will be able to give you independent advice.

#### Complaints

If you wish to make a complaint about the study please contact:

Patient Experience Team
2 – 4 Waterloo Place, Edinburgh, EH1 3EG

0131 536 3370

| Participant ID: | | Centre ID (if applicable) |
|---|---|---|

IRAS ID: 244533 Centre Number: Study Number: AC 18038

Participant Identification Number:

**CONSENT TO PARTICIPATE FORM**

**Study title: Can we measure a diurnal shift in brain temperature in healthy human volunteers using Magnetic Resonance Spectroscopy (MRS)?**

Chief Investigator: DR NINA RZECHORZEK

**Please initial each box**

1. I have read the Participant Information Sheet and Consent to Participate Form (version 1.9, 14th May 2019), and Data Protection Information Sheet (version 1.1, 6th April 2019) in relation to this study. I have been given ample opportunity to consider this information, ask questions about both documents, and have had these answered to my satisfaction. I have all the information I need to **provide informed consent to take part in this study.** ☐

2. I understand that my participation is voluntary; **I am free to withdraw at any time** without giving any reason, and without my medical care or legal rights being affected. As a volunteer, I am aware that I am not being scanned at the request of a doctor for any specific medical condition. ☐

3. I am aware that my scans will be viewed by a doctor qualified in medical imaging. I understand that images of my brain are being collected for research purposes only. These images are **not for diagnostic use** and would not be sufficient to rule out certain brain disorders. ☐

4. I understand that relevant sections of data collected during the study may be looked at by individuals from the Sponsors (NHS Lothian and the University of Edinburgh) where it is relevant to my taking part in this research. I give permission for those individuals to have access to my records and my CHI number. ☐

5. I understand that images obtained during my scan will be stored and processed using computers. After the study has ended, I am aware that these images may be copied onto a **permanent record** which might be studied again at a later date. ☐

6. I am aware that **incidental unexpected findings can occur when scanning healthy volunteers**. I understand that if any significant health-related findings are revealed through imaging my brain, these will be reported back to my GP who will then contact me to discuss their implications, and whether or not further tests are required. ☐

7. I am aware that unexpected findings **may affect insurance policies** ☐

| Participant ID: | | Centre ID (if applicable) |
|---|---|---|

8. I understand that the information collected about me might be used to support other research in future, and might be shared anonymously with other medical and scientific researchers. This will be subject to strict laws and University of Edinburgh policies intended to safeguard my privacy. ☐

9. **I know of no reason why I should not undergo Magnetic Resonance Imaging** or take part in this study. If I lose capacity during the study, I understand that the Study Team will **retain and use** the data collected from me prior to this. ☐

| | YES | NO |
|---|---|---|
| I give consent for my email address to be used to inform me of the study results, and details of new studies that I might be interested in taking part in. | ☐ | ☐ |

**10. By signing below, I give consent to take part in this study.**

Signature of volunteer

Name of volunteer (please print in block capitals)

Person taking consent (signature)

Name of person taking consent (please print in block capitals)

DR NINA RZECHORZEK

Date

CHI Number

Name and address of Volunteer's GP
