## Supplementary Appendix 2 for "Diurnal brain temperature rhythms and mortality after brain injury: a prospective and retrospective cohort study"

### Non-CTIMP Study Protocol

Can we measure a diurnal shift in brain temperature in healthy human volunteers using Magnetic Resonance Spectroscopy (MRS)?

| | |
|---|---|
| | The University of Edinburgh and Lothian Health Board<br>ACCORD<br>The Queen's Medical Research Institute<br>47 Little France Crescent<br>Edinburgh<br>EH16 4TJ |
| Protocol authors | Dr Nina Rzechorzek<br><br>Professor Ian Marshall<br><br>Dr Francesca Chappell<br><br>Dr Michael Thrippleton<br><br>Dr Duncan Martin<br><br>Dr John O'Neill |
| Funder | Medical Research Council |
| Funding Reference Number | MR/S022023/1 |
| Chief Investigator | Dr Nina Rzechorzek (MRC Clinician Scientist Fellow) |
| Sponsor number | AC 18038 |
| REC Number | 18-HV-045 |
| Project registration | UKCRN (NIHR CPMS) Portfolio (registration pending) |
| Version Number and Date | Version 1.7 May 14th 2019 |

| **Amendment classification and number:** | **Summary of change(s)** |
|---|---|
| Version 1.4 | Prospective Funder updated to MRC<br><br>Abbreviations list updated to include MRC, GDPR and UKRI<br><br>9.0 Addition of local SOP title and number<br><br>11.2.5 Data Protection section extended in line with MRC Data Management Plan<br><br>13.1 Authorship Policy extended in line with MRC Data Management Plan |
| Version 1.5 | Study logo and funder logo added<br><br>Title adjusted from 'MRSI' to 'MRS'<br><br>Funder confirmed as MRC<br><br>1.1 Actigraphy wristband added as potential burden and requirement of entry into the study to establish participant-specific chronotype. Pilot scan added to optimize MRS acquisition in deep brain region.<br><br>2. Secondary objective/endpoint relating to cooling effect of positioning/scan length removed<br><br>3.0 Study design adjusted to 'prospective, single-site, cohort', data collection period extended to 4 months (June to September 2019). $T_{Br}$ data to be normalized to individual chronotype. Study procedure adjusted to include actigraphy and scanning times/procedure simplified. BRIC SOP cited for HRF feedback pathway. CI employment details updated. Study schedule modified to remove public open seminar.<br><br>4.0 Inclusion criteria to include wearing of actigrpahy wristband<br><br>5.0 Identifying participants – actigraphy data to be destroyed if no consent is obtained to proceed with scans<br><br>7.0 Data collection to include actigraphy. Scanning protocol simplified/adjusted. |

| | |
|---|---|
| | 8.0 Data analysis to include chronotype<br><br>References – additional reference added relating to chronotype differences between sexes |
| Version 1.7 | Further clarification on collection, transfer and processing of actigraphy wristband data including potential risks.<br><br>Obtaining written informed consent to participate rescheduled (signatures will now be obtained prior to issue of actigraphy wristbands or urine test kits) |

Before the study can commence, all required approvals will be obtained and any conditions of approvals will be met. ... 25
11.2 INVESTIGATOR RESPONSIBILITIES ... 25
11.2.1 Informed Consent ... 25
11.2.2 Study Site Staff ... 26
11.2.3 GCP Training ... 26
11.2.4 Confidentiality ... 26
11.2.5 Data Protection ... 26
**12 STUDY CONDUCT RESPONSIBILITIES ... 27**
12.1 PROTOCOL AMENDMENTS ... 27
12.2 MANAGEMENT OF PROTOCOL NON COMPLIANCE ... 27
12.3 SERIOUS BREACH REQUIREMENTS ... 27
12.4 STUDY RECORD RETENTION ... 27
12.5 END OF STUDY ... 28
12.6 INSURANCE AND INDEMNITY ... 28
**13 REPORTING, PUBLICATIONS AND NOTIFICATION OF RESULTS ... 28**
13.1 AUTHORSHIP POLICY ... 28
**14 REFERENCES ... 29**

### LIST OF ABBREVIATIONS

| | |
|---|---|
| ACCORD | Academic and Clinical Central Office for Research & Development - Joint office for The University of Edinburgh and Lothian Health Board |
| AE | Adverse Event |
| AR | Adverse Reaction |
| BMI | Body mass index |
| CI | Chief Investigator |
| CiBraT | Circadian brain temperature |
| GCP | Good Clinical Practice |
| GDPR | General Data Protection Regulation |
| HRF | Health-related finding |
| ICH | International Conference on Harmonisation |
| MRC | Medical Research Council |
| MRS | Magnetic Resonance Spectroscopy |
| QA | Quality Assurance |
| REC | Research Ethics Committee |
| SAE | Serious Adverse Event |
| SAR | Serious Adverse Reaction |
| SOP | Standard Operating Procedure |
| SPDF | Study Participant Data Form |
| SUSAR | Suspected Unexpected Serious Adverse Reaction |
| $T_{Bo}$ | Body temperature |
| $T_{Br}$ | Brain temperature |
| UKRI | UK Research and Innovation |

Circadian Brain Temperature Study

MRC Medical Research Council

accord Academic and Clinical Central Office for Research and Development

### 1 INTRODUCTION

#### 1.1 BACKGROUND

*Circadian rhythms are fundamental to brain health*
Circadian clocks drive near-24-hour rhythmic changes in biological processes, adapting organisms to Earth's periodic rotation. These imprecise clocks are reset by environmental cues, which in mammals feed into a hierarchical multi-oscillator system; a 'master' clock deep in the brain synchronizes cell-autonomous clocks in the rest of the brain and periphery, coupling systemic adaptations to light-dark cycles. Age-related deterioration of the clock is compounded by modern living, which dissociates endogenous clocks from natural cues. Neurodegeneration — as occurs in dementia — is increasingly linked to clock disruption and presents a burgeoning socioeconomic threat to our healthcare system. To maintain brain health throughout life, we must understand how to maintain neural cellular timekeeping.

*Thermal resilience of the clock is mechanistically elusive*
The phase of mammalian circadian oscillations is highly sensitive to temperature shift, whilst circadian period (clock speed) remains constant across physiological temperatures. This 'temperature compensation' of periodicity comprises a hallmark feature of *bona fide* circadian rhythms, and is poorly understood at the molecular level. Failure of this mechanism may dictate the loss of neural cellular timekeeping in neurodegenerative disease. Thermoregulatory capacity declines with age; suboptimal thermal control could contribute to — and arise from — neural clock dysfunction. Brain temperature ($T_{Br}$) generally exceeds core body temperature ($T_{Bo}$), however, despite the importance of $T_{Br}$ to neural health, a basic understanding of human $T_{Br}$ fluctuation is missing. $T_{Br}$ is difficult to measure and varies by brain region, with neural activity, state of consciousness, and pathology. Although circadian $T_{Bo}$ cycles are well recognized in mammals, and have reduced amplitudes in aged humans, no studies have examined how human $T_{Br}$ varies across circadian time. To extract the relevance of thermal-clock interactions to brain health, the molecular clockwork must be studied at physiological human brain temperatures.

*CICs interact with the clock but causal molecular mechanisms are unknown*
CICs are 'accessory oscillators'; they are themselves circadian, serving both as inputs to, and outputs from, the core clock. Links between CICs and the circadian clock are expanding, but the influence of CICs on timekeeping in response to temperature change remains unexplored. If temperature compensation — and thus clock integrity — depends on CIC-clock interactions, these might be exploited to re-establish clock fidelity in vulnerable neural cells. In order to explore this possibility in the lab, we first need to know how temperature varies around the clock in the human brain. The proposed imaging study forms part of a Fellowship proposal to address the following question:

**How does $T_{Br}$ vary across the circadian cycle?**

Although we know that $T_{Bo}$ in healthy individuals varies in a circadian manner, it is currently not known whether $T_{Br}$ also varies in this way. In the clinic, human $T_{Br}$ can be measured directly using probes inserted into the surface of the brain. However, this invasive recording method can only be justified in critically ill patients with brain injury, and it only captures data from one focal brain region. MRS is currently the only method available to measure $T_{Br}$ in a non-invasive way. A previous study conducted by Edinburgh Imaging researchers demonstrated that this technique can generate $T_{Br}$ data from multiple brain regions in healthy human volunteers. The proposed study will extend our understanding of how human $T_{Br}$ varies around the clock by recording MRS temperature data in each volunteer at three different time points in a single 24-hour cycle. This study will therefore provide critical baseline information about diurnal variation in human $T_{Br}$ in healthy adults. The data produced will be used to inform mechanistic studies in the lab using human stem cell-derived brain cells to understand how $T_{Br}$ interacts with the molecular clockwork. The data will also be used to design future studies looking at how $T_{Br}$ changes in healthy elderly individuals as well as those with neurological diseases, such as Alzheimer's disease.

**Risk, burdens and benefits**

*Potential risks to participants*

- Inadequate informed consent - considered to cause possible harm to an individual, with potentially moderate long-term consequences of harm to that individual.
- Hazards of the interventions - the long-term effects of MRS scanning on the brain are considered negligible. The routine safety risks of MRI are considered negligible as long as the appropriate health and safety procedures are followed. Any participant could potentially experience a local allergic reaction to any new wearable device (e.g. actigraphy wristband), with possible minor harm. This hazard is considered extremely unlikely and would be easily managed without any long-term effects.
- Hazards of assessment methods – identification of a health-related finding (HRF) is considered to cause possible harm to an individual with potentially moderate consequences of harm to that individual.
- Failure to act appropriately upon health information discovered as a consequence of participating in the study – considered to cause possible harm to an individual, with potentially moderate consequences of harm to that individual (depending on the nature of the HRF).
- Failure to protect the privacy of participants – considered to cause possible harm to an individual, with potentially moderate consequences of harm to that individual.

*Potential burdens to participants*

- Requirement to attend in person for the consenting procedure and scanning day
- Requirement to wear a single wristband-based monitor (actigraphy wristband) that measures rest/activity patterns, skin temperature, and light exposure for one week prior to scanning
- Requirement to attend 3 MRI scans on time including a late evening scan
- Requirement to fix times of food and caffeine consumption, and avoid alcohol and excessive physical activity, on the day of scanning

*Potential benefits to participants*

- Better awareness of own health status including body mass index (BMI), sleep hygiene, and menstrual pattern (where applicable)
- Potential identification of an HRF that might be successfully treated at an earlier stage
- Engagement with research, researchers and health professionals

- Contributing data from an under-researched group (women)
- Learning about circadian biology, brain imaging, MRI safety, and non-invasive $T_{Br}$ measurement
- Helping to address a fundamental knowledge gap (contribution to knowledge and understanding of diurnal brain temperature variation in healthy individuals)
- Understanding that the results of the study will provide critical information to inform mechanistic studies *in vitro*
- Understanding that the results of this study may inform the development of novel treatment approaches for neurodegenerative diseases
- Understanding that the results of this study may inform new guidelines for $T_{Br}$ measurement and interpretation in humans

Overall the potential benefits of this study to the wider public outweigh the potential risks and burdens to individual participants.

#### 1.2 RATIONALE FOR STUDY

Circadian ('around 24 hour') rhythms orchestrate many biological functions and are driven by molecular clocks that exist in every cell - including cells of the brain. These biological clocks are essential for optimal brain health and the regulation of $T_{Bo}$. Emerging evidence suggests that brain cell clocks are disrupted in neurodegenerative diseases such as Alzheimer's disease. The risk of these diseases is highest in elderly individuals, who are notoriously poor at regulating their own temperature. In healthy brain cells, molecular clocks maintain circadian rhythms at constant speed, despite temperature fluctuation. Cold-inducible chaperone (CIC) proteins interact with cellular clocks and are highly sensitive to temperature change. We propose that CICs are critical to clock fidelity under $T_{Br}$ fluctuation, and that failure of this mechanism advances neurodegeneration. First, we must determine how human $T_{Br}$ varies with time; this represents a fundamental knowledge gap.

$T_{Br}$ fluctuation is reported in rodents, but circadian effects may be confounded by the sleep-wake cycle. $T_{Br}$ also depends on cerebral haemodynamics, and given species-specific variation in cerebral circulation, extrapolating animal data to humans is problematic. The current gold standard technique for human $T_{Br}$ measurement requires an intracranial probe, retrieving data from a focal point in the brain parenchyma. This is reserved for patients undergoing neurosurgery, or those with traumatic brain injury. With MRS, high-resolution $T_{Br}$ data can now be obtained non-invasively in healthy subjects. Pre-existing round-the-clock $T_{Br}$ data from brain-injured patients has informed the design of an MRS study in human volunteers, to measure healthy $T_{Br}$ at different times of the day. Derived maximum and minimum $T_{Br}$ will be used to explore how CICs are involved in the resilience of clocks to physiological temperature shifts using human stem cell-derived brain cells in the lab, potentially leading to novel therapeutic targets for chronic brain disorders.

*Hypotheses:*
(i) A diurnal shift in mean human $T_{Br}$ can be measured in healthy volunteers.
(ii) Mean human $T_{Br}$ exceeds oral temperature, irrespective of time.
(iii) Mean brain and oral temperatures are higher in luteal-phase women than in men.
*Method:* alongside brain injury effects, several confounders limit extrapolation of

patient data to the general population. Using MRS, we will map diurnal changes in $T_{Br}$ in healthy volunteers to determine normal human maximum and minimum $T_{Br}$ at unparalleled spatial resolution. Participants will be recruited locally and screened for suitability via electronic questionnaire. If eligible and willing to participate, they will be provided with the Participant Information Sheet and Consent to Participate Form. One visit 7 days prior to scanning will be used as the consenting procedure, and to issue actigraphy wristbands, and also urine test kits (where needed).. 40 subjects (20 men, 20 women) will each be scanned at 3 specific times (morning, afternoon, and late evening) within 24h, using a 3 Tesla MRI scanner. The scanning protocol will include whole brain, T1- and T2-weighted structural acquisition followed by MRS in a superficial and deeper brain location . All scans will be conducted within four months (minimizing seasonal variation) in a temperature-controlled room. Meal times and caffeine consumption will be fixed on the day of scanning and participants will be requested to avoid alcohol and excessive physical activity. Oral temperature will be measured prior to each scan, and hormonal influences controlled through urine testing (naturally cycling women only). Protocols for data extraction and temperature estimation by MRS are already validated and published. Anonymised MRS data will be processed blind (to time of day), and then analysed using a linear mixed model that accounts for time of day, subject-specific chronotype (based on activity data), and repeated measures for each participant.

### 2 STUDY OBJECTIVES

#### 2.1 OBJECTIVES

##### 2.1.1 Primary Objective

To determine whether human $T_{Br}$ varies according to time of day using a non-invasive brain imaging technique (MRS).

##### 2.1.2 Secondary Objectives

- Quantify the difference in variability of brain and oral temperatures across a 24-hour period in healthy human volunteers
- Quantify the difference between healthy male and luteal-phase female $T_{Br}$ at each time of day
- Quantify the difference between temperatures at the brain surface and the brain core at each time of day in healthy human volunteers

#### 2.2 ENDPOINTS

##### 2.2.1 Primary Endpoint

A statistically significant change in mean human $T_{Br}$ (across all measured voxels per subject) between time points.

##### 2.2.2 Secondary Endpoints

- The difference in variation of brain and oral temperatures across a 24-hour period

- The difference in brain and oral temperatures between men and luteal-phase women at each time point
- The difference between deep and superficial $T_{Br}$ at each time point

### 3 STUDY DESIGN

This is a prospective, single-site, cohort study in healthy human volunteers based at the Edinburgh Imaging (Royal Infirmary of Edinburgh) Facility. The study duration is 12 months from the start of recruitment to the completion of data analysis and submission for publication. Data collection will be scheduled over a 4-month period from June to September 2019. The anticipated duration of involvement for each participant is up to 17 days (from the date of obtaining informed consent to participate 7-10 days prior to scanning, to 7 days post scanning in case any adverse events need to be reported to the Study Team). The study length will be longer if scans need to be rescheduled for any participant. All participants will have the option to be notified by email when the results of the study are published (with Open Access).

The first key decision during the study design process was to determine the number of scans to be conducted for each participant. The logistical and financial burden of attending for scans was considered, and even though all participants will be local to the scanning facility, our approach is to minimize the impact of scanning on daily life, work, and family commitments. We decided that each participant should be requested to commit only to one scanning day, and that we would capture data at three time points during that single day. Although this removes the effects of day-today variation, it clearly cannot account for it, and this is an accepted risk of our method. The 3 time points were chosen based on available literature relating to $T_{Br}$ fluctuation in rodents, known $T_{Bo}$ cycles in healthy humans, and also pre-existing data collected from brain-injured patients as part of routine critical care. A 2-time point approach was considered but carried the risk of sampling twice at the midpoint between the temperature peak and trough. Overall our 3-time point approach, together with normalization of $T_{Br}$ data to individual participant chronotype, will maximize the opportunity to fulfill our recruitment target and meet primary outcome objectives.

The second key decision was for the inclusion of female participants. Women were excluded from previous studies using MRS to measure $T_{Br}$ in order to remove the potential confounding effect of the menstrual cycle on body (and possibly brain) temperature. We believe it is important, where possible, to include both sexes in studies of human brain physiology, such that the results of our study are as widely applicable as possible. Whilst it is well established that $T_{Bo}$ can fluctuate during different stages of the menstrual cycle, and that there is a clear difference in chronotype between sexes at certain life stages, the primary outcome for this study is whether there is an intra-individual shift in $T_{Br}$ between different times of the day. As such, each individual acts as their own control, and a linear mixed model approach can be used to address the effects of nesting at multiple levels when considering data within and between participants. It is also reasonably straightforward (and inexpensive) to control for chronotype differences (using activity monitors) and the effects of the menstrual cycle (by scanning all naturally cycling female participants at the same respective phase of their cycle i.e. post-ovulatory/luteal phase). Whilst this adds an additional non-invasive measurement (actigraphy) for all participants and an additional procedure (urine testing) for some female participants, we do not believe this would have more than a negligible impact on subject recruitment. Indeed, it means that unexpected chronotype extremes can be accommodated, and both naturally cycling women and women taking monophasic contraceptive medication can be included in the study. It will impact on logistics, reducing the number of dates on which some female participants are eligible for scanning, but this is a manageable risk within the 4-month duration of data collection. Commercially available ovulation urine testing kits were deemed to be a minimally invasive, inexpensive, and effective means of confirming the luteal phase of the cycle in female participants, in a way that could be readily confirmed and recorded by the CI, without compromising the privacy or health of the participant. Commercially-available actigraphy wristbands are widely validated and increasingly used by researchers as well as the health-conscious general public. The data collected from the wristband will be identifiable only by a unique participant identification number and will be downloaded via USB port to a secure University of Edinburgh computer prior to further processing. The data will be safely destroyed without use by the Study Team if a participant decides not to proceed with scanning.

The final key decision was the choice of method for $T_{Bo}$ measurement in order to exclude participants with $T_{Bo}$ beyond the normal range. After discussion of the relative risks and merits of more invasive methods (e.g. rectal temperature) we decided that oral temperature measurement would be widely acceptable to potential participants, would be more reliable than aural temperature measurement, and would be least likely to impact negatively on recruitment.

*Null hypothesis (1):* There is no statistically significant change in human $T_{Br}$ measured at different times of the day within a single 24-hour period
*Alternative hypothesis (1):* There is a statistically significant diurnal shift in human $T_{Br}$ within a single 24-hour period

This alternative hypothesis was chosen on the basis of known circadian fluctuation in human $T_{Bo}$ in healthy humans, recently observed circadian fluctuation in $T_{Br}$ in brain-injured patients that mimics $T_{Bo}$ cycling (unpublished), diurnal $T_{Br}$ fluctuation in rodents, and the ability of 3T MRS to detect small changes in human $T_{Br}$.

*Null hypothesis (2):* The variability in temperature within a single 24-jour period will not differ significantly between brain and oral regions of healthy human volunteers.

*Alternative hypothesis (2):* Human $T_{Br}$ variation will exceed oral temperature variation within a single 24-hour period in healthy human volunteers.

There is conflicting data comparing brain and core body temperatures in humans and other animals, but generally, $T_{Br}$ is considered to exceed core $T_{Bo}$, and we have recently observed a greater daily variability in $T_{Br}$ relative to $T_{Bo}$ in patients with traumatic brain injury (see References).

*Null hypothesis (3):* There is no statistically significant difference between brain temperatures measured in healthy men and women
*Alternative hypothesis (3):* $T_{Br}$ will be higher in luteal-phase women and in women taking monophasic contraceptive medication compared to men, after accounting for chronotype differences.

It is established that an increase in progesterone during the post-ovulatory (luteal) phase of the menstrual cycle increases $T_{Bo}$, and there is published evidence that women in the luteal phase of their cycle have higher body temperatures throughout the circadian cycle relative to men. There is also evidence to suggest that the synthetic steroids (progesterone and oestrogen) provided in the monophasic contraceptive pill are associated with a persistently increased $T_{Bo}$ throughout the cycle compared to the $T_{Bo}$ observed during the natural follicular phase in women not taking oral contraceptives. We predict that this may also be reflected in $T_{Br}$, and therefore we would expect luteal-phase women and women taking monophasic contraception to have higher brain temperatures that men, irrespective of the time of the day.

*Null hypothesis (4):* There is no statistically significant difference between mean temperatures at the brain core and the brain surface in healthy human volunteers
*Alternative hypothesis (4):* Mean temperature at the brain core will be consistently higher than that at the brain surface, irrespective of time.

Intuitively, we would expect mean temperature deeper within the brain to exceed that at the brain surface, based on the laws of physics.

**Study procedure**

Each participant will be scheduled to attend for three brain scans (approximate duration 45 minutes) at specific times on a single day (morning, afternoon, and late evening) using a 3 Tesla MRI scanner located in a temperature-controlled room at the Edinburgh Imaging (Royal Infirmary of Edinburgh) Facility. After consent is obtained, all participants will wear an actigraphy wristband for one week prior to scanning to establish their normal rest/activity, skin temperature, and light exposure patterns (and therefore their individual chronotype). On arrival at the imaging facility on their scanning day, each participant will have their height and weight recorded, and their actigraphy wristband will be removed and the data downloaded. Prior to each scan, the participant will be asked to change into hospital clothing and to complete an MRI safety and quality checklist (i.e. no metallic/cochlear implants, no metallic jewellery, no makeup that might create imaging artefacts). The participant's temperature will then be measured with an oral thermometer and if the temperature lies above or below the normal oral temperature range (33.2–38.1 °C for women; 35.7–37.7 °C for men), the participant will be rescheduled for an alternative day and any scans already performed on that participant will be excluded from the analysis. For female participants with natural menstrual cycles, a commercially-available urine testing kit (Clearblue) will be provided to confirm ovulation 7-10 days prior to their scheduled scanning date (several test strips supplied during the consenting interview so that urine can be tested for a luteinizing hormone surge on 3 consecutive days if necessary). Should ovulation not be confirmed, further test strips will be provided and the scans rescheduled for the following month. Kits will not be required for women taking monophasic contraception. No urine samples will be collected or stored from any participant; women using the kits must confirm ovulation by notification to the CI ahead of the planned scanning date, and will have the option of bringing a digital photo of the test result with them on the morning of scanning. The result will be viewed by the CI but no photos will be transferred or stored.

For each scan, the protocol will start with structural imaging of the whole brain using standard structural MRI sequences. The MRS protocol will immediately follow the structural scan and will consist of acquiring a 1cm thick section of MRS data near the brain surface, and MRS data from another region situated more deeply (including the hypothalamus). The participant will be given written guidance on which activities and consumable products are permissible in between scans and at what time. Meal times will be fixed on the day of scanning to 6am to 8am, 12noon to 2pm, and 6pm to 8pm. Caffeine consumption will be limited to 6am to 8am and 12 noon to 2pm. Participants will be permitted to leave the site between scans but must return 15 minutes prior to their next scheduled scan. If a participant fails to attend their first morning scan (or attends late), they will be invited to reschedule their scanning day once. Participants will be advised that if they attend late to their afternoon or evening scans, these cannot proceed, since the data would be invalid. Participants will be asked to inform the CI via email of any adverse events in the 7 days post-scanning.

All structural scans will be reviewed by a neuroradiologist to check for any abnormalities. All relevant HRFs will be confidentially reported to the participant and their respective GP, adhering to Edinburgh Imaging CRFSOP 19.02 BRIC v03 Radiological Reporting of Research Scans. The neuroradiologist will inform the CI whether or not a participant's data is suitable for inclusion in the analysis (but the CI will not be given any specific information about reasons for exclusion). Only MRS data from structurally-normal brains will be included for analysis.

The study will be led by Dr Nina Rzechorzek (the CI; MRC Clinician Scientist Fellow at the MRC Laboratory of Molecular Biology, Cambridge, and Honorary Staff Member at the Centre for Clinical Brain Sciences, University of Edinburgh), supervised by Prof Ian Marshall (Centre for Clinical Brain Sciences and Edinburgh Imaging). Dr Rzechorzek is a European Board-eligible Specialist in Veterinary Neurology and Neurosurgery and has previous research experience in both advanced neuroimaging (using MRI in canine patients), and also in conducting human clinical trials. Dr Rzechorzek has also completed GCP training. Prof Marshall holds a Personal Chair in Magnetic Resonance Physics and supervised a previous study testing MRS to measure brain temperature in healthy human volunteers. The Edinburgh Imaging Study Manager (Dr Duncan Martin) will oversee logistics and has extensive experience in managing MRI-based research studies with human volunteers. Dr Francesca Chappell (a Medical Statistician based at the Centre for Clinical Brain Sciences) has performed calculations to estimate the number of volunteers required to answer our research question, and she will oversee the statistical analysis of the data generated. Dr Michael Thrippleton (a Medical Physicist in MRI based at the Centre for Clinical Brain Sciences) led a previous study on brain temperature mapping using MRS and will provide intellectual support for this work. This study is part of a wider Fellowship programme designed by Dr Rzechorzek to understand the interaction between $T_{Br}$ fluctuation and the molecular clock in human brain cells. The Core Sponsor for this programme is Dr John O'Neill (MRC Laboratory of Molecular Biology, Cambridge), who is an internationally-recognised expert in circadian biology and has assisted with the design of the imaging arm of the proposal and the selected time points for imaging.

*Reasoning for study design and methodology:*
MRS is currently the only method available to accurately measure human $T_{Br}$ non-invasively. MRI-based brain imaging provides exceptional resolution of brain structure and is the gold-standard means by which brain lesions are diagnosed routinely in the clinic. MRS is a special form of MRI that can be used to estimate $T_{Br}$ very precisely and carries no additional risk to participants compared to MRI alone. MRI is a very

safe method of brain imaging with no known lasting effects on the brain after repeated imaging. Although intravenous contrast methods are sometimes used for MRI diagnostics, contrast is not required for MRS and will not be used in this study. Therefore no invasive procedures will be performed, but the lack of post-contrast imaging means that some HRFs may require follow-up imaging to verify them. The feedback and follow-up pathway is explained in the Participant Information Sheet.

A 3 Tesla (3T) MRI scanner is proposed for this work, because a recent study published by members of the Study Team showed that 3T MRS was more reliable than 1.5T MRS in estimating $T_{Br}$ in healthy men. In that study, all scans were carried out at the same time of day to control for any effects of diurnal temperature fluctuation. In this study, the diurnal variation in human $T_{Br}$ is the key parameter of interest and so each volunteer will be scanned at specific times in the morning, afternoon, and late evening on their scheduled scanning date. MRI-based brain imaging remains a fairly time-consuming process requiring high technical skill for acquisition and processing. Repeated brain imaging of each volunteer on a single day places a level of burden on that individual and on staff running the scanner. A three time point design was chosen to maximize the opportunity to meet our primary research objective whilst minimizing the burden on individual volunteers and radiographers. The selected time points are based on preliminary analysis of pre-existing data from human brain injury patients showing when $T_{Br}$ was recorded at its maximum and minimum level via intracranial probe. Whilst we cannot conclude that $T_{Br}$ peaks and troughs will occur at these same times in healthy human volunteers, the spacing between the time points should ensure that we do not inadvertently capture two times at which $T_{Br}$ will be the same within a 24 hour cycle. The selected time points are also sympathetic to volunteer and staff needs, and aim to prevent any disruption to sleep patterns that may affect the results. More frequent sampling would be neither practical, nor cost-effective and would likely present a major barrier to recruitment.

The inclusion and exclusion criteria have been specifically developed to maximize participant safety, compliance, and wellbeing, whilst controlling for confounding effects that might impact upon $T_{Br}$ and $T_{Bo}$, and/or thermoregulation generally. In particular, $T_{Br}$ is highly dependent on blood flow to the brain, and on the sleep/wake cycle. We have therefore excluded people with known sleep disorders or with medical conditions that would be expected to affect general circulation, cerebral blood flow, or thermoregulation. Thermoregulation is also known to decline with age and is subject to variation in body mass, and so the age range has been limited to 20-40 years, and the BMI to 18.5-29.9. The proposed age range is consistent with the ages of healthy adult males recruited into the previously published study, and from which $T_{Br}$ data was successfully acquired through MRS. The inclusion of women is a step forward from the previous study, which excluded women to avoid potential confounding effects of the menstrual cycle on $T_{Br}$. For the results of the prospective study to be widely applicable, it is important to include women whilst controlling for these effects. We believe that our approach is scientifically justified, easy to achieve, inexpensive, and logistically feasible within the duration of the study. We accept that for naturally cycling women there will be additional burden (urine testing in a location of their choosing, and a reduced selection of scanning dates) that may impact on their recruitment. However, we believe that this small impact is outweighed by the negative impact on recruitment if these women were to be excluded. Finally, the actigraphy data will allow us to control for differences in chronotype which are known to vary between the sexes, as well as between individuals of the same sex.

**Proposed timetable**

- September 2018 IRAS submission for R&D and ethics approval
- March 2019 Funding outcome announced
- March 2019 Study Team meeting to review/update Study Protocol and finalise recruitment strategy
- April/May 2019 revised and finalised Study Documents submitted for updated approvals ahead of recruitment
- May 2019 Commence recruitment
- May/June 2019 GP letters sent for 50 prospective participants
- June/July 2019 Complete initial recruitment (with 40 subjects called for interview and 10 subjects on 'stand-by' list)
- June to September 2019 Scanning, data collection, preliminary MRS data processing
- October to November 2019 Data analysis, statistical analysis
- December 2019 Manuscript preparation (followed by submission when finalized)
- From December 2019 Presentation of results at conferences/symposia, aggregate feedback to participants
- Deposition of averaged anonymised MRS data in a secure electronic Open Access repository upon publication

*Procedures to detect and compensate for potential researcher bias*

- The first 20 men and 20 women that satisfy inclusion/exclusion criteria will be invited for a consenting interview; an additional 5 men and 5 women will be invited onto a 'stand-by' list and may be invited to attend for scanning if there are interim withdrawals or exclusions.
- Recruitment will close after 25 eligible men and 25 eligible women are identified, and will re-open only if necessary.
- Volunteers will be randomised to a single scanning weekday (according to their availability) within the 7-10 days following consent
- No subjects will be scanned on a weekend
- All MRS data processing will be performed blind to sex and time of day
- After processing is complete, the CI will be unblinded in order to perform data interpretation and statistical analysis
- Statistical analyses will be supervised by a medical statistician (Dr Chappell)

### 4 STUDY POPULATION

#### 4.1 NUMBER OF PARTICIPANTS

The required sample to achieve our primary endpoint is 36 healthy human volunteers, aged 20-40 years, and living within a 5 mile radius of the Imaging Facility. All scans will be conducted at one site. The study cohort will include equal numbers of men and women. 40 subjects will be recruited for scanning to account for participant withdrawal, exclusion due to HRFs, and scan failure (for technical reasons). Recruitment will take place over a 5-month period (May 2019 to September 2019).

#### 4.2 INCLUSION CRITERIA

- Healthy adult human volunteers (men or women)
- Age 20-40 years
- Body mass index 18.5-29.9
- Live within 5-mile radius of Edinburgh Imaging (Royal Infirmary of Edinburgh) Facility
- No MRI contraindications
- Women with regular natural menstrual cycles for a minimum of 6 months, or women that have taken monophasic hormonal contraception (fixed dose oestrodiol and synthetic progestin) for a minimum of 3 months prior to scanning and will continue to do so during the month of scanning.
- Capacity to understand written and verbal information provided in English, and able to provide valid written informed consent to participate
- Able to commit to 3 x 45-minute scanning protocol within selected 24-hour period (in the post-ovulation phase of their cycle for women with natural menstrual cycles), and within the specified study period.
- Able to wear an actigraphy wristband for at least 7 days prior to scanning
- Able to commit to fixed times of food and caffeine consumption on the day of scanning
- Able to avoid alcohol and excessive physical activity on the day of scanning

#### 4.3 EXCLUSION CRITERIA

- Pregnant women
- Early menopause, irregular menstrual cycles, premenstrual syndrome, lack of luteinizing hormone surge (according to urine test) prior to two scheduled scanning days (women with natural menstrual cycles only)
- Body mass index < 18.5 or > 29.9
- Age < 20 years or > 40 years
- MRI contraindications (e.g. cardiac pacemaker, cochlear implant, claustrophobia, any prior accident in which metal penetrated one or both eyes)
- Oral temperature outside of normal range (33.2–38.1°C for women; 35.7–37.7°C for men) prior to any scan
- Medical history that might limit activity or alter cerebral blood flow (hypothyroidism, stroke, severe arthritis, Parkinson's disease, dementia, history of brain trauma, brain tumour or epilepsy, significant mental illness besides clinical depression, spinal cord injuries, recent serious burn, diabetes, dehydration)
- Taking medications (except seasonal allergy medication, over-the-counter NSAIDs, or contraceptives), acetaminophen (paracetamol), drug abuse
- Known neurodevelopmental, neuropsychiatric or neurodegenerative disorder
- Known sleep or chronotype disorder (delayed sleep phase syndrome (CRY1), familial advanced sleep phase syndrome (PER2))
- Known family history of cardiovascular disease at < 40 years of age
- Consumption of food or caffeine out with the specified time periods on the day of scanning
- Consumption of alcohol or participation in excessive physical activity on the day of scanning
- Failure to attend, or late attendance at, two independently scheduled morning scans

- Participant-reported ill-health on the day before or the morning of scanning

### 5 PARTICIPANT SELECTION AND ENROLMENT

#### 5.1 IDENTIFYING PARTICIPANTS

Recruitment will be conducted locally by the CI using standard approaches employed by Edinburgh Imaging. This will include a recruitment drive using mailshots to University of Edinburgh and NHS staff (including Edinburgh Imaging and Edinburgh Neuroscience subscribers), social media posts, and posters displayed at University of Edinburgh campuses and NHS Lothian hospitals. Only names and email addresses will be collected at the initial recruitment stage. Prospective participants will then be invited by email to complete an electronic questionnaire during which they can view the full list of inclusion and exclusion criteria for the study, the Participant Information Sheet, and the Consent to Participate Form. They will then be asked whether they meet the study criteria and whether or not they wish to participate. Non-willing participants will be invited to comment on why they have declined to participate. Willing participants will be asked to indicate their availability for consenting interview and scanning, and to provide written consent for the CI to contact their GP, by providing the name and address of their GP, and their own post code.

Review of medical records may only be conducted by the direct care team (the participant's GP) and/or regulatory authorities of the co-sponsors (where it is relevant to an individual's participation in the study). Letters will be sent by the CI to each prospective participant's GP, to notify them of the participant's willingness to take part in the study and listing the inclusion/exclusion criteria. GPs will have the opportunity to contact the CI with any concerns, but will not be obliged to review medical records or confirm eligibility to participate. GPs will be advised that they will be contacted by the Study Team if HRFs need to be discussed or acted upon.

The first 20 eligible men and 20 eligible women will be invited by the CI via email to attend a consenting interview at the Imaging Facility. During this interview, each participant will be provided with hard copies of the Participant Information Sheet and Consent to Participate Form to review. The participant will be invited to ask any questions and then initial and sign the Form (witnessed and co-signed by the CI). They will then be issued with an actigraphy wristband (and urine test kit if needed). Recruitment will be finalised by the CI on the morning of scanning, during which willingness to participate will be rechecked and documented. Only the minimum required person-identifiable data (name, email address, postcode, and GP details) will be collected during recruitment. Actigraphy data will be downloaded via USB port to a secure University of Edinburgh computer and will be used only if volunteers choose to proceed with scanning, otherwise it will be safely destroyed. For collection and transfer, the actigraphy data will be identifiable only by a unique participant identification number.

Participants will not be financially advantaged or disadvantaged as a result of participating in the study, nor will any forms of coercion be used to encourage recruitment. No incentives (promises of therapeutic benefit, diagnostic information, or access to personal imaging data) will be offered during recruitment. There will be no financial incentives for participation, but travel and meal expenses to take part in the study will be provided. Participation in the study will be entirely voluntary. No undue influence will be exerted when approaching potential participants and no sanctions will follow if the participant decides to leave the research at any time. Vulnerable persons and adults lacking capacity will not be knowingly approached for recruitment.

#### 5.2 CONSENTING PARTICIPANTS

The CI is a practising referral-level veterinary clinician who has completed specialist training in Neurology and Neurosurgery. As such, the CI has extensive experience of neuroimaging (particularly MRI techniques) and has undergone both GCP and Consent and Transparency training. The CI was also involved with the recruitment of healthy human participants for two successful double-blinded randomized controlled trials. The CI therefore understands the ethical principles underpinning informed consent and is familiar with the legal frameworks governing the consenting procedure. This procedure will ensure that each prospective participant has been provided with sufficient information and has the capacity to:

- Understand the purpose and nature of the research
- Understand what the research involves, its benefits (or lack of benefits), risks and burdens
- Understand the alternatives to taking part
- Retain the information long enough to make an effective decision
- Make a free choice
- Make this particular decision at the time it needs to be made

Consent will be obtained electronically to notify prospective participants' GPs of the participant's willingness to take part. The first 40 eligible participants (20 men and 20 women) will be invited (by emailed letter) for interview 7-10 days in advance of their selected scanning date to conduct the consenting procedure. Remaining eligible participants will be informed (by emailed letter) that they are on a 'stand-by' list and may be invited to participate at a later date in the event of subject withdrawal/exclusion.

All prospective participants will be given a Participant Information Sheet to explain the purpose and procedures of the study and valid, written informed consent to participate will be obtained once remotely (to notify respective GPs), and again during face-to-face interview with the CI at the Royal Infirmary of Edinburgh, 7-10 days prior to scanning. During this interview all participants will be provided with an actigraphy wristband to wear until their scan date and female participants with natural menstrual cycles, will be provided with a commercially-available urine testing kit (Clearblue) to confirm ovulation prior to their scheduled scanning date.

Following local guidelines, all participants must agree during the consenting procedure to be informed of any relevant HRF information arising from their brain scan. They must also understand that this information will be reported confidentially to their GP. Written consent to participate will be obtained once the participant has had time to consider all of the information provided, and after willingness to participate has been rechecked.

In addition, the consenting procedure will cover the following:

- Current and potential future risks of data use
- The risks of uncovering an HRF and how this will be handled
- The right and ability to withdraw; the circumstances under which it will not be possible to withdraw pre-existing unlinked anonymised participant data will be made clear

It is a condition of this study that all participants have the capacity to understand written and verbal information provided in advance of obtaining written informed consent to participate. Whilst the risks of MRI are very few and can be effectively controlled through appropriate safety screening, the harm to an individual if they do not understand these risks is potentially very serious. The CI will be responsible for ensuring that each participant is provided with sufficient information to understand the nature of the research and the risks involved. A number of techniques (written, visual, and interactive verbal) will be used to repeat the most salient points and confirm understanding of each participant.

The consent material and the consent process is appropriate for the study population, and meets ethical requirements. Potential participants will have sufficient time to discuss the study or consider whether to take part. The proposed study and consenting procedure will be subject to appropriate ethics review. A record of consent and willingness to participate will be kept for each interaction of the participant with Study Team members. The CI fulfills the inclusion criteria for participation and is well placed to understand the needs and perspectives of the study population.

##### 5.2.1 Withdrawal of Study Participants

Participants are free to withdraw from the study at any point or a participant can be withdrawn by the Investigator. If withdrawal occurs, the primary reason for withdrawal will be documented in the Study Participant Data Form (SPDF), if possible. The participant will have the option of withdrawal from all aspects of the study but it is made clear in the Participant Information Sheet that anonymised data collected from that individual up to the point of withdrawal may still be used by the Study Team. If a participant loses capacity to consent during the study, they would be withdrawn from the study. However, identifiable and anonymised data already collected with consent would be retained and used in the study. No further data would be collected or any other research procedures carried out on or in relation to the participant.

The right and ability to withdraw is clearly explained in the Participant Information Sheet and Consent to Participate Form; the circumstances in which withdrawal of pre-existing unlinked anonymised participant data will not be possible is also articulated in these documents. Notification of withdrawal can be indicated verbally but should also be confirmed in writing (email or letter) to the Study Manager who will also inform the CI using linked anonymisation. The decision to withdraw will be documented by the Study Manager in the source data on the date of receipt of notification. The respective participant's GP will also be notified in writing of a withdrawal, which they must then record in the participant's clinical notes. There are no anticipated safety issues post-withdrawal. All members of the Study Team know the process to follow in the event of a withdrawal (i.e. who to inform and how it should be flagged that a participant has withdrawn).

The CI reserves the right to withdraw any participant at any time for one or more of the following reasons:

- Identification of an HRF
- A participant becomes unable to meet the inclusion criteria during the course of the study
- A participant satisfies a criterion for exclusion during the conduct of the study
- The participant loses capacity to give valid consent to participate during the study

### 6 STUDY ASSESSMENTS

#### 6.1 STUDY ASSESSMENTS

Each participant will undergo the same assessments as part of the research protocol. Women with natural menstrual cycles will undergo one additional assessment; this will comprise a urine test to confirm post-ovulation status during the scanning sessions. A urine testing kit will be provided during the consenting procedure (7-10 days prior to scanning) and urine testing will be performed by the participant at home; optionally they may record the result by digital photograph (e.g. on a personal mobile phone). The participant will be asked to confirm ovulation by notification to the CI prior to attending the scanning day. On the morning of scanning, the participant will have the option of showing the CI photographic evidence of the urine test result. The test result will be viewed and recorded by the CI but no photographic images will be transferred or stored.

Each participant will undergo three (approximately 45-minute) brain scanning sessions in a 3T MRI scanner at the Edinburgh Imaging (Royal Infirmary of Edinburgh) Facility on their scheduled scan date (in the morning, afternoon, and late evening). Just prior to their first scheduled scan, each participant will have their actigraphy wristband removed, and their height and weight measured to calculate their body mass index (BMI). They will then be asked to complete an MRI safety checklist before their oral temperature is measured by digital thermometer. The MRI safety checklist and oral temperature measurement will be repeated prior to the second and third scans of the day. During each scan, the whole brain will be imaged to check for any structural abnormalities and to provide structural information to aide localisation and interpretation of $T_{Br}$ data. MRS scanning will then be performed in superficial and deep regions of the brain in order to obtain data that can be used to estimate brain temperature.

### 7 DATA COLLECTION

*Details of observational components of the research methodology and how these will be carried out:*

- Time points for data collection will be specified for each participant in the morning, afternoon, and late evening on their designated day of scanning. BMI will only be measured once immediately prior to the first scan.
- Body weight, height, and oral temperature measurements will be collected by the CI; structural and MRS brain scanning data will be collected by the attending radiographer

- Standardised tools will include wristband-based actigraphy in the 7 days preceding scanning, BMI calculation from body weight and height measurements, oral temperature measurements via digital thermometer, urine testing for ovulation checks (where applicable), structural brain analysis via MRI, $T_{Br}$ estimation via MRS.
- Methods to maximise completeness of data collection will involve the CI sending an email reminder to each participant 24 hours before scanning. Participants who fail to attend for their first morning scan will be emailed to reschedule their scanning day (there will be one opportunity to reschedule for failed attendance)

Participant rest/activity, skin temperature, and light exposure patterns will be monitored non-invasively over a minimum of 5 days prior to scanning using an actigraphy wristband.

Oral temperature will be measured by the CI immediately prior to subject positioning on the scanner using a precalibrated digital thermometer covered in a disposable one-use thermometer sleeve. The thermometer will be positioned under the tongue with the mouth closed and held in place for 40 seconds. Date, time and temperature will be recorded. Scanning will proceed only if the temperature is within the normal range.

Urine testing will only be necessary for naturally cycling women recruited into the study. The CI will have prior knowledge of which women require urine testing and a urine testing kit will be provided during the consenting procedure. Participants that require urine testing will be asked to perform the test as directed in the manufacturer's instructions and then take a note of the result together with the date and time it was obtained (optionally they can take a digital photograph of the result using a mobile phone or alternative). They must notify the CI of the result prior to the scanning day so that confirmation of a luteal surge can be recorded by the CI in the Source Data. Participants have the option of bringing a photographic image of the result on the morning of scanning but no images will be transferred or stored.

At each scanning time point the scanning protocol will consist of whole-brain T1- and T2-weighted structural acquisition followed by MRS acquisition in a superficial brain region then a deep brain region. Protocols for data extraction and temperature estimation are established (see Thrippleton *et al. NMR Biomed* (2014)). Spectroscopic acquisition from the deep brain region will include the hypothalamus (which contains the suprachiasmatic nucleus where the ‘master clock’ is situated), and the procedure will be optimized through a pilot scan at Edinburgh Imaging. Scanning will be performed using a PRISMA 3-T clinical MRI scanner (Siemens AG, Healthcare Sector, Erlangen, Germany). MRS $T_{Br}$ data will be obtained using sequences optimized for each brain region. All images and MRS data will be analysed offline using validated methods (see References).

#### 7.1 Source Data Documentation

Source documents will include the following:

*Electronic questionnaire* — emailed to prospective participants who have given consent to be contacted in this way by providing their email address. The questionnaire will document whether or not a participants believes they meet the criteria for inclusion; whether or not they are willing to participate; and their availability for the consenting interview and scanning. It will also provide an option to provide reasons for non-willingness to participate. By providing the name and address of their GP as well as their own postcode, prospective participants will be giving consent for their GP to be contacted.

*Consent to Participate Form* — to be completed by the participant with CI immediately prior to issue of the actigraphy wristband (7-10 days prior to scanning). Three copies must be signed by the participant and co-signed by the CI; one copy for the participant to keep, one copy to be added to the medical notes (sent to GP), and one copy for the Source Data File.

*Source Data notes in standardized hard copy File Note and Appointment Checklist formats* — on the day of scanning the CI will record participant ID, document willingness to participate, document any adverse events occurring since the consenting interview (and during the 7 days post-scanning), record confirmation of actigraphy wristband use (minimum 5 days) and ovulation (where applicable), record date, time, body weight, height, oral temperature, any deviations from or non-compliance with the Study Protocol or GCP, and any Serious Breaches.

*Image files* — MRI and MRS data will be stored on secure servers at Edinburgh Imaging prior to further processing.

*Actigraphy data files* – data will be identifiable only by unique participant number during collection and transfer via USB port to a secure University of Edinburgh computer. Data will be processed and used only if the participant proceeds with scanning, otherwise it will be safely destroyed.

##### 7.2 Study Participant Data Forms

Source Data (participant ID, date, time, sex, DOB, height, weight, BMI, oral temperature, confirmed actigraphy recording and ovulation where applicable) will be carefully copied to a standardized paper-format SPDF template by the CI on the day of scanning. Completed SPDFs will be added to the Site File.

#### 8 STATISTICS AND DATA ANALYSIS

##### 8.1 SAMPLE SIZE CALCULATION

The sample size was estimated by a medical statistician (Dr Francesca Chappell) based on achieving the primary outcome using a linear mixed model for the analysis, and using previously published data exploring the reliability of MRS to measure $T_{Br}$ in healthy human subjects. With a sample size of 36 subjects, and conservative true mean $T_{Br}$ difference of 0.5°C for the primary outcome measure, there is 80% power to detect a statistically significant difference between time points at the 5% significance level. In total we aim to fully consent and scan 40 eligible participants to account for losses due to withdrawal, pertinent incidental HRFs that may require subject exclusion, and failed scans (e.g. technical failure of the scanner). A completion of a feedback pathway for 2 volunteers is expected (based on 5% prevalence of incidental HRFs using high-resolution MRI (see References). A 5-month recruitment period is considered entirely feasible to meet the required sample size.

#### 8.2 PROPOSED ANALYSES

*Dependent variables* = $T_{Br}$; oral temperature. Means and standard deviations will be reported
*Independent variables* = time of day; sex; BMI; chronotype; phase of menstrual cycle (for naturally cycling women); brain region (deep/superficial); duration of scan

The data collected during the course of this study is nested at several levels (participant, sex, chronotype, time of day, duration of scan etc), and as such, complex statistical modelling is required. The expertise of a medical statistician are critical to the analysis of the data; Dr Chappell will oversee and supervise all statistical analyses required for this study. A linear mixed model approach will be used to accommodate data incorporating repeated measurements per participant. A linear mixed model will account for the fact that $T_{Br}$ measurements will be correlated within individuals in the analysis; it will also allow us to use data from all participants provided they undergo at least one, but not necessarily all 3 scans. This will ensure best use of participant and staff time.

HRF-based exclusion, and non-compliance with the Study Protocol will render the entire data set retrieved from a given individual unusable. The consequences of non-compliance are clearly explained in the Participant Information Sheet. Participants who are late for, or fail to attend, their first scan will be invited to reschedule their entire scanning day. Naturally cycling female participants who cannot demonstrate ovulation via urine test prior to their scheduled scanning day will be invited to reschedule urine testing and scanning for the following month.

The core analysis focuses on comparing mean $T_{Br}$ between different time points across the cohort. Pre-defined subgroup analyses will include mean $T_{Br}$ comparisons between men and women at each time point, and mean $T_{Br}$ between deep and superficial brain regions at each time point.

Given the small scale of this study, interim analyses and reports are not appropriate. Additional volunteers will be recruited if sufficient eligible participants are not identified from the initial approach

### 9 ADVERSE EVENTS

Adverse events will be recorded in the Source Data; for HRFs reporting will adhere to Edinburgh Imaging CRFSOP 19.02 BRIC v03 Radiological Reporting of Research Scans.

### 10 OVERSIGHT ARRANGEMENTS

#### 10.1 INSPECTION OF RECORDS

Investigators and institutions involved in the study will permit the review of all study documentation by the sponsors and the REC. In the event of audit or monitoring, the Investigator agrees to allow the representatives of the sponsor direct access to all study records and source documentation.

### 11 GOOD CLINICAL PRACTICE

#### 11.1 ETHICAL CONDUCT

The study will be conducted in accordance with the principles of the International Conference on Harmonisation Tripartite Guideline for Good Clinical Practice (ICH GCP).

Before the study can commence, all required approvals will be obtained and any conditions of approvals will be met.

#### 11.2 INVESTIGATOR RESPONSIBILITIES

The Chief Investigator is responsible for the overall conduct of the study at the site and compliance with the protocol and any protocol amendments. In accordance with the principles of ICH GCP, the following areas listed in this section are also the responsibility of the Investigator. Responsibilities may be delegated to an appropriate member of study site staff.

##### 11.2.1 Informed Consent

The Chief Investigator is responsible for ensuring informed consent is obtained before any protocol specific procedures are carried out. The decision of a participant to participate in clinical research is voluntary and should be based on a clear understanding of what is involved.

Participants will receive adequate oral and written information – appropriate Participant Information Sheets and informed Consent to Participate Forms will be provided. The oral explanation to the participant will be performed by the Chief Investigator, and must cover all the elements specified in the Participant Information Sheet and Consent to Participate Form.

The participant will be given every opportunity to clarify any points they do not understand and, if necessary, ask for more information. The participant will be given sufficient time to consider the information provided. It will be emphasised that the participant may withdraw their consent to participate at any time without loss of benefits to which they otherwise would be entitled.

The participant will be informed and agree to their medical records being inspected by regulatory authorities and representatives of the sponsors.

The Chief Investigator and the participant will sign and date the informed Consent to Participate Form to confirm that consent has been obtained. The participant will receive a copy of this document and a copy will be filed in the Investigator Site File (ISF) and participant's medical notes (complete form sent to GP).

##### 11.2.2 Study Site Staff

The Chief Investigator must be familiar with the protocol and the study requirements. It is the Investigator's responsibility to ensure that all staff assisting with the study are adequately informed about the protocol and their study-related duties.

##### 11.2.3 GCP Training

For non-CTIMP studies all researchers are encouraged to undertake GCP training in order to understand the principles of GCP. However, this is not a mandatory requirement unless deemed so by the Sponsor. GCP training status for the Chief Investigator is indicated in their respective CV.

##### 11.2.4 Confidentiality

All evaluation forms, reports, and other records must be identified in a manner designed to maintain participant confidentiality. All records must be kept in a secure storage area with limited access. Clinical information will not be released without the written permission of the participant. The Investigator and study site staff involved with this study may not disclose or use for any purpose other than performance of the study, any data, record, or other unpublished, confidential information disclosed to those individuals for the purpose of the study. Prior written agreement from the sponsor or its designee must be obtained for the disclosure of any said confidential information to other parties.

##### 11.2.5 Data Protection

All Investigators and study site staff involved with this study must comply with the requirements of relevant UK Data Protection legislation with regard to the collection, storage, processing and disclosure of personal information and will uphold the Act's core principles. Access to collated participant data will be restricted to individuals from the research team interacting with the participants and representatives of the sponsors.

Computers used to collate the data will have limited access measures via user names and passwords. Published results will not contain any personal data that could allow identification of individual participants.

All data collected will be compliant with the UK Data Service Data Security standards (https://www.ukdataservice.ac.uk/manage-data/store/security) and the revised GDPR (https://www.eugdpr.org). MRC guidance on confidentiality and data security (https://mrc.ukri.org/publications/browse/mrc-policy-and-guidance-on-sharing-of-research-datafrom-population-and-patient-studies/) and UK Data Service guidance on anonymisation and controlling access to shared research data (http://www.ukdataservice.ac.uk/manage-data/legalethical/ access-control) will be followed.

### 12 STUDY CONDUCT RESPONSIBILITIES

#### 12.1 PROTOCOL AMENDMENTS

Any changes in research activity, except those necessary to remove an apparent, immediate hazard to the participant in the case of an urgent safety measure, must be reviewed and approved by the Chief Investigator.

Amendments will be submitted to a sponsor representative for review and authorisation before being submitted in writing to the appropriate REC, and local R&D for approval prior to participants being enrolled into an amended protocol.

#### 12.2 MANAGEMENT OF PROTOCOL NON COMPLIANCE

Prospective protocol deviations, i.e. protocol waivers, will not be approved by the sponsors and therefore will not be implemented, except where necessary to eliminate an immediate hazard to study participants. If this necessitates a subsequent protocol amendment, this should be submitted to the REC, and local R&D for review and approval if appropriate.

Protocol deviations will be recorded in a protocol deviation log and logs will be submitted to the sponsors every 3 months. Each protocol violation will be reported to the sponsor within 3 days of becoming aware of the violation. All protocol deviation logs and violation forms should be emailed to

Deviations and violations are non-compliance events discovered after the event has occurred. Deviation logs will be maintained for each site in multi-centre studies. An alternative frequency of deviation log submission to the sponsors may be agreed in writing with the sponsors.

#### 12.3 SERIOUS BREACH REQUIREMENTS

A serious breach is a breach which is likely to effect to a significant degree:

(a) the safety or physical or mental integrity of the participants of the study; or

(b) the scientific value of the study.

If a potential serious breach is identified by the Chief Investigator or delegates, the co-sponsors must be notified within 24 hours. It is the responsibility of the co-sponsors to assess the impact of the breach on the scientific value of the study, to determine whether the incident constitutes a serious breach and report to research ethics committees as necessary.

#### 12.4 STUDY RECORD RETENTION

All study documentation will be kept for a minimum of 3 years from the protocol defined end of study point. When the minimum retention period has elapsed, study documentation will not be destroyed without permission from the sponsor.

#### 12.5 END OF STUDY

The end of study is defined as the last participant's last visit (anticipated September 2019).

The Investigators or the co-sponsors have the right at any time to terminate the study for clinical or administrative reasons.

The end of the study will be reported to the REC, and R&D Office and co-sponsors within 90 days, or 15 days if the study is terminated prematurely. The Investigators will inform participants of the premature study closure and ensure that the appropriate follow up is arranged for all participants involved. End of study notification will be reported to the co-sponsors via email to.

A summary report of the study will be provided to the REC within 1 year of the end of the study.

#### 12.6 INSURANCE AND INDEMNITY

The co-sponsors are responsible for ensuring proper provision has been made for insurance or indemnity to cover their liability and the liability of the Chief Investigator and staff.

The following arrangements are in place to fulfil the co-sponsors' responsibilities:

- The Protocol has been designed by the Chief Investigator and researchers employed by the University and collaborators. The University has insurance in place (which includes no-fault compensation) for negligent harm caused by poor protocol design by the Chief Investigator and researchers employed by the University.
- Sites participating in the study will be liable for clinical negligence and other negligent harm to individuals taking part in the study and covered by the duty of care owed to them by the sites concerned. The co-sponsors require individual sites participating in the study to arrange for their own insurance or indemnity in respect of these liabilities.
- Sites which are part of the United Kingdom's National Health Service will have the benefit of NHS Indemnity.

### 13 REPORTING, PUBLICATIONS AND NOTIFICATION OF RESULTS

#### 13.1 AUTHORSHIP POLICY

Ownership of the data arising from this study resides with the study team. Aggregated results of the study will be fed back to participants and published with Open Access. The UKRI Open Access Policy and MRC Data Sharing Policy will be followed to maximize opportunities for data linkage and interoperability (https://mrc.ukri.org/documents/pdf/mrc-data-sharing-policy/). Data preservation will comply with the UK Data Service Preservation Policy (https://data-archive.ac.uk/media/514523/cd062-preservationpolicy.pdf). Metadata will be provided according to DataCite Metadata Schema (https://schema.datacite.org/) allowing datasets to be discovered, interpreted, and used by others. Human datasets will be de-identified via the Safe Harbor Method (https://www.hhs.gov/hipaa/forprofessionals/privacy/special-topic s/deidentification/index.html#standard) prior to deposit. Positive and negative research findings will be published with Open Access and according to FORCE11 Data Citation Principles (https://www.force11.org/group/joint-declaration-data-citation-principles-final). Datasets of value to the research community will be shared immediately upon publication. For data derived from human participants, intention to publish and re-use this data for research purposes will be explained during the consenting procedure. Datasets will be uploaded to recognised repositories such that the data can be preserved and curated beyond the lifetime of the funded study. Anonymised and averaged MRS-derived $T_{Br}$ maps will be deposited securely in NeuroVault (http://neurovault.org) under a Creative Commons Public Domain Dedication License (CC0), or will be made available via the University of Edinburgh Datashare (https://datashare.is.ed.ac.uk). Open Access manuscripts will detail DOIs and repositories of deposited datasets, including details of software (version/accessibility) required to view/re-use data, or to replicate analyses. Data derived from human participants will be managed and shared according to terms described in the Participant Consent Form, in compliance with the revised GDPR (https://www.eugdpr.org). Terms will include provision for sharing anonymised and aggregate data that maximizes its value for wider research use, whilst retaining participant confidentiality. Current and potential future risks of data use will be explained during the consenting procedure. UKRI Knowledge Exchange Principles will be followed.

### 14 REFERENCES

Baker FC, Waner JI, Viera EF, Taylor SR, Driver HS, Mitchell D (2001) Sleep and 24 hour body temperatures: a comparison in young men, naturally cycling women and women taking hormonal contraceptives. *J Physiol* 530(Pt 3):565–574.

Booth TC, Jackson A, Wardlaw JM, Taylor SA, Waldman AD (2010) Incidental findings found in “healthy” volunteers during imaging performed for research: current legal and ethical implications. *Br J Radiol* 83:456-65.

Booth TC, Waldman AD, Wardlaw JM, Taylor SA, Jackson A (2012) Management of incidental findings during imaging research in “healthy” volunteers: current UK practice. *Br J Radiol* 85:11-21.

Brown MA (2002) Time-domain combination of MR spectroscopy data acquired using phased-array coils. *Magn Reson Med* 52:1207–1213.

Fischer D, Lombardi DA, Marucci-Wellman H, Roenneberg T (2017) Chronotypes in the US – influence of age and sex. *PLoS One* 12(6): e0178782.

Circadian Brain Temperature Study

Medical Research Council

Academic and Clinical Central Office for Research and Development

Morris Z, Whiteley WN, Longstreth WT Jr, Weber F, Lee YC, Alphs H, Ladd SC, Warlow C, Wardlaw JM, Al-Shahi Salman R (2009) Incidental findings on brain magnetic resonance imaging: systematic review and meta-analysis. *BMJ* 339:b3016.

RC Little *et al.* SAS for mixed models, Second Edition, Chapter 12.

Rzechorzek NM, Rhodes J, Andrews P, O'Neill J (2018) Circadian brain temperature fluctuation in patients with traumatic brain injury. *EuroNeuro2018,* Brussels, Belgium.

Scheenen TWJ, Klomp DWJ, Wijnen JP, Heerschap A (2008) Short echo time H-1-MRSI of the human brain at 3T with minimal chemical shift displacement errors using adiabatic refocusing pulses. *Magn Reson Med* 59:1–6.

Thrippleton MJ, Parikh J, Harris BA, Hammer SJ, Semple SI, Andrews PJ, Wardlaw JM, Marshall I (2014) Reliability of MRSI brain temperature mapping at 1.5 and 3 T. *NMR Biomed* 27:183-90.
