## Supplementary Appendix 3 for "Diurnal brain temperature rhythms and mortality after brain injury: a prospective and retrospective cohort study"

| \\Study Protocols\BRAIN\Other\CiBraT_E192051\t2_space_v4 |
|---|
| TA: 3:42 PM: REF Voxel size: 0.9×0.9×0.9 mmPAT: 4 Rel. SNR: 1.00 : spcR |

### Properties

| | |
|---|---|
| Prio recon | Off |
| Load images to viewer | On |
| Inline movie | Off |
| Auto store images | On |
| Load images to stamp segments | On |
| Load images to graphic segments | Off |
| Auto open inline display | Off |
| Auto close inline display | Off |
| Start measurement without further preparation | Off |
| Wait for user to start | Off |
| Start measurements | Single measurement |

### Routine

| | |
|---|---|
| Slab group | 1 |
| Slabs | 1 |
| Position | Isocenter |
| Orientation | Transversal |
| Phase enc. dir. | R >> L |
| AutoAlign | Head > Brain |
| Phase oversampling | 0 % |
| Slice oversampling | 18.2 % |
| Slices per slab | 176 |
| FoV read | 240 mm |
| FoV phase | 100.0 % |
| Slice thickness | 0.90 mm |
| TR | 3200 ms |
| TE | 408 ms |
| Averages | 1.4 |
| Concatenations | 1 |
| Filter | Raw filter, Distortion Corr.(2D), Prescan Normalize |
| Coil elements | HEA;HEP |

### Contrast - Common

| | |
|---|---|
| TR | 3200 ms |
| TE | 408 ms |
| MTC | Off |
| Magn. preparation | None |
| Fat suppr. | None |
| Blood suppr. | Off |
| Restore magn. | On |

### Contrast - Dynamic

| | |
|---|---|
| Averages | 1.4 |
| Reconstruction | Magnitude |
| Measurements | 1 |
| Multiple series | Each measurement |

### Resolution - Common

| | |
|---|---|
| FoV read | 240 mm |
| FoV phase | 100.0 % |
| Slice thickness | 0.90 mm |
| Base resolution | 256 |
| Phase resolution | 100 % |
| Slice resolution | 100 % |
| Phase partial Fourier | Allowed |
| Slice partial Fourier | Off |
| Interpolation | Off |

### Resolution - iPAT

| | |
|---|---|
| PAT mode | GRAPPA |
| Accel. factor PE | 2 |
| Ref. lines PE | 24 |
| Accel. factor 3D | 2 |
| Ref. lines 3D | 24 |
| Reference scan mode | Integrated |

### Resolution - Filter Image

| | |
|---|---|
| Image Filter | Off |
| Distortion Corr. | On |
| Mode | 2D |
| Unfiltered images | On |
| Prescan Normalize | On |
| Unfiltered images | Off |
| Normalize | Off |
| B1 filter | Off |

### Resolution - Filter Rawdata

| | |
|---|---|
| Raw filter | On |
| Elliptical filter | Off |

### Geometry - Common

| | |
|---|---|
| Slab group | 1 |
| Slabs | 1 |
| Position | Isocenter |
| Orientation | Transversal |
| Phase enc. dir. | R >> L |
| Slice oversampling | 18.2 % |
| Slices per slab | 176 |
| FoV read | 240 mm |
| FoV phase | 100.0 % |
| Slice thickness | 0.90 mm |
| TR | 3200 ms |
| Series | Interleaved |
| Concatenations | 1 |

### Geometry - AutoAlign

| | |
|---|---|
| Slab group | 1 |
| Position | Isocenter |
| Orientation | Transversal |
| Phase enc. dir. | R >> L |
| AutoAlign | Head > Brain |
| Initial Position | Isocenter |
| L | 0.0 mm |
| P | 0.0 mm |
| H | 0.0 mm |
| Initial Rotation | 90.00 deg |
| Initial Orientation | Transversal |

### Geometry - Saturation

| | |
|---|---|
| Fat suppr. | None |
| Restore magn. | On |
| Special sat. | None |

### Geometry - Navigator

### Geometry - Tim Planning Suite

| | |
|---|---|
| Set-n-Go Protocol | Off |
| Table position | H |
| Table position | 0 mm |

### Geometry - Tim Planning Suite

| | |
|---|---|
| Inline Composing | Off |

### System - Miscellaneous

| | |
|---|---|
| Positioning mode | REF |
| Table position | H |
| Table position | 0 mm |
| MSMA | S - C - T |
| Sagittal | R >> L |
| Coronal | A >> P |
| Transversal | F >> H |
| Coil Combine Mode | Adaptive Combine |
| Save uncombined | Off |
| Matrix Optimization | Performance |
| AutoAlign | Head > Brain |
| Coil Select Mode | On - AutoCoilSelect |

### System - Adjustments

| | |
|---|---|
| B0 Shim mode | Tune up |
| B1 Shim mode | TrueForm |
| Adjust with body coil | Off |
| Confirm freq. adjustment | Off |
| Assume Dominant Fat | Off |
| Assume Silicone | Off |
| Adjustment Tolerance | Auto |

### System - Adjust Volume

| | |
|---|---|
| Position | Isocenter |
| Orientation | Transversal |
| Rotation | 0.00 deg |
| A >> P | 263 mm |
| R >> L | 350 mm |
| F >> H | 350 mm |
| Reset | Off |

### System - pTx Volumes

| | |
|---|---|
| B1 Shim mode | TrueForm |
| Excitation | Slab-sel. |

### System - Tx/Rx

| | |
|---|---|
| Frequency 1H | 123.244480 MHz |
| Correction factor | 1 |
| Gain | High |
| Img. Scale Cor. | 1.000 |
| Reset | Off |
| ? Ref. amplitude 1H | 0.000 V |

### Physio - Signal1

| | |
|---|---|
| 1st Signal/Mode | None |
| Trigger delay | 0 ms |
| TR | 3200 ms |
| Concatenations | 1 |

### Physio - Cardiac

| | |
|---|---|
| Magn. preparation | None |
| Fat suppr. | None |
| Dark blood | Off |
| FoV read | 240 mm |
| FoV phase | 100.0 % |
| Phase resolution | 100 % |

### Physio - PACE

| | |
|---|---|
| Resp. control | Off |
| Concatenations | 1 |

### Inline - Common

| | |
|---|---|
| Subtract | Off |
| Measurements | 1 |
| StdDev | Off |
| Save original images | On |

### Inline - MIP

| | |
|---|---|
| MIP-Sag | Off |
| MIP-Cor | Off |
| MIP-Tra | Off |
| MIP-Time | Off |
| Save original images | On |

### Inline - Composing

| | |
|---|---|
| Inline Composing | Off |
| Distortion Corr. | On |
| Mode | 2D |
| Unfiltered images | On |

### Sequence - Part 1

| | |
|---|---|
| Introduction | On |
| Dimension | 3D |
| Elliptical scanning | Off |
| Reordering | Linear |
| Flow comp. | No |
| Echo spacing | 3.61 ms |
| Adiabatic-mode | Off |
| Bandwidth | 723 Hz/Px |

### Sequence - Part 2

| | |
|---|---|
| Echo train duration | 910 ms |
| RF pulse type | Normal |
| Gradient mode | Fast |
| Excitation | Slab-sel. |
| Flip angle mode | T2 var |
| Turbo factor | 282 |

### Sequence - Assistant

| | |
|---|---|
| Allowed delay | 30 s |

| \\Study Protocols\BRAIN\Other\CiBraT_E192051\t1_mprage_sag_p3_iso_Munich |
|---|
| TA: 3:45 PM: REF Voxel size: 1.0×1.0×1.0 mmPAT: 3 Rel. SNR: 1.00 : tfl |

### Properties

| | |
|---|---|
| Prio recon | Off |
| Load images to viewer | On |
| Inline movie | Off |
| Auto store images | On |
| Load images to stamp segments | On |
| Load images to graphic segments | Off |
| Auto open inline display | Off |
| Auto close inline display | Off |
| Start measurement without further preparation | Off |
| Wait for user to start | Off |
| Start measurements | Single measurement |

### Routine

| | |
|---|---|
| Slab group | 1 |
| Slabs | 1 |
| Dist. factor | 50 % |
| Position | Isocenter |
| Orientation | Sagittal |
| Phase enc. dir. | A >> P |
| AutoAlign | Head > Basis |
| Phase oversampling | 0 % |
| Slice oversampling | 0.0 % |
| Slices per slab | 192 |
| FoV read | 256 mm |
| FoV phase | 100.0 % |
| Slice thickness | 1.00 mm |
| TR | 2500.0 ms |
| TE | 4.37 ms |
| Averages | 1 |
| Concatenations | 1 |
| Filter | Distortion Corr.(2D), Prescan Normalize |
| Coil elements | HEA;HEP |

### Contrast - Common

| | |
|---|---|
| TR | 2500.0 ms |
| TE | 4.37 ms |
| Magn. preparation | Non-sel. IR |
| TI | 1100 ms |
| Flip angle | 7 deg |
| Fat suppr. | Water excit. fast |
| Water suppr. | None |

### Contrast - Dynamic

| | |
|---|---|
| Averages | 1 |
| Averaging mode | Long term |
| Reconstruction | Magnitude |
| Measurements | 1 |
| Multiple series | Each measurement |

### Resolution - Common

| | |
|---|---|
| FoV read | 256 mm |
| FoV phase | 100.0 % |
| Slice thickness | 1.00 mm |
| Base resolution | 256 |
| Phase resolution | 100 % |
| Slice resolution | 100 % |
| Phase partial Fourier | 7/8 |
| Slice partial Fourier | Off |

### Resolution - Common

| | |
|---|---|
| Interpolation | Off |

### Resolution - iPAT

| | |
|---|---|
| PAT mode | GRAPPA |
| Accel. factor PE | 3 |
| Ref. lines PE | 24 |
| Accel. factor 3D | 1 |
| Reference scan mode | Integrated |

### Resolution - Filter Image

| | |
|---|---|
| Image Filter | Off |
| Distortion Corr. | On |
| Mode | 2D |
| Unfiltered images | On |
| Prescan Normalize | On |
| Unfiltered images | Off |
| Normalize | Off |
| B1 filter | Off |

### Resolution - Filter Rawdata

| | |
|---|---|
| Raw filter | Off |
| Elliptical filter | Off |

### Geometry - Common

| | |
|---|---|
| Slab group | 1 |
| Slabs | 1 |
| Dist. factor | 50 % |
| Position | Isocenter |
| Orientation | Sagittal |
| Phase enc. dir. | A >> P |
| Slice oversampling | 0.0 % |
| Slices per slab | 192 |
| FoV read | 256 mm |
| FoV phase | 100.0 % |
| Slice thickness | 1.00 mm |
| TR | 2500.0 ms |
| Multi-slice mode | Single shot |
| Series | Interleaved |
| Concatenations | 1 |

### Geometry - AutoAlign

| | |
|---|---|
| Slab group | 1 |
| Position | Isocenter |
| Orientation | Sagittal |
| Phase enc. dir. | A >> P |
| AutoAlign | Head > Basis |
| Initial Position | Isocenter |
| L | 0.0 mm |
| P | 0.0 mm |
| H | 0.0 mm |
| Initial Rotation | 0.00 deg |
| Initial Orientation | Sagittal |

### Geometry - Navigator

### Geometry - Tim Planning Suite

| | |
|---|---|
| Set-n-Go Protocol | Off |
| Table position | H |
| Table position | 0 mm |
| Inline Composing | Off |

### System - Miscellaneous

| | |
|---|---|
| Positioning mode | REF |
| Table position | H |
| Table position | 0 mm |
| MSMA | S - C - T |
| Sagittal | R >> L |
| Coronal | A >> P |
| Transversal | F >> H |
| Coil Combine Mode | Adaptive Combine |
| Save uncombined | Off |
| Matrix Optimization | Off |
| AutoAlign | Head > Basis |
| Coil Select Mode | On - AutoCoilSelect |

### System - Adjustments

| | |
|---|---|
| B0 Shim mode | Standard |
| B1 Shim mode | TrueForm |
| Adjust with body coil | Off |
| Confirm freq. adjustment | Off |
| Assume Dominant Fat | Off |
| Assume Silicone | Off |
| Adjustment Tolerance | Auto |

### System - Adjust Volume

| | |
|---|---|
| Position | Isocenter |
| Orientation | Sagittal |
| Rotation | 0.00 deg |
| A >> P | 256 mm |
| F >> H | 256 mm |
| R >> L | 192 mm |
| Reset | Off |

### System - pTx Volumes

| | |
|---|---|
| B1 Shim mode | TrueForm |
| Excitation | Non-sel. |

### System - Tx/Rx

| | |
|---|---|
| Frequency 1H | 123.244480 MHz |
| Correction factor | 1 |
| Gain | Low |
| Img. Scale Cor. | 1.000 |
| Reset | Off |
| ? Ref. amplitude 1H | 0.000 V |

### Physio - Signal1

| | |
|---|---|
| 1st Signal/Mode | None |
| TR | 2500.0 ms |
| Concatenations | 1 |

### Physio - Cardiac

| | |
|---|---|
| Magn. preparation | Non-sel. IR |
| TI | 1100 ms |
| Fat suppr. | Water excit. fast |
| Dark blood | Off |
| FoV read | 256 mm |
| FoV phase | 100.0 % |
| Phase resolution | 100 % |

### Physio - PACE

| | |
|---|---|
| Resp. control | Off |
| Concatenations | 1 |

### Inline - Common

| | |
|---|---|
| Subtract | Off |

### Inline - Common

| | |
|---|---|
| Measurements | 1 |
| StdDev | Off |
| Save original images | On |

### Inline - MIP

| | |
|---|---|
| MIP-Sag | Off |
| MIP-Cor | Off |
| MIP-Tra | Off |
| MIP-Time | Off |
| Save original images | On |

### Inline - Composing

| | |
|---|---|
| Inline Composing | Off |
| Distortion Corr. | On |
| Mode | 2D |
| Unfiltered images | On |

### Inline - MapIt

| | |
|---|---|
| Save original images | On |
| MapIt | None |
| Flip angle | 7 deg |
| Measurements | 1 |
| TR | 2500.0 ms |
| TE | 4.37 ms |

### Sequence - Part 1

| | |
|---|---|
| Introduction | Off |
| Dimension | 3D |
| Elliptical scanning | Off |
| Reordering | Linear |
| Asymmetric echo | Off |
| Flow comp. | No |
| Multi-slice mode | Single shot |
| Echo spacing | 11.1 ms |
| Bandwidth | 140 Hz/Px |

### Sequence - Part 2

| | |
|---|---|
| RF pulse type | Fast |
| Gradient mode | Fast |
| Excitation | Non-sel. |
| RF spoiling | On |
| Incr. Gradient spoiling | Off |
| Turbo factor | 192 |

### Sequence - Assistant

| | |
|---|---|
| Mode | Off |

### \\Study Protocols\BRAIN\Other\CiBraT_E192051\csi_centrum_semiovale_TBr_144

TA: 4:52 PM: REF Voxel size: 10.0×10.0×10.0 mmRel. SNR: 1.00 : csislsr

#### Properties

| | |
|---|---|
| Prio recon | Off |
| Load images to viewer | On |
| Inline movie | Off |
| Auto store images | On |
| Load images to stamp segments | Off |
| Load images to graphic segments | Off |
| Auto open inline display | Off |
| Auto close inline display | Off |
| Start measurement without further preparation | Off |
| Wait for user to start | Off |
| Start measurements | Single measurement |

#### Routine

| | |
|---|---|
| Position | L0.3 A12.2 H15.4 mm |
| Orientation | T > C-2.8 > S-2.4 |
| Rotation | 1 deg |
| Slices | 1 |
| Vol A >> P | 100 mm |
| Vol R >> L | 90 mm |
| FoV A >> P | 160 mm |
| FoV R >> L | 160 mm |
| Thickness F >> H | 10 mm |
| TR | 1200 ms |
| TE | 144 ms |
| Averages | 3 |
| Filter | Prescan Normalize, Hamming |
| Coil elements | HEA;HEP |

#### Contrast

| | |
|---|---|
| TR | 1200 ms |
| TE | 144 ms |
| Averages | 3 |
| Averaging mode | Long term |
| Flip angle | 65 deg |
| Water suppr. | Weak water suppr. |
| Water suppr. BW | 50 Hz |
| Measurements | 1 |

#### Resolution - Common

| | |
|---|---|
| FoV R >> L | 160 mm |
| FoV A >> P | 160 mm |
| Thickness F >> H | 10 mm |
| Scan res. R >> L | 16 |
| Scan res. A >> P | 16 |
| Interpol. res. R >> L | 16 |
| Interpol. res. A >> P | 16 |
| Hamming | On |
| Width | 50 |
| Prescan Normalize | On |
| Vector size | 1024 |

#### Geometry - Common

| | |
|---|---|
| Position | L0.3 A12.2 H15.4 mm |
| Orientation | T > C-2.8 > S-2.4 |
| Rotation | 1 deg |
| FoV R >> L | 160 mm |
| FoV A >> P | 160 mm |
| Thickness F >> H | 10 mm |

#### Geometry - Common

| | |
|---|---|
| Vol R >> L | 90 mm |
| Vol A >> P | 100 mm |
| Sat. region | 1 |
| Thickness | 40 mm |
| Position | R67.7 P0.2 H6.1 mm |
| Orientation | S > T5.2 > C0.2 |
| Sat. delta frequ. | -3.40 ppm |
| Sat. region | 2 |
| Thickness | 40 mm |
| Position | L67.1 A1.0 F2.4 mm |
| Orientation | S > T2.0 > C0.9 |
| Sat. delta frequ. | -3.40 ppm |
| Sat. region | 3 |
| Thickness | 40 mm |
| Position | L5.5 A7.2 F13.3 mm |
| Orientation | T > C-2.7 > S-2.5 |
| Sat. delta frequ. | -3.40 ppm |
| Sat. region | 4 |
| Thickness | 40 mm |
| Position | R1.6 A84.8 H4.3 mm |
| Orientation | C > T2.9 > S-1.0 |
| Sat. delta frequ. | -3.40 ppm |
| Sat. region | 5 |
| Thickness | 40 mm |
| Position | L0.9 P58.8 F3.2 mm |
| Orientation | C > T3.1 > S-0.9 |
| Sat. delta frequ. | -3.40 ppm |
| Sat. region | 6 |
| Thickness | 40 mm |
| Position | L7.9 A4.3 H42.4 mm |
| Orientation | T > C-3.0 > S-2.5 |
| Sat. delta frequ. | -3.40 ppm |

#### Geometry - AutoAlign

| | |
|---|---|
| Slice group | 1 |
| Position | L0.3 A12.2 H15.4 mm |
| Orientation | T > C-2.8 > S-2.4 |
| Phase enc. dir. | A >> P |
| AutoAlign | Head > Brain |
| Initial Position | L0.7 A0.5 H33.7 |
| L | 0.7 mm |
| A | 0.5 mm |
| H | 33.7 mm |
| Initial Rotation | 91.75 deg |
| Initial Orientation | Transversal |

#### System - Miscellaneous

| | |
|---|---|
| Positioning mode | REF |
| Table position | F |
| Table position | 23 mm |
| MSMA | S - C - T |
| Sagittal | R >> L |
| Coronal | A >> P |
| Transversal | F >> H |
| Save uncombined | Off |
| AutoAlign | Head > Brain |
| Coil Select Mode | Default |

#### System - Adjustments

| | |
|---|---|
| B0 Shim mode | Brain |

### System - Adjustments

| | |
|---|---|
| B1 Shim mode | TrueForm |
| Adj. water suppr. | On |
| Adjust with body coil | Off |
| Confirm freq. adjustment | On |
| Only after freq. change | On |
| Assume Dominant Fat | Off |
| Assume Silicone | Off |
| Adjustment Tolerance | Auto |

### System - Adjust Volume

| | |
|---|---|
| Position | L0.3 A12.2 H15.4 mm |
| Orientation | T > C-2.8 > S-2.4 |
| Rotation | 91.00 deg |
| R >> L | 90 mm |
| A >> P | 100 mm |
| F >> H | 10 mm |
| Reset | Off |

### System - pTx Volumes

| | |
|---|---|
| B1 Shim mode | TrueForm |

### System - Tx/Rx

| | |
|---|---|
| Frequency 1H | 123.244480 MHz |
| Gain | High |
| Img. Scale Cor. | 1.000 |
| Reset | Off |
| ? Ref. amplitude 1H | 0.000 V |

### Sequence - Common

| | |
|---|---|
| Preparation scans | 4 |
| Dimension | 2D |
| Delta frequency | -2.70 ppm |
| Phase encoding | Weighted |
| Bandwidth | 2000 Hz |
| Acquisition duration | 512 ms |
| Remove oversampling | Off |

| \\Study Protocols\BRAIN\Other\CiBraT_E192051\svs_hypothalamus_TBr_144 |
|---|
| TA: 5:13 PM: REF Vol: 10 ×20 ×10 mmRel. SNR: 1.00 : svs_se |

### Properties

| | |
|---|---|
| Prio recon | Off |
| Load images to viewer | On |
| Inline movie | Off |
| Auto store images | On |
| Load images to stamp segments | Off |
| Load images to graphic segments | Off |
| Auto open inline display | Off |
| Auto close inline display | Off |
| Start measurement without further preparation | Off |
| Wait for user to start | Off |
| Start measurements | Single measurement |

### Routine

| | |
|---|---|
| Position | Isocenter |
| Orientation | Transversal |
| Rotation | 0 deg |
| Vol R >> L | 20 mm |
| Vol R >> L | 20 mm |
| Vol F >> H | 10 mm |
| TR | 1200 ms |
| TE | 144 ms |
| Averages | 256 |
| Filter | Prescan Normalize |
| Coil elements | HE1-4 |

### Contrast

| | |
|---|---|
| TR | 1200 ms |
| TE | 144 ms |
| Averages | 256 |
| Flip angle | 65 deg |
| Water suppr. | Weak water suppr. |
| Water suppr. BW | 50 Hz |
| Spectral suppr. | None |
| Measurements | 1 |

### Resolution - Common

| | |
|---|---|
| Prescan Normalize | On |
| Vector size | 1024 |

### Geometry - Common

| | |
|---|---|
| Position | Isocenter |
| Orientation | Transversal |
| Rotation | 0 deg |
| Vol R >> L | 20 mm |
| Vol A >> P | 10 mm |
| Vol F >> H | 10 mm |

### Geometry - AutoAlign

| | |
|---|---|
| AutoAlign | --- |
| Initial Position | Isocenter |
| L | 0 mm |
| P | 0 mm |
| H | 0 mm |
| Initial Rotation | 0.00 deg |
| Initial Orientation | Transversal |

### Geometry - Navigator

### System - Miscellaneous

| | |
|---|---|
| Positioning mode | REF |
| Table position | H |
| Table position | 0 mm |
| MSMA | S - C - T |
| Sagittal | R >> L |
| Coronal | A >> P |
| Transversal | F >> H |
| Save uncombined | Off |
| Save single averages | Off |
| AutoAlign | --- |
| Coil Select Mode | Default |

### System - Adjustments

| | |
|---|---|
| B0 Shim mode | Brain |
| B1 Shim mode | TrueForm |
| Adj. water suppr. | On |
| Adjust with body coil | Off |
| Confirm freq. adjustment | On |
| Only after freq. change | On |
| Assume Dominant Fat | Off |
| Assume Silicone | Off |
| Adjustment Tolerance | Auto |

### System - Adjust Volume

| | |
|---|---|
| Position | Isocenter |
| Orientation | Transversal |
| Rotation | 0.00 deg |
| A >> P | 10 mm |
| R >> L | 20 mm |
| F >> H | 10 mm |
| Reset | Off |

### System - pTx Volumes

| | |
|---|---|
| B1 Shim mode | TrueForm |

### System - Tx/Rx

| | |
|---|---|
| Frequency 1H | 123.244480 MHz |
| Gain | High |
| Img. Scale Cor. | 1.000 |
| Reset | Off |
| ? Ref. amplitude 1H | 0.000 V |

### Physio - Signal1

| | |
|---|---|
| 1st Signal/Mode | None |
| TR | 1200 ms |

### Physio - PACE

| | |
|---|---|
| Resp. control | Off |

### Sequence - Common

| | |
|---|---|
| Preparation scans | 4 |
| Delta frequency | -2.3 ppm |
| Ref. scan mode | Save all |
| No. of ref. scans | 1 |
| Phase cycling | Auto |
| Bandwidth | 2000 Hz |
| Acquisition duration | 512 ms |
| Remove oversampling | Off |

| \\Study Protocols\BRAIN\Other\CiBraT_E192051\svs_thalamus_TBr_144 |
|---|
| TA: 2:40 PM: REF Vol: 15 ×15 ×15 mmRel. SNR: 1.00 : svs_se |

### Properties

| | |
|---|---|
| Prio recon | Off |
| Load images to viewer | On |
| Inline movie | Off |
| Auto store images | On |
| Load images to stamp segments | Off |
| Load images to graphic segments | Off |
| Auto open inline display | Off |
| Auto close inline display | Off |
| Start measurement without further preparation | Off |
| Wait for user to start | Off |
| Start measurements | Single measurement |

### Routine

| | |
|---|---|
| Position | Isocenter |
| Orientation | Transversal |
| Rotation | 0 deg |
| Vol R >> L | 15 mm |
| Vol R >> L | 15 mm |
| Vol F >> H | 15 mm |
| TR | 1200 ms |
| TE | 144 ms |
| Averages | 128 |
| Filter | Prescan Normalize |
| Coil elements | HE1-4 |

### Contrast

| | |
|---|---|
| TR | 1200 ms |
| TE | 144 ms |
| Averages | 128 |
| Flip angle | 65 deg |
| Water suppr. | Weak water suppr. |
| Water suppr. BW | 50 Hz |
| Spectral suppr. | None |
| Measurements | 1 |

### Resolution - Common

| | |
|---|---|
| Prescan Normalize | On |
| Vector size | 1024 |

### Geometry - Common

| | |
|---|---|
| Position | Isocenter |
| Orientation | Transversal |
| Rotation | 0 deg |
| Vol R >> L | 15 mm |
| Vol A >> P | 15 mm |
| Vol F >> H | 15 mm |

### Geometry - AutoAlign

| | |
|---|---|
| AutoAlign | --- |
| Initial Position | Isocenter |
| L | 0 mm |
| P | 0 mm |
| H | 0 mm |
| Initial Rotation | 0.00 deg |
| Initial Orientation | Transversal |

### Geometry - Navigator

### System - Miscellaneous

| | |
|---|---|
| Positioning mode | REF |
| Table position | H |
| Table position | 0 mm |
| MSMA | S - C - T |
| Sagittal | R >> L |
| Coronal | A >> P |
| Transversal | F >> H |
| Save uncombined | Off |
| Save single averages | Off |
| AutoAlign | --- |
| Coil Select Mode | Default |

### System - Adjustments

| | |
|---|---|
| B0 Shim mode | Brain |
| B1 Shim mode | TrueForm |
| Adj. water suppr. | On |
| Adjust with body coil | Off |
| Confirm freq. adjustment | On |
| Only after freq. change | On |
| Assume Dominant Fat | Off |
| Assume Silicone | Off |
| Adjustment Tolerance | Auto |

### System - Adjust Volume

| | |
|---|---|
| Position | Isocenter |
| Orientation | Transversal |
| Rotation | 0.00 deg |
| A >> P | 15 mm |
| R >> L | 15 mm |
| F >> H | 15 mm |
| Reset | Off |

### System - pTx Volumes

| | |
|---|---|
| B1 Shim mode | TrueForm |

### System - Tx/Rx

| | |
|---|---|
| Frequency 1H | 123.244480 MHz |
| Gain | High |
| Img. Scale Cor. | 1.000 |
| Reset | Off |
| ? Ref. amplitude 1H | 0.000 V |

### Physio - Signal1

| | |
|---|---|
| 1st Signal/Mode | None |
| TR | 1200 ms |

### Physio - PACE

| | |
|---|---|
| Resp. control | Off |

### Sequence - Common

| | |
|---|---|
| Preparation scans | 4 |
| Delta frequency | -2.3 ppm |
| Ref. scan mode | Save all |
| No. of ref. scans | 1 |
| Phase cycling | Auto |
| Bandwidth | 2000 Hz |
| Acquisition duration | 512 ms |
| Remove oversampling | Off |
