## Supplementary Appendix 4 for "Diurnal brain temperature rhythms and mortality after brain injury: a prospective and retrospective cohort study"

Academic and Clinical Central Office for Research and Development

| | | | | | |
|---|---|---|---|---|---|
| **IRAS Number:** | 244533 | **REC Number:** | 18-HV-045 | **R&D Number:** | 2019/0133 |
| **Sponsor Number:** | AC 18038 | **Site ID:** | E192051 | **Study Acronym:** | CiBraT |
| **NIHR CPMS ID:** | 42644 | **Participant ID:** | CiBraT_ | **Participant Initials:** | |

### Study Participant Data Form (Case Report Form)

Abbreviations: Y = yes, N = no, NA = not applicable, NR = not recruited, NK = not known, ND = not done, WD = withdrawn, RS = rescheduled, AE = adverse event, DC = declined

#### Study Details

| | | | |
|---|---|---|---|
| Study title | Can we measure a diurnal shift in brain temperature in healthy human volunteers using Magnetic Resonance Spectroscopy (MRS)? | | |
| Short study title | Circadian Brain Temperature (CiBraT) Study | | |
| Chief Investigator | Dr Nina Rzechorzek (also Principal Investigator) | | |
| Medical Statistician | Dr Francesca Chappell | Study Manager | Dr Duncan Martin |
| Local Collaborator | Prof Ian Marshall | Medical Physicist | Dr Michael J Thrippleton |
| Neuroradiologist | Dr Grant Mair | Location | Edinburgh Imaging (RIE) Facility |
| Study design | Prospective, single site, cohort study in healthy volunteers | | |
| Jisc URL | https://mrc.onlinesurveys.ac.uk/cibrat | | |

*I declare that this Section is complete and accurate to the best of my knowledge*

| | | | |
|---|---|---|---|
| Chief Investigator | [signature] MRCVS | Date | DD / MMM / YYYY 2019 |

#### Participant Details

| | | | |
|---|---|---|---|
| Initials | | Sex | Male / Female |
| Age | YY / MM | Study ID | C i B r a T _ |
| Postcode | | Within 5 miles? | Y / N |
| Jisc response date | DD / MMM / YYYY | Urine kit needed? | Y / N / NA |

*I declare that this Section is complete and accurate to the best of my knowledge*

| | | | |
|---|---|---|---|
| Chief Investigator | [signature] MRCVS | Date | DD / MMM / YYYY 2019 |

Academic and Clinical Central Office for Research and Development

| IRAS Number: | 244533 | REC Number: | 18-HV-045 | R&D Number: | 2019/0133 |
|---|---|---|---|---|---|
| Sponsor Number: | AC 18038 | Site ID: | E192051 | Study Acronym: | CiBraT |
| NIHR CPMS ID: | 42644 | Participant ID: | CiBraT_ | Participant Initials: | |

#### NHS Details

| | | | |
|---|---|---|---|
| GP name | | | |
| GP address | | | |
| GP notified? | Y / N | GP letter date | DD MMM 2019 (YYYY) |
| GP concerns? | Y / N / NK | Cycle control: Pill / Patch / NA | Admin time: HH / NA |
| CHI number | | Brand/NA | |

*I declare that this Section is complete and accurate to the best of my knowledge*

| | | | |
|---|---|---|---|
| Chief Investigator | [signature] | Date | DD MMM 2019 (YYYY) |

#### Consenting Interview (Pre-Scan Visit)

| | | | |
|---|---|---|---|
| Date | DD MMM 2019 (YYYY) | Time | HH MM |
| Location | | | |
| Data entry | Dr Nina Rzechorzek | Willingness to participate rechecked? | Y / N / NR |

| | |
|---|---|
| Has participant read and understood PIS, CTPF, DPIS? | Y / N |
| Three copies of CTPF completed and signed? | Y / N / NR |
| Hard copies of PIS, signed CTPF, and DPIS given to participant? | Y / N / NR |
| Participant elected to be emailed about (a) study results and/or (b) future studies? | (a) Y / N; (b) Y / N |

| | | | |
|---|---|---|---|
| MRI checklist completed? | Y / N | Checklist queries reported to radiography team? | Y / N / NA |
| Urine Test Kit supplied? | Y / N / NA / NR | Number of days to scan date | |
| ActTrust2 device issued | | ActTrust2 participant ID | CiBraT_ |
| Scans booked? | Y / N / NA | Travel expenses received? | Y / N / NA |

*I declare that this Section is complete and accurate to the best of my knowledge*

| | | | |
|---|---|---|---|
| Chief Investigator | [signature] | Date | DD MMM 2019 (YYYY) |

Academic and Clinical Central Office for Research and Development

| IRAS Number: | 244533 | REC Number: | 18-HV-045 | R&D Number: | 2019/0133 |
|---|---|---|---|---|---|
| Sponsor Number: | AC 18038 | Site ID: | E192051 | Study Acronym: | CiBraT |
| NIHR CPMS ID: | 42644 | Participant ID: | CiBraT_ | Participant Initials: | |

#### Scan Visit 1 (Morning)

| | | | |
|---|---|---|---|
| Date | DD / MMM / YYYY: 2019 | Time | HH : MM |
| Participant arrived by 8.45am (female) or 9.15am (male)? | Y / N / NA | Room temp. | __ __ . __ |
| Location | Edinburgh Imaging (RIE) Facility (BRIC2) | | |
| Data entry | Dr Nina Rzechorzek | Radiographer | |
| Participant well? | Y / N — Any AEs? Y / N | AEs followed up? | Y / N / NA |
| Willingness checked? | Y / N / WD / RS | Actigraph removed? | Y / N |
| Number of days Actigraph worn | Free / Scheduled | Patch removed? | Y / N / NA |
| MRI checklist signed? | Y / N | Ovulation confirmed? | Y / N / NA |
| Oral temperature (°C) | __ __ . __ | Within range? | Y / N — 33.2–38.1 °C women; 35.7–37.7 °C men |
| Height (m) | __ . __ __ | Weight (kg) | __ __ __ . __ |
| BMI (kg/m²) | __ __ . __ | BMI within range (18.5-29.9)? | Y / N |
| Prohibited medications or alcohol? | Y / N | Food or caffeine after 8am? | Y / N |
| Vigorous exercise this morning? | Y / N | Eligibility confirmed? | Y / N |
| Participant changed into hospital clothing? | Y / N | All MR sequences completed? | Y / N |
| Reason for reschedule (if applicable, otherwise NA) | | | |
| Reason for withdrawal (if given, otherwise NA) | | | |
| Participant reported falling asleep during scan? | Y / N / NK | Scan duration (minutes) | |
| Meal vouchers (lunch) issued? | Y / N / DC | Visit duration (minutes) | |
| Actigraphy data transferred? | Y / N | ActTrust2 data erased? | Y / N |
| *I declare that this Section is complete and accurate to the best of my knowledge* | | | |
| Chief Investigator | [signature] | Date | DD / MMM / YYYY: 2019 |

Academic and Clinical Central Office for Research and Development

| | | | | | |
|---|---|---|---|---|---|
| **IRAS Number:** | 244533 | **REC Number:** | 18-HV-045 | **R&D Number:** | 2019/0133 |
| **Sponsor Number:** | AC 18038 | **Site ID:** | E192051 | **Study Acronym:** | CiBraT |
| **NIHR CPMS ID:** | 42644 | **Participant ID:** | CiBraT_ | **Participant Initials:** | |

#### Scan Visit 2 (Afternoon)

| | | | |
|---|---|---|---|
| Date | DD / MMM / YYYY 2019 | Time | HH / MM |
| Participant arrived by 3.45pm (female) or 4.15am (male)? | Y / N / NA | Room temp. | . |
| Location | Edinburgh Imaging (RIE) Facility (BRIC2) | | |
| Data entry | Dr Nina Rzechorzek | Radiographer | |
| Any AEs? | Y / N / NA | AEs followed up? | Y / N / NA |
| Participant reports feeling well? | Y / N | Willingness rechecked? | Y / N / WD / RS |
| MRI checklist reviewed and signed? | Y / N | Patch removed? | Y / N / NA |
| Oral temperature (°C) | . | Within range? | Y / N 33.2–38.1 °C women 35.7–37.7 °C men |
| Prohibited medications or alcohol? | Y / N | Food or caffeine beyond 12-2pm? | Y / N |
| Vigorous exercise since this morning? | Y / N | Eligibility confirmed? | Y / N |
| Participant changed into hospital clothing? | Y / N | All MR sequences completed? | Y / N |
| Reason for withdrawal (if given, otherwise NA) | | | |
| Participant reported falling asleep during scan? | Y / N / NK | Scan duration (minutes) | |
| Meal vouchers (dinner) issued? | Y / N / DC | Visit duration (minutes) | |
| *I declare that this Section is complete and accurate to the best of my knowledge* | | | |
| Chief Investigator | [signature] | Date | DD / MMM / YYYY 2019 |

accord
Academic and Clinical Central Office for Research and Development

| | | | | | |
|---|---|---|---|---|---|
| **IRAS Number:** | 244533 | **REC Number:** | 18-HV-045 | **R&D Number:** | 2019/0133 |
| **Sponsor Number:** | AC 18038 | **Site ID:** | E192051 | **Study Acronym:** | CiBraT |
| **NIHR CPMS ID:** | 42644 | **Participant ID:** | CiBraT_ | **Participant Initials:** | |

#### Scan Visit 3 (Evening)

| | | | |
|---|---|---|---|
| Date | __ __ / __ __ __ / 2 0 1 9 (DD MMM YYYY) | Time | __ __ : __ __ (HH MM) |
| Participant arrived by 10.45pm (female) or 11.15pm (male)? | Y / N / NA | Room temp. | __ __ . __ |
| Location | Edinburgh Imaging (RIE) Facility (BRIC2) | | |
| Data entry | Dr Nina Rzechorzek | Radiographer | |
| Any AEs? | Y / N / NA | AEs followed up? | Y / N / NA |
| Participant reports feeling well? | Y / N | Willingness rechecked? | Y / N / WD / RS |
| MRI checklist reviewed and signed? | Y / N | Patch removed? | Y / N / NA |
| Oral temperature (°C) | __ __ . __ | Within range? | Y / N — 33.2–38.1 °C women; 35.7–37.7 °C men |
| Medications, caffeine or alcohol? | Y / N | Food beyond 6-8pm? | Y / N |
| Vigorous exercise since last scan? | Y / N | Eligibility confirmed? | Y / N |
| Participant changed into hospital clothing? | Y / N | All MR sequences completed? | Y / N |
| Reason for withdrawal (if given, otherwise NA) | | | |
| Participant reported falling asleep during scan? | Y / N / NK | Scan duration (minutes) | __ __ |
| Notes | | Visit duration (minutes) | __ __ |
| Participant travel expenses received? | Y / N / NA | Expenses reimbursed? | Y / N / NA |
| *I declare that this Section is complete and accurate to the best of my knowledge* | | | |
| Chief Investigator | [signature] | Date | __ __ / __ __ __ / 2 0 1 9 (DD MMM YYYY) |

Academic and Clinical Central Office for Research and Development

| IRAS Number: | 244533 | REC Number: | 18-HV-045 | R&D Number: | 2019/0133 |
|---|---|---|---|---|---|
| Sponsor Number: | AC 18038 | Site ID: | E192051 | Study Acronym: | CiBraT |
| NIHR CPMS ID: | 42644 | Participant ID: | CiBraT_ | Participant Initials: | |

#### Post-scan

| | | | |
|---|---|---|---|
| Any AEs within 7 days? | Y / N / NA | AEs followed up? | Y / N / NA |
| Neuroradiology report completed? | Y / N | Any HRFs identified? | Y / N |
| HRF exclusion reported to CI? | Y / N / NA | HRFs reported to participant? | Y / N / NA |
| HRFs reported to GP? | Y / N / NA | Clinical radiology report sent to GP? | Y / N |

*I declare that this Section is complete and accurate to the best of my knowledge*

| | | | |
|---|---|---|---|
| Chief Investigator | | Date | DD / MMM / YYYY: 2019 |

#### Actigraphy chronotype data

| Night | Type* | Sleep start | Sleep end | Sleep duration (min) | Sleep midpoint | $MSF_{sc}$/$MSW_{sc}$ |
|---|---|---|---|---|---|---|
| 1 | F / S | HH MM | HH MM | | HH MM | HH MM |
| 2 | F / S | HH MM | HH MM | | HH MM | HH MM |
| 3 | F / S | HH MM | HH MM | | HH MM | HH MM |
| 4 | F / S | HH MM | HH MM | | HH MM | HH MM |
| 5 | F / S | HH MM | HH MM | | HH MM | HH MM |
| 6 | F / S | HH MM | HH MM | | HH MM | HH MM |
| 7 | F / S | HH MM | HH MM | | HH MM | HH MM |
| 8 | F / S | HH MM | HH MM | | HH MM | HH MM |
| Mean | F / S | HH MM | HH MM | | Acrophase | HH MM |
| Mean $MSF_{sc}$ | | HH MM | Mean $MSW_{sc}$ | HH MM | $SJL_{sc}$ | HH MM |

*F = free; S = scheduled (relating to wake time the following morning). $MSF_{sc}$/$MSW_{sc}$ = sleep corrected midpoint of sleep on free/work days = sleep onset on free/work days plus half of the average weekly sleep duration (all days). $SJL_{sc}$ = sleep corrected social jetlag ($MSF_{sc}$-$MSW_{sc}$ = absolute difference between sleep onset on free and work days).

*I declare that this Section is complete and accurate to the best of my knowledge*

| | | | |
|---|---|---|---|
| Chief Investigator | | Date | DD / MMM / YYYY: 2019 |

*refer to MRS database for blinded analysis and data points from each voxel

| | | | | | |
|---|---|---|---|---|---|
| **IRAS Number:** | 244533 | **REC Number:** | 18-HV-045 | **R&D Number:** | 2019/0133 |
| **Sponsor Number:** | AC 18038 | **Site ID:** | E192051 | **Study Acronym:** | CiBraT |
| **NIHR CPMS ID:** | 42644 | **Participant ID:** | CiBraT_ | **Participant Initials:** | |

#### MRS data (post-unblinding)*

| | | | |
|---|---|---|---|
| MRS data blinded by Study Manager? | Y N | | |
| Scan visit | v_1 | v_2 | v_3 |
| MRS data quality | | | |
| Mean temperature superficial voxels °C | . | . | . |
| Temperature thalamic voxel °C | . | . | . |
| Temperature hypothalamic voxel °C | . | . | . |
| Time to nearest acrophase | HH MM | HH MM | HH MM |
| Time since previous corrected sleep midpoint | HH MM | HH MM | HH MM |
| Time since average $MSF_{sc}$ | HH MM | HH MM | HH MM |

*refer to MRS database for blinded analysis and data points from each voxel

| | | | |
|---|---|---|---|
| *I declare that this Section is complete and accurate to the best of my knowledge* | | | |
| Chief Investigator | [signature] | Date | DD MMM 2019 (YYYY) |

#### Deviations or Non-compliance with Study Protocol and Serious Breaches

| | | | |
|---|---|---|---|
| Date | DD MMM YYYY | Time | HH MM |
| Nature of event | | | |
| Event requires exclusion? | Y N | | |
| Event impacts on data quality or integrity? | Y N | | |
| Event requires follow-up? | Y N | | |
| Sponsor notified? | Y N NA | | |
| GP notified? | Y N NA | | |
| *I declare that this Section is complete and accurate to the best of my knowledge* | | | |
| Chief Investigator | [signature] | Date | DD MMM 2019 (YYYY) |

accord
Academic and Clinical Central Office for Research and Development
NHS Lothian

| **IRAS Number:** | 244533 | **REC Number:** | 18-HV-045 | **R&D Number:** | 2019/0133 |
|---|---|---|---|---|---|
| **Sponsor Number:** | AC 18038 | **Site ID:** | E192051 | **Study Acronym:** | CiBraT |
| **NIHR CPMS ID:** | 42644 | **Participant ID:** | CiBraT_ | **Participant Initials:** | |

#### Participant withdrawal

| | | | |
|---|---|---|---|
| Date | DD / MMM / YYYY | | |
| Elected withdrawal or CI-determined? | | | |
| Method of notification | | | |
| Primary reason for withdrawal (if available) | | | |
| GP notified of withdrawal by CI? | | | |
| *I declare that this Section is complete and accurate to the best of my knowledge* | | | |
| Chief Investigator | [signature] | Date | DD / MMM / 2019 |

#### Adverse event (AE) recording

| | | | |
|---|---|---|---|
| Nature of event | | | |
| Start date | DD / MMM / YYYY | | |
| Stop date | DD / MMM / YYYY | | |
| Time | HH / MM / NK | | |
| Location | | | |
| Seriousness | | Severity | |
| Expectedness (only if possibly related) | | Relatedness | |
| Outcome | | | |
| CI/PI oversight | | | |
| MedDRA code | | | |
| *I declare that this Section is complete and accurate to the best of my knowledge* | | | |
| Chief Investigator | [signature] | Date | DD / MMM / 2019 |

| ***I declare that this Source Data Document is complete and accurate to the best of my knowledge*** | | | |
|---|---|---|---|
| Chief Investigator Final Sign Off | [signature] | Date | DD / MMM / 2019 |
