## Supplementary Appendix 5 for "Diurnal brain temperature rhythms and mortality after brain injury: a prospective and retrospective cohort study"

#### Analytic code for statistical models

The following code was used to generate the key statistical models for Rzechorzek et al. (2021) *‘Daily brain temperature rhythms and mortality after brain injury: a prospective and retrospective cohort study’*. Generalized linear and linear mixed modelling were performed using R version 3.6.3 (R Core Team, 2020) and the *lme4* (v1.1–23; Bates et al. 2020), *effects* (v4.1-4; Fox et al. 2019), *afex* (Singmann et al. 2020), *Matrix* (v1.2-18; Bates et al. 2019), *Cairo* (v1.5-12.2; Urbanek and Horner 2020), *yarrr* (v0.1.5; Phillips 2017), and *car* (v3.0-8; Fox et al. 2020) packages.

##### Linear mixed model for oral temperature in healthy volunteers

```
OralTemp = read.table("OralTemp.txt", header=TRUE, sep="",  na.strings="NA", dec=".", strip.white=TRUE)
# loading txt file

LMM = lmer(Toral ~ Time +Sex +Age +BMI +EdTemp + (1 +Time| Subject), data=OralTemp, REML=FALSE)
# coding the model
# Time = time of day normalized for chronotype (this is the ‘time distance’ between the oral temperature measurement
and the subject’s MSFsc), a continuous variable specified as the proportion of a linearized unit circle, where 0=MSFsc
and 1=24 hours).
# Toral = sublingual temperature measured in each subject at 3 time points in one day
# Sex = male, luteal female, or non-luteal female
# Age = age in years at recruitment
# BMI = BMI on day of scanning
# EdTemp = ground temperature in Edinburgh at time and date of scanning
# Random effects for intercept by Subject, and for slope by Subject with respect to Time
# REML = FALSE (choosing maximum likelihood estimation)

>vif(LMM)
         GVIF Df GVIF^(1/(2*Df))
Time   1.093069  1        1.045499
Sex    1.325030  2        1.072893
Age    1.101824  1        1.049678
BMI    1.245074  1        1.115829
EdTemp 1.128850  1        1.062473
# testing variance inflation factors to check for collinearity – all ok

> summary(LMM)

Linear mixed model fit by maximum likelihood . t-tests use
  Satterthwaite's method [lmerModLmerTest]
Formula: Toral ~ Time + Sex + Age + BMI + EdTemp + (1 + Time | Subject)
   Data: OralTemp

     AIC      BIC   logLik deviance df.resid
   152.8    183.2    -65.4    130.8      106

Scaled residuals:
     Min       1Q   Median       3Q      Max
-2.44156 -0.48825  0.01042  0.60551  2.40927

Random effects:
 Groups   Name        Variance Std.Dev. Corr
 Subject  (Intercept) 0.06957  0.2638
          Time        0.02183  0.1478   0.55
 Residual             0.11725  0.3424
Number of obs: 117, groups:  Subject, 40

Fixed effects:
             Estimate Std. Error        df t value Pr(>|t|)
```

```
(Intercept)  35.034571   0.543976 51.461916  64.405   <2e-16 *** # predicted minimum OralTemp (at MSFsc) for all subjects is around 35.0°C
Time        -0.210072   0.139268 45.511215  -1.508   0.1384
SexMale      -0.303267   0.134295 40.631303  -2.258   0.0294 * # male oral temp is ~0.3°C below luteal females
SexNonluteal -0.091260   0.199003 41.491085  -0.459   0.6489
Age          0.003190   0.010572 40.564646   0.302   0.7644
BMI          0.044214   0.018915 40.858005   2.338   0.0244 *  # oral temp increases with BMI
EdTemp       0.009742   0.013472 83.672125   0.723   0.4716
---
Signif. codes:  0 '***' 0.001 '**' 0.01 '*' 0.05 '.' 0.1 ' ' 1

Correlation of Fixed Effects:
            (Intr) Time   SexMal SxNnlt Age    BMI
Time        -0.208
SexMale      0.165  0.017
SexNonlutel -0.192  0.069  0.330
Age         -0.423 -0.027 -0.162 -0.215
BMI         -0.702  0.003 -0.273  0.196 -0.176
EdTemp      -0.423  0.290  0.050  0.185  0.010  0.027

> confint(LMM)
Computing profile confidence intervals ...
                    2.5 %      97.5 %
.sig01        0.086293063  0.50958981
.sig02       -1.000000000  1.00000000
.sig03        0.000000000  0.71967056
.sigma        0.276894519  0.40596712
(Intercept)  33.951564765 36.12679402
Time         -0.487124063  0.07182091
SexMale      -0.575373534 -0.03016486
SexNonluteal -0.491599951  0.30947985
Age          -0.018278499  0.02466916
BMI           0.005646613  0.08278511
EdTemp       -0.017252937  0.03646063
```

```
plot(LMM, col="dimgray", cex=0.4, xlab="Fitted oral temperature", ylab="Residuals", grid=FALSE)
# plot to check residuals

quartz.save("Quartz 2 [*]", type = "tiff", device = dev.cur(), dpi = 300)
# graphics support for transparent colours required for some devices

qqnorm(resid(LMM), col="dimgray", cex=0.4)
qqline(resid(LMM), col="dimgray")
# QQ plot to check residual fits

plot(allEffects(LMM, residuals=TRUE), lwd=4, lines=list(col="blue"), residuals.color=yarrr::transparent("black", trans.val=.3), residuals.cex=0.8, residuals.pch=16, partial.residuals=list(smooth=TRUE, lty="dashed"), residuals.smooth.color="yellow", confint=list(alpha=0.3))
# plotting all predictor effects with residuals and partial residuals for summary overview

(plot(predictorEffect("BMI", LMM, residuals=TRUE), lwd=4, lines=list(col="blue"), confint=list(alpha=0.3), residuals.color=yarrr::transparent("black", trans.val=.3), residuals.cex=0.8, residuals.pch=16, xlab="BMI", ylab="Oral temperature", partial.residuals=list(smooth=TRUE, lty="dashed"), residuals.smooth.color="yellow"))
# plotting BMI effect only

(plot(predictorEffect("Sex", LMM, residuals=TRUE), lwd=6, lines=list(col="blue"), confint=list(col="blue", style="bars"), residuals.color=yarrr::transparent("black", trans.val=.3), residuals.cex=0.8, residuals.pch=16, xlab="Sex", ylab="Oral temperature"))
# plotting Sex effect only
```

```
(plot(predictorEffect("Time", LMM, residuals=TRUE), lwd=4, lines=list(col="blue"), confint=list(alpha=0.3),
residuals.color=yarrr::transparent("black", trans.val=.3), residuals.cex=0.8, residuals.pch=16, xlab="Time", ylab="Oral
temperature", partial.residuals=list(smooth=TRUE, lty="dashed"), residuals.smooth.color="yellow"))
# plotting Time effect only

(plot(predictorEffect("Age", LMM, residuals=TRUE), lwd=4, lines=list(col="blue"), confint=list(alpha=0.3),
residuals.color=yarrr::transparent("black", trans.val=.3), residuals.cex=0.8, residuals.pch=16, xlab="Age (y)",
ylab="Oral temperature", partial.residuals=list(smooth=TRUE, lty="dashed"), residuals.smooth.color="yellow"))
# plotting Age effect only

(plot(predictorEffect("EdTemp", LMM, residuals=TRUE), lwd=4, lines=list(col="blue"), confint=list(alpha=0.3),
residuals.color=yarrr::transparent("black", trans.val=.3), residuals.cex=0.8, residuals.pch=16, xlab="EdTemp",
ylab="Oral temperature", partial.residuals=list(smooth=TRUE, lty="dashed"), residuals.smooth.color="yellow"))
# plotting EdTemp effect only
```

**Linear mixed model for global brain temperature in healthy volunteers**

```
BrainTemp = read.table("BrainTemp.txt", header=TRUE, sep="",  na.strings="NA", dec=".", strip.white=TRUE)
#loading txt file

str(BrainTemp)
'data.frame':	8868 obs. of  8 variables:
 $ Subject    : int  1 1 1 1 1 1 1 1 1 1 ...
 $ Sex        : chr  "Luteal" "Luteal" "Luteal" "Luteal" ...
 $ Age        : num  23 23 23 23 23 23 23 23 23 23 ...
 $ BMI        : num  24.5 24.5 24.5 24.5 24.5 24.5 24.5 24.5 24.5 24.5 ...
 $ Sleep      : chr  "No" "Possible" "No" "No" ...
 $ BrainRegion: chr  "DSuperficial" "DSuperficial" "DSuperficial" "CSuperficial" ...
 $ Time       : num  0.175 0.465 0.755 0.175 0.465 ...
 $ TBrain     : num  38 38.1 38.2 38 38.1 ...

LMM = lmer(TBrain ~ Time +Sex +Age +Sleep +BrainRegion + (1 +Time| Subject), data=BrainTemp, REML=FALSE)
# coding the model
# Time = time of day normalized for chronotype (this is the 'time distance' between the oral temperature measurement
and the subject's MSF_sc), a continuous variable specified as the proportion of a linearized unit circle, where 0=MSF_sc
and 1=24 hours).
# TBrain = brain temperature measured in each subject at each brain voxel at 3 time points in one day
# Sex = male, luteal female, or non-luteal female
# Age = age in years at recruitment
# Sleep = whether subject reported falling asleep during scan (yes, no, maybe)
# BrainRegion = one of 6 brain MRS regions (Superficial regions A, B, C, D, Thalamus, Hypothalamus)
# Random effects for intercept by Subject, and for slope by Subject with respect to Time
# REML = FALSE (choosing maximum likelihood estimation)

>vif(LMM)
                GVIF Df GVIF^(1/(2*Df))
Time        1.005456  1        1.002724
Sex         1.084022  2        1.020374
Age         1.084473  1        1.041381
Sleep       1.010824  2        1.002695
BrainRegion 1.000027  5        1.000003
#testing variance inflation factors to check for collinearity – all ok

> summary(LMM)

Linear mixed model fit by maximum likelihood . t-tests use
  Satterthwaite's method [lmerModLmerTest]
Formula: TBrain ~ Time + Sex + Age + Sleep + BrainRegion + (1 + Time |
    Subject)
```

```
 Data: BrainTemp

     AIC      BIC   logLik deviance df.resid
  8875.4   8988.8  -4421.7   8843.4     8828

Scaled residuals:
    Min      1Q  Median      3Q     Max
-5.8415 -0.6122  0.0081  0.6359  5.9689

Random effects:
 Groups   Name        Variance Std.Dev. Corr
 Subject  (Intercept) 0.1173   0.3425
          Time        0.2789   0.5281   -0.86
 Residual             0.1543   0.3928
Number of obs: 8844, groups:  Subject, 39

Fixed effects:
                          Estimate Std. Error        df t value Pr(>|t|)
(Intercept)              3.790e+01  1.619e-01  4.566e+01 234.124  < 2e-16 # predicted minimum global BrainTemp (at MSFsc) for all subjects is ~37.9°C
Time                    -5.724e-01  8.720e-02  3.836e+01  -6.564 9.21e-08 # diurnal variation of  ~0.57°C across unit circle (24h)
SexLutealNon            -3.588e-01  9.517e-02  3.817e+01  -3.770 0.000553 # global BrainTemp ~0.36°C lower in follicular phase females relative to luteal phase females
SexMale                 -3.558e-01  6.457e-02  3.824e+01  -5.510 2.63e-06 # global BrainTemp ~0.36°C lower in males relative to luteal phase females
Age                      1.502e-02  5.127e-03  3.846e+01   2.930 0.005670 # global BrainTemp increases by ~0.02°C with each unit (year) of age across this cohort
SleepPossible           -4.625e-02  3.151e-02  6.400e+03  -1.468 0.142266 # global BrainTemp not significantly different in those who might have slept versus those that did not sleep
SleepYes                 1.125e-01  1.894e-02  5.930e+03   5.940 3.00e-09 # global BrainTemp ~0.11°C higher in those that reported sleeping during the scan versus those that did not
BrainRegionBSuperficial  6.739e-01  1.334e-02  8.765e+03  50.514  < 2e-16 # Layer B ~0.67°C higher than layer A
BrainRegionCSuperficial  8.507e-01  1.279e-02  8.765e+03  66.522  < 2e-16 # Layer C ~0.85°C higher than layer A
BrainRegionDSuperficial  6.528e-01  1.262e-02  8.766e+03  51.734  < 2e-16 # Layer D ~0.65°C higher than layer A
BrainRegionHypothalamus  1.078e+00  3.814e-02  8.765e+03  28.278  < 2e-16 # Hypothalamus ~1.08°C higher than layer A
BrainRegionThalamus      1.642e+00  3.814e-02  8.765e+03  43.049  < 2e-16 # Thalamus ~1.64°C higher than layer A

(Intercept)             ***
Time                    ***
SexLutealNon            ***
SexMale                 ***
Age                     **
SleepPossible
SleepYes                ***
BrainRegionBSuperficial ***
BrainRegionCSuperficial ***
BrainRegionDSuperficial ***
BrainRegionHypothalamus ***
BrainRegionThalamus     ***
---
Signif. codes:  0 ‘***’ 0.001 ‘**’ 0.01 ‘*’ 0.05 ‘.’ 0.1 ‘ ’ 1

Correlation of Fixed Effects:
            (Intr) Time   SxLtlN SexMal Age    SlpPss SlepYs BrnRBS
Time        -0.306
SexLutealNn  0.040 -0.003
SexMale      0.006  0.006  0.422
Age         -0.906  0.011 -0.208 -0.250
```

```
SleepPossbl  0.006  0.019  0.016 -0.026 -0.024
SleepYes     0.038 -0.066  0.034  0.019 -0.055  0.162
BrnRgnBSprf -0.047  0.000  0.000  0.000  0.000  0.000  0.000
BrnRgnCSprf -0.049  0.000  0.000  0.000  0.000  0.000  0.000  0.589
BrnRgnDSprf -0.049  0.000  0.002 -0.001  0.000  0.000 -0.001  0.597
BrnRgnHypth -0.016  0.000  0.000  0.000  0.000  0.000  0.000  0.198
BrnRgnThlms -0.016  0.000  0.000  0.000  0.000  0.000  0.000  0.198
            BrnRCS BrnRDS BrnRgH
Time
SexLutealNn
SexMale
Age
SleepPossbl
SleepYes
BrnRgnBSprf
BrnRgnCSprf
BrnRgnDSprf  0.622
BrnRgnHypth  0.206  0.208
BrnRgnThlms  0.206  0.208  0.069
```

```
> confint(LMM)

Computing profile confidence intervals ...
                              2.5 %      97.5 %
.sig01                  0.274283463  0.44046140
.sig02                 -0.923902085 -0.73767152
.sig03                  0.422787477  0.67945006
.sigma                  0.387104810  0.39873724
(Intercept)            37.562384575 38.23685713
Time                   -0.747546353 -0.39691142
SexLutealNon           -0.550016709 -0.16709918
SexMale                -0.486451011 -0.22545069
Age                     0.004196089  0.02578471
SleepPossible          -0.108068257  0.01562339
SleepYes                0.074971478  0.15018257
BrainRegionBSuperficial  0.647710310  0.70000779
BrainRegionCSuperficial  0.825612445  0.87574561
BrainRegionDSuperficial  0.628042895  0.67750973
BrainRegionHypothalamus  1.003664525  1.15317024
BrainRegionThalamus      1.566971542  1.71647726
```

plot(LMM, col="dimgray", cex=0.1, xlab="Fitted brain temperature", ylab="Residuals", grid=FALSE)
*# plots residuals - looks ok at high resolution*

quartz.save("Quartz 2 [*]", type = "tiff", device = dev.cur(), dpi = 300)
*# graphics support for transparent colours required for some devices*

qqnorm(resid(LMM), col="dimgray", cex=0.25)
qqline(resid(LMM), col="dimgray")
*# QQ plot shows symmetric distribution with fat tails = some degree of kurtosis but not enough to rule out normal distribution*

plot(allEffects(LMM, residuals=TRUE), lwd=4, lines=list(col="red"), residuals.color=yarrr::transparent("black", trans.val=.3), residuals.cex=0.2, residuals.pch=16, partial.residuals=list(smooth=TRUE, lty="dashed", lwd=2), residuals.smooth.color="yellow", confint=list(alpha=0.3))
*# plotting all predictor effects with residuals and partial residuals for summary overview*

```
(plot(predictorEffect("Age", LMM, residuals=TRUE), lwd=4, lines=list(col="red"), confint=list(alpha=0.3),
residuals.color=yarrr::transparent("black", trans.val=.3), residuals.cex=0.2, residuals.pch=16, xlab="Age (y)",
ylab="Brain temperature", partial.residuals=list(smooth=TRUE, lty="dashed", lwd=2),
residuals.smooth.color="yellow"))
```
*# plotting Age effect only*

```
(plot(predictorEffect("Sex", LMM, residuals=TRUE), lwd=6, lines=list(col="red"), confint=list(col="red", style="bars"),
residuals.color=yarrr::transparent("black", trans.val=.8), residuals.cex=0.2, residuals.pch=16, xlab="Sex", ylab="Brain
temperature"))
```
*# plotting Sex effect only*

```
(plot(predictorEffect("Time", LMM, residuals=TRUE), lwd=4, lines=list(col="red"), confint=list(alpha=0.3),
residuals.color=yarrr::transparent("black", trans.val=.3), residuals.cex=0.2, residuals.pch=16, xlab="Time", ylab="Brain
temperature", partial.residuals=list(smooth=TRUE, lty="dashed", lwd=2), residuals.smooth.color="yellow"))
```
*# plotting Time effect only*

```
(plot(predictorEffect("Sleep", LMM, residuals=TRUE), lwd=6, lines=list(col="red"), confint=list(col="red",
style="bars"), residuals.color=yarrr::transparent("black", trans.val=.8), residuals.cex=0.2, residuals.pch=16,
xlab="Sleep", ylab="Brain temperature"))
```
*# plotting Sleep effect only*

```
(plot(predictorEffect("BrainRegion", LMM, residuals=TRUE), lwd=6, lines=list(col="red"), confint=list(col="red",
style="bars"), residuals.color=yarrr::transparent("black", trans.val=.5), residuals.cex=0.2, residuals.pch=16, xlab="Brain
region", ylab="Brain temperature"))
```
*# plotting BrainRegion effect only*

```
LMM = lmer(TBrain ~ Time +Sex +Age +BMI +BrainRegion + (1 +Time| Subject), data=BrainTemp, REML=FALSE)
```
*#replacing fixed effect ‘Sleep’ with ‘BMI’*

```
>vif(LMM)

GVIF Df GVIF^(1/(2*Df))
Time        1.003045  1        1.001521
Sex         1.294049  2        1.066566
Age         1.122437  1        1.059451
BMI         1.255759  1        1.120607
BrainRegion 1.000000  1        1.000000
```
*#testing variance inflation factors to check for collinearity – all ok*

```
> summary(LMM)

Linear mixed model fit by maximum likelihood . t-tests use
  Satterthwaite's method [lmerModLmerTest]
Formula:
TBrain ~ Time + Sex + Age + BMI + BrainRegion + (1 + Time | Subject)
   Data: BrainTemp

     AIC      BIC   logLik deviance df.resid
   506.7    544.4   -242.4    484.7      215

Scaled residuals:
    Min      1Q  Median      3Q     Max
-2.1699 -0.6736 -0.0181  0.6044  3.1712

Random effects:
 Groups   Name        Variance Std.Dev. Corr
 Subject  (Intercept) 0.1561   0.3951
          Time        0.2660   0.5158   -0.61
 Residual             0.4174   0.6460
Number of obs: 226, groups:  Subject, 38
```

```
Fixed effects:
                      Estimate Std. Error         df t value Pr(>|t|)
(Intercept)          38.883586   0.559424  40.533015  69.506  < 2e-16 ***
Time                 -0.878656   0.197539  37.305782  -4.448 7.54e-05 ***
SexLutealNon         -0.753159   0.223372  37.286577  -3.372 0.001751 **
SexMale              -0.605066   0.154559  37.607107  -3.915 0.000368 ***
Age                   0.025862   0.012093  37.745055   2.139 0.039006 *
BMI                   0.004784   0.021476  38.184096   0.223 0.824901    #Global brain temperature does not vary with BMI
BrainRegionThalamus   0.559667   0.085947 153.246981   6.512 1.01e-09 ***
---
Signif. codes:  0 '***' 0.001 '**' 0.01 '*' 0.05 '.' 0.1 ' ' 1

Correlation of Fixed Effects:
            (Intr) Time   SxLtlN SexMal Age    BMI
Time        -0.150
SexLutealNn -0.134  0.025
SexMale      0.196 -0.003  0.341
Age         -0.435 -0.048 -0.242 -0.171
BMI         -0.754  0.002  0.209 -0.261 -0.199
BrnRgnThlms -0.077  0.000  0.000  0.000  0.000  0.000

>confint(LMM)

Computing profile confidence intervals ...
                          2.5 %      97.5 %
.sig01               0.023080184  0.70715187
.sig02              -1.000000000  1.00000000
.sig03               0.000000000  1.10756097
.sigma               0.579867609  0.72543846
(Intercept)         37.746597713 40.02491753
Time                -1.275906313 -0.48075480
SexLutealNon        -1.212342177 -0.29571036
SexMale             -0.922751101 -0.28816908
Age                  0.001462829  0.05016401
BMI                 -0.038820467  0.04825773
BrainRegionThalamus  0.390153101  0.72918141

(plot(predictorEffect("BMI", LMM, residuals=TRUE), lwd=4, lines=list(col="red"), confint=list(alpha=0.3),
residuals.color=yarrr::transparent("black", trans.val=.3), residuals.cex=0.2, residuals.pch=16, xlab="BMI", ylab="Brain
temperature", partial.residuals=list(smooth=TRUE, lty="dashed", lwd=2), residuals.smooth.color="yellow"))
```

**Linear mixed model for global brain temperature amplitude in healthy volunteers**

```
BrainTempAmp = read.table("BrainTempAmp.txt", header=TRUE, sep="",  na.strings="NA", dec=".",
strip.white=TRUE)
#loading txt file

>str(BrainTempAmp)

'data.frame':         2956 obs. of  8 variables:
 $ Subject    : int  1 1 1 1 1 1 1 1 1 1 ...
 $ Sex        : chr  "Luteal" "Luteal" "Luteal" "Luteal" ...
 $ Age        : num  23 23 23 23 23 23 23 23 23 23 ...
 $ BMI        : num  24.5 24.5 24.5 24.5 24.5 24.5 24.5 24.5 24.5 24.5 ...
 $ Sleep      : chr  "No" "No" "No" "No" ...
 $ BrainRegion: chr  "BSuperficial" "CSuperficial" "DSuperficial" "CSuperficial" ...
 $ Time       : num  0.755 0.755 0.755 0.755 0.755 ...
 $ TBrainAmp  : num  0.0262 0.0433 0.0507 0.0696 0.1124 ...
```

```
LMM = lmer(TBrainAmp ~ Time +Sex +Age +BMI +BrainRegion + (1 +Time| Subject), data=BrainTempAmp,
REML=FALSE)
#replacing absolute brain temperature with brain temperature amplitude over time (temporal range across the day)

>vif(LMM)

            GVIF Df GVIF^(1/(2*Df))
Time        1.656332  1        1.286986
Sex         1.508782  2        1.108298
Age         1.562454  1        1.249982
BMI         1.287352  1        1.134615
BrainRegion 1.000049  5        1.000005

> summary(LMM)

Linear mixed model fit by maximum likelihood . t-tests use
  Satterthwaite's method [lmerModLmerTest]
Formula: TBrainAmp ~ Time + Sex + Age + BMI + BrainRegion + (1 + Time |
    Subject)
   Data: BrainTempAmp

     AIC      BIC   logLik deviance df.resid
  -759.4   -669.5    394.7   -789.4     2941

Scaled residuals:
    Min      1Q  Median      3Q     Max
-4.9971 -0.5695 -0.0675  0.4387  7.8182

Random effects:
 Groups   Name        Variance Std.Dev. Corr
 Subject  (Intercept) 0.81404  0.9022
          Time        0.86191  0.9284   -1.00
 Residual             0.04276  0.2068
Number of obs: 2956, groups:  Subject, 38

Fixed effects:
                          Estimate Std. Error        df t value Pr(>|t|)
(Intercept)             -3.675e-02  3.743e-01  2.906e+01  -0.098    0.922
Time                     3.317e-01  4.911e-01  3.610e+01   0.675    0.504
SexLutealNon             7.173e-03  9.154e-02  5.006e+01   0.078    0.938
SexMale                  4.398e-02  5.118e-02  2.657e+01   0.859    0.398
Age                      3.903e-03  4.644e-03  3.063e+01   0.840    0.407
BMI                      4.272e-03  6.959e-03  2.267e+01   0.614    0.545
BrainRegionBSuperficial -1.257e-02  1.216e-02  2.920e+03  -1.034    0.301
BrainRegionCSuperficial  1.033e-02  1.166e-02  2.920e+03   0.886    0.376
BrainRegionDSuperficial  5.733e-02  1.149e-02  2.920e+03   4.992 6.33e-07
BrainRegionHypothalamus  7.349e-01  3.477e-02  2.920e+03  21.136  < 2e-16
BrainRegionThalamus      4.930e-01  3.477e-02  2.920e+03  14.180  < 2e-16

(Intercept)
Time
SexLutealNon
SexMale
Age
BMI
BrainRegionBSuperficial
BrainRegionCSuperficial
BrainRegionDSuperficial ***
BrainRegionHypothalamus ***
```

```
BrainRegionThalamus     ***
---
Signif. codes:  0 '***' 0.001 '**' 0.01 '*' 0.05 '.' 0.1 ' ' 1

Correlation of Fixed Effects:
            (Intr) Time   SxLtlN SexMal Age    BMI    BrnRBS BrnRCS
Time        -0.869
SexLutealNn -0.301  0.266
SexMale      0.236 -0.119  0.262
Age          0.211 -0.486 -0.247 -0.192
BMI         -0.284 -0.090  0.145 -0.254 -0.124
BrnRgnBSprf -0.019  0.000  0.000  0.000  0.000  0.000
BrnRgnCSprf -0.019  0.000 -0.001  0.000  0.000  0.000  0.589
BrnRgnDSprf -0.018 -0.001  0.003 -0.001  0.000 -0.001  0.598  0.623
BrnRgnHypth -0.007  0.000  0.000  0.000  0.000  0.000  0.198  0.206
BrnRgnThlms -0.007  0.000  0.000  0.000  0.000  0.000  0.198  0.206
            BrnRDS BrnRgH
Time
SexLutealNn
SexMale
Age
BMI
BrnRgnBSprf
BrnRgnCSprf
BrnRgnDSprf
BrnRgnHypth  0.209
BrnRgnThlms  0.209  0.069
convergence code: 0
boundary (singular) fit: see ?isSingular
```

```
>confint(LMM)

Computing profile confidence intervals ...
                                2.5 %     97.5 %
.sig01                    0.877384844 0.93626871
.sig02                   -1.000000000 1.00000000
.sig03                    0.807360811 0.95781837
.sigma                    0.201594024 0.21220870
(Intercept)              -0.798999336 0.72294354
Time                     -0.655490296 1.32758198
SexLutealNon             -0.177191135 0.19263988
SexMale                  -0.059603628 0.15075616
Age                      -0.005453221 0.01338030
BMI                      -0.011808158 0.01828080
BrainRegionBSuperficial  -0.036419840 0.01127112
BrainRegionCSuperficial  -0.012530144 0.03318714
BrainRegionDSuperficial   0.034814684 0.07985178
BrainRegionHypothalamus   0.666703486 0.80304028
BrainRegionThalamus       0.424853486 0.56119028
There were 50 or more warnings (use warnings() to see the first 50)
```

```
(plot(predictorEffect("Age", LMM, residuals=TRUE), lwd=4, lines=list(col="red"), confint=list(alpha=0.3), residuals.color=yarrr::transparent("black", trans.val=.3), residuals.cex=0.2, residuals.pch=16, xlab="Age", ylab="Brain temperature amplitude", partial.residuals=list(smooth=TRUE, lty="dashed", lwd=2), residuals.smooth.color="yellow"))
```

**Linear mixed model for deep brain temperature in healthy volunteers**

```
BrainTemp = read.table("BrainTempDeep.txt", header=TRUE, sep="",  na.strings="NA", dec=".", strip.white=TRUE)
# loading txt file
```

```
str(BrainTemp)
'data.frame':   226 obs. of  8 variables:
 $ Subject    : int  1 1 1 1 1 1 2 2 2 2 ...
 $ Sex        : chr  "Luteal" "Luteal" "Luteal" "Luteal" ...
 $ Age        : num  23 23 23 23 23 ...
 $ BMI        : num  24.5 24.5 24.5 24.5 24.5 24.5 29.5 29.5 29.5 29.5 ...
 $ Sleep      : chr  "No" "Possible" "No" "No" ...
 $ BrainRegion: chr  "Thalamus" "Thalamus" "Thalamus" "Hypothalamus" ...
 $ Time       : num  0.175 0.465 0.755 0.175 0.465 ...
 $ TBrain     : num  39 38.7 39.6 40.3 39.8 ...
```

```
LMM = lmer(TBrain ~ Time +Sex +Age +Sleep +BrainRegion + (1 +Time| Subject), data=BrainTemp, REML=FALSE)
```

*# coding the model*
*# Time = time of day normalized for chronotype (this is the 'time distance' between the oral temperature measurement and the subject's $MSF_{sc}$), a continuous variable specified as the proportion of a linearized unit circle, where 0=$MSF_{sc}$ and 1=24 hours).*
*# TBrain = brain temperature measured in each subject at each deep brain voxel at 3 time points in one day*
*# Sex = male, luteal female, or non-luteal female*
*# Age = age in years at recruitment*
*# Sleep = whether subject reported falling asleep during scan (yes, no, maybe)*
*# BrainRegion = one of two deep brain MRS voxels (Thalamus, Hypothalamus)*
*# Random effects for intercept by Subject, and for slope by Subject with respect to Time*
*# REML = FALSE (choosing maximum likelihood estimation)*

```
>vif(LMM)
                GVIF Df GVIF^(1/(2*Df))
Time        1.040578  1        1.020087
Sex         1.108491  2        1.026084
Age         1.106316  1        1.051815
Sleep       1.092063  2        1.022261
BrainRegion 1.000000  1        1.000000
```

*# testing variance inflation factors to check for collinearity – all ok*

```
> summary(LMM)

Linear mixed model fit by maximum likelihood . t-tests use
  Satterthwaite's method [lmerModLmerTest]
Formula: TBrain ~ Time + Sex + Age + Sleep + BrainRegion + (1 + Time |
    Subject)
   Data: BrainTemp

     AIC      BIC   logLik deviance df.resid
   505.6    546.7   -240.8    481.6      214

Scaled residuals:
    Min      1Q  Median      3Q     Max
-2.1642 -0.6011 -0.0333  0.6194  3.1716

Random effects:
 Groups   Name        Variance Std.Dev. Corr
 Subject  (Intercept) 0.1950   0.4416
          Time        0.2438   0.4938   -0.71
 Residual             0.4110   0.6411
Number of obs: 226, groups:  Subject, 38

Fixed effects:
              Estimate Std. Error       df t value Pr(>|t|)
(Intercept)   38.94911    0.37156 43.38210 104.825  < 2e-16 ***
```

*# predicted minimum for deep regions at $MSF_{sc}$ is ~38.9°C*

```
Time              -0.85561    0.19844  38.12657  -4.312 0.000110 *** # diurnal variation of ~0.86°C in deep regions
across a unit circle (24h)
SexLutealNon       -0.82053    0.22092  38.00242  -3.714 0.000653 *** # deep brain temp ~0.82°C higher in luteal
phase females than follicular phase females
SexMale            -0.60253    0.15079  38.31558  -3.996 0.000283 *** # deep brain temp ~0.60°C higher in luteal phase
females than males
Age                0.02920    0.01204  39.01803   2.426 0.019985 *   # ~0.03°C increase in deep brain temp per year of age
in this cohort
SleepPossible      -0.29299    0.21706 157.70181  -1.350 0.179021    # no significant effect of reported possible sleep on
deep brain temp
SleepYes           -0.18707    0.12875 164.22369  -1.453 0.148135    # no significant effect of reported sleep on deep
brain temp
BrainRegionThalamus   0.55967    0.08529 152.87677   6.562 7.77e-10 *** # thalamus ~0.56°C higher than
hypothalamus
---
Signif. codes:  0 '***' 0.001 '**' 0.01 '*' 0.05 '.' 0.1 ' ' 1

Correlation of Fixed Effects:
            (Intr) Time   SxLtlN SexMal Age    SlpPss SlepYs
Time        -0.247
SexLutealNn  0.042  0.013
SexMale      0.007 -0.019  0.413
Age         -0.905 -0.027 -0.218 -0.237
SleepPossbl  0.008  0.075  0.045 -0.103 -0.060
SleepYes     0.099 -0.162  0.092  0.032 -0.150  0.145
BrnRgnThlms -0.115  0.000  0.000  0.000  0.000  0.000  0.000

> confint(LMM)

Computing profile confidence intervals ...
                          2.5 %      97.5 %
.sig01               0.094841077  0.75856893
.sig02              -1.000000000  1.00000000
.sig03               0.000000000  1.09278289
.sigma               0.575332896  0.71964104
(Intercept)         38.200358244 39.69442448
Time                -1.257067569 -0.45790353
SexLutealNon        -1.276551404 -0.36850999
SexMale             -0.917553794 -0.28941499
Age                  0.005027576  0.05347177
SleepPossible       -0.747267324  0.15518136
SleepYes            -0.442360067  0.07043526
BrainRegionThalamus  0.391449742  0.72788477

plot(LMM, col="dimgray", cex=0.4, xlab="Fitted brain temperature", ylab="Residuals", grid=FALSE)
# plots residuals to check

quartz.save("Quartz 2 [*]", type = "tiff", device = dev.cur(), dpi = 300)
# graphics support for transparent colours required for some devices

qqnorm(resid(LMM), col="dimgray", cex=0.4)
qqline(resid(LMM), col="dimgray")
# QQ plot shows symmetric distribution with fat tails = some degree of kurtosis but not enough to rule out normal
distribution

plot(allEffects(LMM, residuals=TRUE), lwd=4, lines=list(col="red"), residuals.color=yarrr::transparent("black",
trans.val=.3), residuals.cex=0.2, residuals.pch=16, partial.residuals=list(smooth=TRUE, lty="dashed", lwd=2),
residuals.smooth.color="yellow", confint=list(alpha=0.3))
# plotting all predictor effects with residuals and partial residuals for summary overview
```

```
(plot(predictorEffect("Age", LMM, residuals=TRUE), lwd=4, lines=list(col="red"), confint=list(alpha=0.3), residuals.color=yarrr::transparent("black", trans.val=.3), residuals.cex=0.4, residuals.pch=16, xlab="Age (y)", ylab="Brain temperature", partial.residuals=list(smooth=TRUE, lty="dashed", lwd=2), residuals.smooth.color="yellow"))
# plotting Age effect only

(plot(predictorEffect("Sex", LMM, residuals=TRUE), lwd=6, lines=list(col="red"), confint=list(col="red", style="bars"), residuals.color=yarrr::transparent("black", trans.val=.3), residuals.cex=0.4, residuals.pch=16, xlab="Sex", ylab="Brain temperature"))
# plotting Sex effect only

(plot(predictorEffect("Time", LMM, residuals=TRUE), lwd=4, lines=list(col="red"), confint=list(alpha=0.3), residuals.color=yarrr::transparent("black", trans.val=.3), residuals.cex=0.4, residuals.pch=16, xlab="Time", ylab="Brain temperature", partial.residuals=list(smooth=TRUE, lty="dashed", lwd=2), residuals.smooth.color="yellow"))
# plotting Time effect only

(plot(predictorEffect("Sleep", LMM, residuals=TRUE), lwd=6, lines=list(col="red"), confint=list(col="red", style="bars"), residuals.color=yarrr::transparent("black", trans.val=.3), residuals.cex=0.4, residuals.pch=16, xlab="Sleep", ylab="Brain temperature"))
# plotting Sleep effect only

(plot(predictorEffect("BrainRegion", LMM, residuals=TRUE), lwd=6, lines=list(col="red"), confint=list(col="red", style="bars"), residuals.color=yarrr::transparent("black", trans.val=.3), residuals.cex=0.4, residuals.pch=16, xlab="Brain region", ylab="Brain temperature"))
# plotting BrainRegion effect only
```

**Generalized linear mixed model for outcome in TBI patients**

```
Outcome = read.table("Outcome.txt", header=TRUE, sep="\t", na.strings="NA", dec=".", strip.white=TRUE)

str(Outcome)

'data.frame':	114 obs. of  15 variables:
 $ Subject   : chr  "P6" "P83" "P16" "P21" ... # Patient ID number
 $ Age       : int  16 16 17 17 17 19 19 19 19 20 ... # Patient age in years
 $ Sex       : chr  "Male" "Male" "Male" "Male" ... # Patient sex (menstrual cycle phase not known)
 $ Outcome   : int  0 1 0 0 0 0 0 0 0 0 ... # Patient outcome in intensive care: 0=alive, 1=dead
 $ BrainMean : num  36.9 37.7 37.5 39.1 36.6 ... # Absolute mean brain temperature for patient across processed data set (after excluding artefacts etc)
 $ BrainRange: num  2.8 3.38 4.8 3.88 2.44 NA 3.32 2.92 3.94 2.66 ... # Range in patient brain temperature across processed data set
 $ BodyRange : num  NA 2.4 4 2.8 3.2 4.7 3.5 1.9 1.8 2 ... # Range in patient body temperature across processed data set
 $ BrainMax  : num  38.4 39.6 40.6 40.3 38 ... # Maximum patient brain temperature across processed data set
 $ BrainMin  : num  35.6 36.3 35.8 36.5 35.6 ... # Minimum patient brain temperature across processed data set
 $ BodyMax   : num  NA 38.4 39 38.3 37.4 38.5 40.2 38.4 38.7 39.6 ... # Maximum patient body temperature across processed data set
 $ BodyMin   : num  NA 36 35 35.5 34.2 33.8 36.7 36.5 36.9 37.6 ... # Minimum patient body temperature across processed data set
 $ Diurnal   : chr  "Yes" "No" "No" "Yes" ... # Was there evidence of diurnal rhythmicity in temperature across processed data set? Yes or No
 $ PLR       : int  2 2 1 2 NA 2 2 2 2 2 ... # Pupillary light reflex present in both eyes (2),one eye (1), or no eyes (0)
 $ GCS       : int  9 7 14 4 NA 6 6 7 15 6 ... # Glasgow Coma Scale score
 $ GCSM      : int  5 3 6 2 NA 3 4 5 6 4 ... # Glasgow Coma Scale Motor response score

GLMM = glmer(Outcome ~ Age +Sex + BrainMean +BrainRange +Diurnal + (1|Subject), data=Outcome, family=binomial, nAGQ=0)
# coding the model
```

*# model includes several fixed effects plus random effects for intercept by Subject*
*# there were no significant relationships between maximum BrainTemp, minimum BrainTemp, maximum BodyTemp, minimum BodyTemp, PLR, GCS or GCSM and Outcome and including these fixed effects did not improve the model fit*

```
>summary(GLMM)

Generalized linear mixed model fit by maximum likelihood (Adaptive
  Gauss-Hermite Quadrature, nAGQ = 0) [glmerMod]
 Family: binomial  ( logit )
Formula: Outcome ~ Age + Sex + BrainMean + BrainRange + Diurnal + (1 |
    Subject)
   Data: Outcome

     AIC      BIC   logLik deviance df.resid
    88.7    107.0    -37.4     74.7       93

Scaled residuals:
    Min       1Q  Median       3Q      Max
-2.7359 -0.4460 -0.2028 -0.0298  6.2977

Random effects:
 Groups  Name        Variance  Std.Dev.
 Subject (Intercept) 3.674e-17 6.061e-09
Number of obs: 100, groups:  Subject, 100

Fixed effects:
            Estimate Std. Error z value Pr(>|z|)
(Intercept) 23.46074   14.57763   1.609 0.107536
Age          0.09477    0.02554   3.711 0.000207 ***
SexMale      0.83033    0.75493   1.100 0.271387
BrainMean   -0.79984    0.38987  -2.052 0.040210 *
BrainRange   0.19476    0.22753   0.856 0.391997
DiurnalYes  -2.45258    1.16209  -2.110 0.034816 *
---
Signif. codes:  0 ‘***’ 0.001 ‘**’ 0.01 ‘*’ 0.05 ‘.’ 0.1 ‘ ’ 1

Correlation of Fixed Effects:
           (Intr) Age    SexMal BranMn BrnRng
Age         0.198
SexMale     0.220  0.221
BrainMean  -0.991 -0.311 -0.282
BrainRange -0.184  0.302  0.139  0.095
DiurnalYes -0.094 -0.215  0.047  0.107 -0.045

plot(GLMM, col="dimgray", cex=0.8, xlab="Mortality", ylab="Residuals", grid=FALSE)
```

*# plots residuals – as expected*

```
quartz.save("Quartz 2 [*]", type = "tiff", device = dev.cur(), dpi = 300)
```

*# graphics support for transparent colours required for some devices*

```
qqnorm(resid(GLMM), col="dimgray", cex=0.8)
qqline(resid(GLMM), col="dimgray")
```

*# QQ plot*

```
plot(allEffects(GLMM, residuals=TRUE), lwd=4, lines=list(col="purple"), residuals.color=yarrr::transparent("black",
trans.val=.3), residuals.cex=0.2, residuals.pch=16, partial.residuals=list(smooth=TRUE, lty="dashed", lwd=2),
residuals.smooth.color="yellow", confint=list(alpha=0.3))
```

*# plots all predictor effects for summary overview*

```
(plot(predictorEffect("Age", GLMM, residuals=TRUE), lwd=4, lines=list(col="purple"), confint=list(alpha=0.3),
residuals.color=yarrr::transparent("black", trans.val=.3), residuals.cex=0.6, residuals.pch=16, xlab="Age",
ylab="Mortality", partial.residuals=list(smooth=TRUE, lty="dashed", lwd=2), residuals.smooth.color="yellow"))
# plots Age effect only

(plot(predictorEffect("Diurnal", GLMM, residuals=TRUE), lwd=4, lines=list(col="purple"), confint=list(alpha=0.3),
residuals.color=yarrr::transparent("black", trans.val=.3), residuals.cex=0.6, residuals.pch=16, xlab="Diurnal?",
ylab="Mortality"))
# plots Diurnal effect only

(plot(predictorEffect("BrainRange", GLMM, residuals=TRUE), lwd=4, lines=list(col="purple"), confint=list(alpha=0.3),
residuals.color=yarrr::transparent("black", trans.val=.3), residuals.cex=0.6, residuals.pch=16, xlab="BrainRange",
ylab="Mortality", partial.residuals=list(smooth=TRUE, lty="dashed", lwd=2), residuals.smooth.color="yellow"))
# plots BrainRange effect only (not significant)

(plot(predictorEffect("BrainMean", GLMM, residuals=TRUE), lwd=4, lines=list(col="purple"), confint=list(alpha=0.3),
residuals.color=yarrr::transparent("black", trans.val=.3), residuals.cex=0.6, residuals.pch=16, xlab="BrainMean",
ylab="Mortality", partial.residuals=list(smooth=TRUE, lty="dashed", lwd=2), residuals.smooth.color="yellow"))
# plots BrainMean temperature effect only

(plot(predictorEffect("Sex", GLMM, residuals=TRUE), lwd=4, lines=list(col="purple"), confint=list(alpha=0.3),
residuals.color=yarrr::transparent("black", trans.val=.3), residuals.cex=0.6, residuals.pch=16, xlab="Sex",
ylab="Mortality"))
# plots Sex effect only (not significant)

exp(0.09477) # calculating Age odds for death
[1] 1.099406

exp(-0.79984) # calculating BrainMean temperature odds for death
[1] 0.4494009

exp(-2.45258) # calculating Diurnal odds for death
[1] 0.08607124

1/0.08607124 # calculating Diurnal odds for survival
[1] 11.61828

# calculate 95% confidence intervals using estimate +/- (1.96*standard error) for each of the significant predictors

LogAgeCI = 1.96*0.02554
print(LogAgeCI)
[1] 0.0500584 # so 95% CI for Age in log odds = 0.09477+/- 0.0500584

0.09477+LogAgeCI
[1] 0.1448284

0.09477-LogAgeCI
[1] 0.0447116 # now convert to CI on the odds

exp(c(0.0447116, 0.1448284))
[1] 1.045726 1.155841# good – it doesn't include 1
# CIs are numerically asymmetric once turned back into odds

LogBrainMeanCI = 1.96* 0.38987
print(LogBrainMeanCI)
[1] 0.7641452 # so 95% CI for BrainMean in log odds = -0.79984+/- 0.7641452

-0.79984+LogBrainMeanCI
[1] -0.0356948
```

-0.79984-LogBrainMeanCI
[1] -1.563985 # *now convert to CI on the odds*

exp(c(-1.563985, -0.0356948))
[1] 0.2093003 0.9649347 # *good – it doesn't include 1*
### *CIs are numerically asymmetric once turned back into odds*

LogDiurnalCI = 1.96* 1.16209
print(LogDiurnalCI)
[1] 2.277696 # *so 95% CI for Diurnal in log odds = -2.45258+/- 2.277696*

-2.45258+LogDiurnalCI
[1] -0.1748836

-2.45258-LogDiurnalCI
[1] -4.730276 # *now convert to CI on the odds*

exp(c(-4.730276, -0.1748836))
[1] 0.008824035 0.839554739 # *good – it doesn't include 1*
### *CIs are numerically asymmetric once turned back into odds*
